# Fine-scale spatial and social patterns of SARS-CoV-2 transmission from identical pathogen sequences

**DOI:** 10.1101/2024.05.24.24307811

**Authors:** Cécile Tran-Kiem, Miguel I. Paredes, Amanda C. Perofsky, Lauren A. Frisbie, Hong Xie, Kevin Kong, Amelia Weixler, Alexander L. Greninger, Pavitra Roychoudhury, JohnAric M. Peterson, Andrew Delgado, Holly Halstead, Drew MacKellar, Philip Dykema, Luis Gamboa, Chris D. Frazar, Erica Ryke, Jeremy Stone, David Reinhart, Lea Starita, Allison Thibodeau, Cory Yun, Frank Aragona, Allison Black, Cécile Viboud, Trevor Bedford

## Abstract

Pathogen genomics can provide insights into underlying infectious disease transmission patterns, but new methods are needed to handle modern large-scale pathogen genome datasets and realize this full potential. In particular, genetically proximal viruses should be highly informative about transmission events as genetic proximity indicates epidemiological linkage. Here, we leverage pairs of identical sequences to characterise fine-scale transmission patterns using 114,298 SARS-CoV-2 genomes collected through Washington State (USA) genomic sentinel surveillance with associated age and residence location information between March 2021 and December 2022. This corresponds to 59,660 sequences with another identical sequence in the dataset. We find that the location of pairs of identical sequences is highly consistent with expectations from mobility and social contact data. Outliers in the relationship between genetic and mobility data can be explained by SARS-CoV-2 transmission between postal codes with male prisons, consistent with transmission between prison facilities. We find that transmission patterns between age groups vary across spatial scales. Finally, we use the timing of sequence collection to understand the age groups driving transmission. Overall, this work improves our ability to leverage large pathogen genome datasets to understand the determinants of infectious disease spread.

## Introduction

Pathogen transmission is impacted by a multiplicity of factors associated with individual, population and environmental characteristics. As exposure and transmission aren’t directly observed, evaluating the contribution of these different factors to epidemic dynamics generally proves difficult. In order to anticipate the burden associated with epidemics and guide control policies, it is however pivotal to understand how these different elements shape transmission risk.

Sequence data can provide insights into the proximity of individuals in a transmission chain. Phylogeographic approaches have helped characterise how pathogens spread between different geographical regions [1, 2] and demographic groups [3]. However, these methods currently face multiple limitations. First, they do not scale well past a few hundred or few thousand sequences due to difficulties in scaling phylogenetic tree inference. Second, conclusions can be highly biased when sequencing is uneven [4]. We thus critically need new methods to analyse large pathogen genome datasets, such as those produced during the COVID-19 pandemic, which number in the millions of genomes [5].

As mutations accrue over the course of transmission events, we expect epidemiologically related individuals to be infected by pathogens that are genetically similar. Genetic distance cutoff have been used to distinguish plausibly linked infections from infections resulting from distinct introductions within densely sampled outbreaks such as healthcare facilities or nursing homes [6, 7]. Here, we build from this expectation to characterise transmission patterns at the population level.

We introduce a novel statistical framework describing the relative risk of observing genetically proximal sequences in specific subgroups of the population. Our metric of association accounts for heterogeneity in sequencing effort between sampled locations and does not require building a phylogenetic tree, thus making this approach directly scalable to large pathogen genomic datasets. We use this framework to investigate the spatial and social drivers of severe acute respiratory syndrome coronavirus 2 (SARS-CoV-2) transmission in Washington state (WA) by analysing 114,298 sequences (with associated age and home location information) collected through genomic sentinel surveillance in WA between March 2021 and December 2022.

## Results and Discussion

### Identical sequences are imprinted by SARS-CoV-2 spatial patterns of spread

As mutations accrue over time on pathogen sequences, individuals who are close together within a transmission chain are expected to be infected by genetically proximal viruses (Figure 1A). For example, we expect that 64% of SARS-CoV-2 infected individuals are infected by a virus with the same consensus genome as their infector (Figure 1B). Identical sequences should hence be highly informative about SARS-CoV-2 transmission events as they are preferentially collected from the most epidemiologically linked individuals. Therefore, their geographic clustering should be informative about spatial patterns of transmission. In this work, we leverage the clustering of identical sequences between groups to characterise transmission at the population level (Figure 1A). In WA, we identify 17,231 clusters of identical sequences excluding singletons, corresponding to 59,660 sequences (Figure 1C). In some large clusters of identical sequences, we observe local spread prior to wider geographic expansion (Figure 1D). Using postal codes and collection dates, we estimated cluster radius in km. Across clusters, we find that the spatial expansion of clusters increases over time (Figure 1E) and is significantly lower than expected at random (Figure S1). The probability for a cluster to remain within the county and zip code where it was first identified decreases over time. These probabilities are significantly higher than expected at random (Figure S1). This confirms that clusters of identical sequences contain a strong spatial and temporal signature of spread.

**Figure 1.**
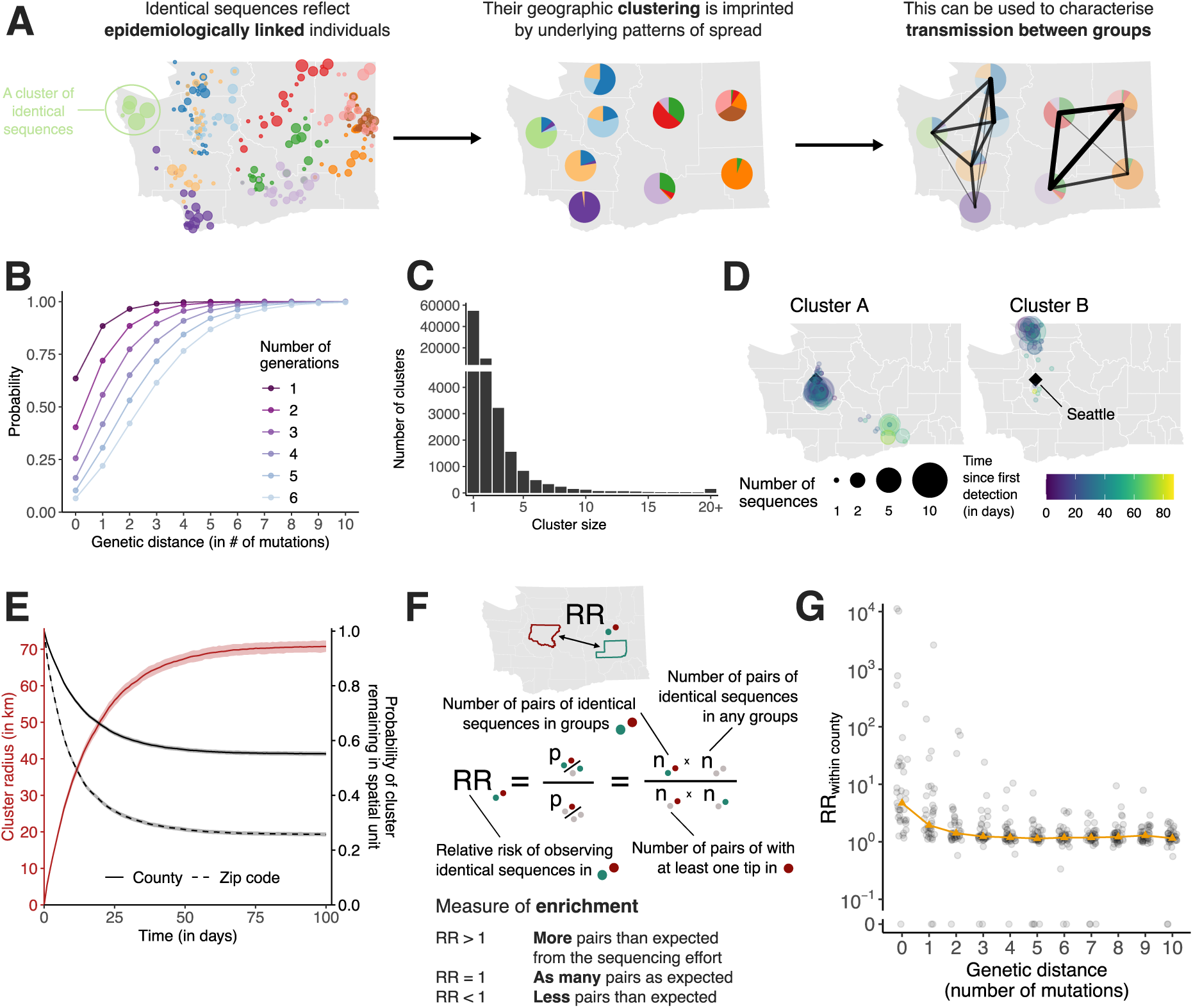
Temporal and spatial signature of spread in clusters of identical SARS-CoV-2 sequences. **A.** The clustering of identical pathogen sequences across population groups reflects underlying disease transmission patterns at the population level and can be use to characterise spread patterns between groups. In this toy figure, each colour represents a different cluster of identical sequences. **B.** Probability for two individuals separated by a fixed number of transmission generations of being infected by viruses at a given genetic distance assuming a Poisson process for the occurrence of substitutions (at a rate *µ* = 8.98 · 10^−2^ substitutions per day) and Gamma distributed generation time (of mean 5.9 days and standard deviation 4.8 days). **C.** Size distribution of clusters of identical sequences in the WA dataset. Clusters of size 1 correspond to singletons and are hence not included in the relative risk computations. **D.** Spatio-temporal dynamics of sequence collection in two large clusters of identical sequences. Black diamonds indicate the location of Seattle, the largest city in WA. **E.** Radius of clusters of identical sequences and probability for all sequences within a cluster of identical sequences of remaining in the same spatial units as a function of time since first sequence collection. In D, the cluster radius is computed as the mean spatial expansion of clusters of identical sequences. **F.** Definition of the relative risk of observing pairs of sequences in two subgroups as a measure of enrichment. **G.** Relative risk of observing pairs of sequences within the same county as a function of the genetic distance separating them. Grey points correspond to values for individual counties. Orange triangles correspond to the median across counties.

### Relative risk framework to look at the distribution of identical sequences

To quantify the association between subgroups of the population (such as geographical units or age groups) from genetic data, we introduce a measure of relative risk (RR) describing how the number of pairs of sequences separated by a fixed genetic distance observed in two subgroups differs from what we expect from the sequencing effort (Figure 1F, see *Methods*). This RR can be interpreted as a measure of enrichment describing how the number of pairs shared by these two subgroups differs from what we expect from the overall number of pairs observed in these two subgroups.

Figure 1G depicts the relationship between the RR of observing sequences within the same county and the genetic distance between pairs. Among all counties, the median RR of observing identical sequences within the same county is equal to 4.7 (interquartile range: 2.4–21.2) across the time period. When considering a greater genetic distance between pairs, this signal decreases to plateau at 1. This confirms that the location of genetically close sequences (less than a couple mutations away) and especially identical sequences is informative about local spread patterns, wherein infected individuals transmit more often within their home county.

We observe this trend across variants and periods (Figure S2). The magnitude of the absolute RR along with the speed at which it decays as a function of genetic distance vary. For example, during the period where the prevalence of Omicron rises and Delta declines, the RR of observing identical sequences in the same county is higher among Delta than Omicron sequences. This can be explained by differences in transmission intensity: a higher transmission rate results in larger clusters of identical sequences [8] that will tend to be more geographically widespread (Figure S2C). The spatial signal from genetically close sequences is hence weaker in periods characterised by a higher transmission intensity. Other factors, such as changes in mixing and travels patterns, can also impact the magnitude of the RR.

Sampling biases can considerably impact the results of phylogeographic inference [4]. Here, although the proportion of pairs of identical sequences observed in a county is highly correlated with the number of sequences observed in this county, we find that the RR is no longer correlated with sequencing effort (Figure S3). Using a simulation approach, we show that our RR metric captures the migration probability between population subgroups, including when sequencing effort is heterogeneous (Figure S4, Table S1). This contrasts with migration rates obtained from a discrete trait analysis (DTA) [9] which are poorly correlated with true migration rates when sequencing effort differs between regions (Figure S4, Table S1). These DTA results are obtained by inputting the exact simulated transmission tree. In practice, inferring the underlying tree will decrease accuracy due to phylogenetic uncertainty so that these DTA estimates represent an upper bound of DTA’s potential performance. If we compare DTA accuracy between input phylogeny and phylogeny estimated from sequence data we find that Pearson correlation between true migration rates and estimated migration rates changes from 0.54 to 0.10 for unbiased sampling and changes from −0.22 to 0.15 for biased sampling (Table S1). Running the phylogenetic DTA analysis on simulated data with 1745 sequences requires 1 day when using the empirical tree and 24 days when jointly inferring the tree and the migration rates (see Methods). Running our RR analysis on the same sequence dataset takes 33 seconds. This result demonstrates that the RR framework constitutes an appropriate approach to study the determinants of SARS-CoV-2 transmission by explicitly accounting for sequencing effort and uneven sequencing between population subgroups.

### Patterns of SARS-CoV-2 spread between WA counties

We explore geographic spread through analysing patterns of occurrence of identical sequences in WA counties (Figure 2). The matrix of pairwise RRs between counties (Figure S5) is characterised by a strong diagonal, which is consistent with within-county transmission. To better understand the spatial patterns of SARS-CoV-2 spread between counties, we display these RRs on choropleth maps indicating RR for different focal counties (Figure 2A, Figure S6). These maps suggest that identical sequences have a higher risk of falling within counties that are geographically nearby. Across all pairs of counties, we find a geographic gradient in the RR of identical sequences, where the risk is highest within the same county, intermediate between adjacent counties, and lowest between non-adjacent counties (Figure 2B). The risk of observing identical sequences between counties also decays as a function of geographic distance (Figure 2C) and is no longer significant at distances greater than 177 km (95% confidence interval (CI): 137-241).

**Figure 2.**
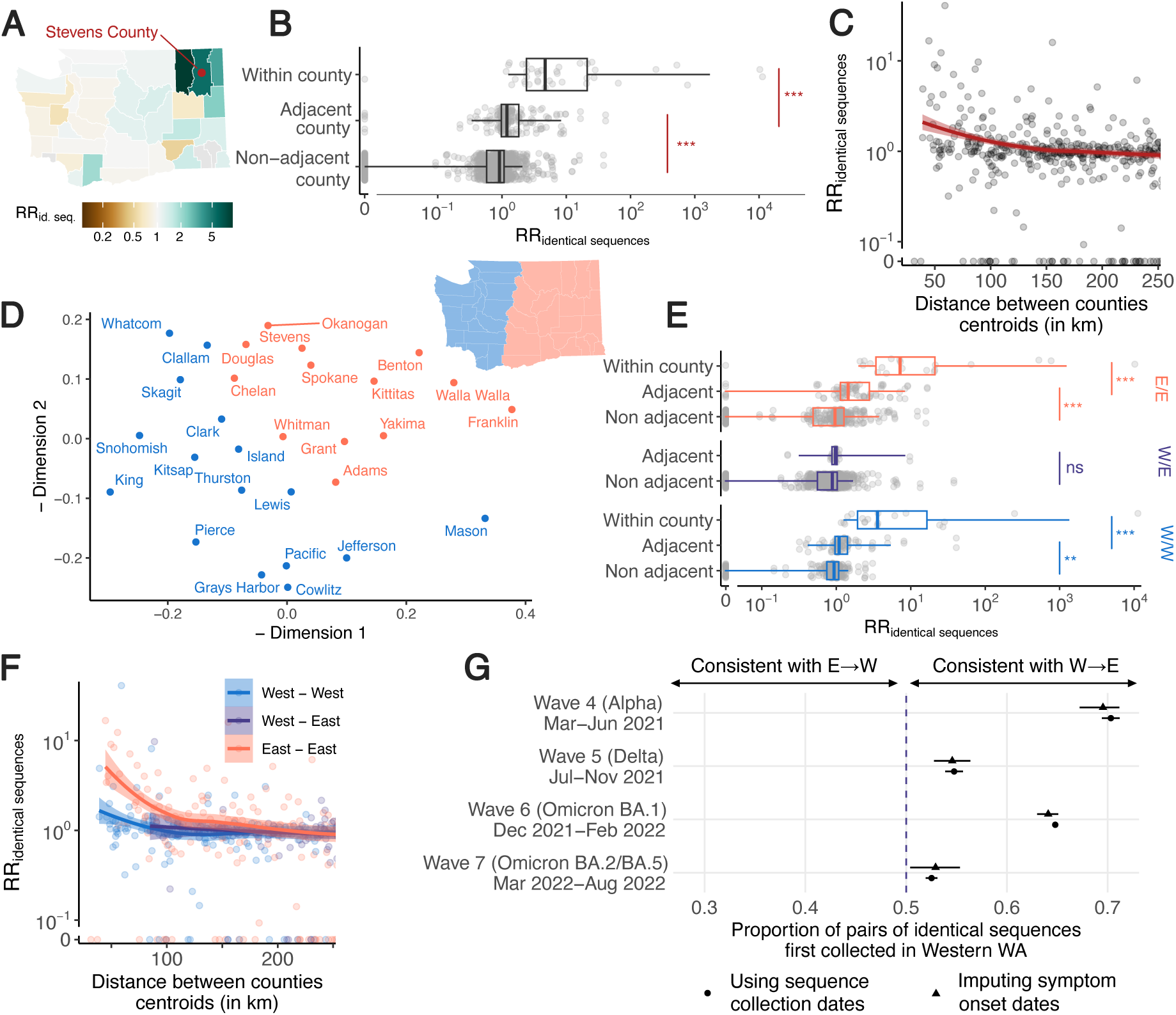
Identical sequences reveal patterns of spread between WA counties. **A.** Illustration of the pairwise relative risk of observing identical sequences between counties, using sequences shared between Stevens County (red point) and other counties in WA as an example. Similar maps for the other counties are depicted in Figure S6. **B.** Relative risk of observing pairs of identical sequences by counties’ adjacency status. **C.** Relative risk of observing pairs of identical sequences as a function of the geographic distance between counties’ centroids. **D.** Similarity between WA counties obtained from MDS based on the relative risk of observing pairs of identical sequences in two counties. Counties are colored by East / West region membership. **E.** Relative risk of observing pairs of identical sequences by counties’ adjacency status stratified by counties East / West region membership. **F.** Relative risk of observing pairs of identical sequences as a function of the geographic distance between counties’ centroids stratified by counties East / West region membership. **G.** Proportion of pairs of identical sequences observed in Eastern and Western WA that were first observed in Western WA. In C and F, the lines correspond to LOESS curves on the logarithmic scale. In B and E, p-values for Wilcoxon tests: *** *<* 0.0001, ** *<* 0.001, * *<* 0.05, ns ≥ 0.05. In B, *W*_within, adjacent_ = 6195 (*p* = 3.7 · 10^−12^) and *W*_adjacent, non adjacent_ = 65542 (*p <* 2.2 · 10^−16^). In E, for within Eastern WA, *W*_within, adjacent_ = 120.5 (*p* = 6.7 · 10^−6^) and *W*_adjacent, non adjacent_ = 4555.5 (*p* = 4.0·10^−6^). For within Western WA, *W*_within, adjacent_ = 95 (*p* = 9.9·10^−7^) and *W*_adjacent, non adjacent_ = 4555.53626 (*p* = 1.1 · 10^−4^). For between Eastern and Western WA, *W* = 2719 (*p* = 0.17).

To assess whether global spatial structure is maintained, we implement a multidimensional scaling (MDS) algorithm by defining a similarity metric based on the RR of observing identical sequences between counties. MDS enables us to display the relatedness of observations based on a distance matrix. This MDS ordination shows country relationships that recapitulate the Western and Eastern WA regions, two regions separated by the Cascades mountain range (Figure 2D). Within Eastern and Western WA, we find a strong signal for local spread, with identical sequences having a higher risk of being observed between adjacent than between non-adjacent counties (Figure 2E). Across the Eastern / Western WA border, we no longer find that identical sequences have an increased risk of being observed in adjacent counties. Results are similar when analysing pairs of identical sequences at the postal code level (Table S2). This lack of association is not affected by the low number of pairs of adjacent counties across the Eastern / Western WA border (Figure S7). This illustrates how heterogeneous physical landscape features can impact and distort patterns of disease spread and genetic diversity [10–13]. We also find that the association between the RR of observing identical sequences in two counties is significant at greater distance within Eastern WA than within Western WA (Figure 2F). We do not find any association with distance across the W/E border, though this might be explained by the lack of counties with low distances across the Eastern / Western WA border.

Finally, we find that, across epidemic waves, pairs of identical sequences observed on both sides of the Cascades are consistently observed first in Western WA (Figure 2G, Figure S8). As testing behaviour and access to healthcare are likely influenced by county demographic characteristics and how rural or urban a county is, we explore how this trend varied when using symptom onset dates instead of sequence collection dates, which provides similar trends (Figure 2G). Despite the existence of negative serial interval for SARS-CoV-2 [14], this analysis provides direct insight into the typical transmission direction between groups as the proportion of SARS-CoV-2 transmission pairs with positive serial intervals in greater than 50% (Supplementary text 1) [15]. This asymmetry suggests that identical sequence clusters tend to percolate from Western to Eastern WA more so than the reverse, indicating transmission generally flows from Western to Eastern WA. This trend is similar to the one reported in phylogeographic analyses of the first COVID-19 wave in WA, that concluded that more introductions occurred from Western to Eastern WA than from Eastern to Western WA [16].

### Human mobility predicts the location of pairs of identical sequences

Next, we explore to what extent spatial transmission patterns inferred from identical sequences can be explained by human mobility indicators. To compute the RR of movement between two counties or regions, we use aggregated mobile phone location data obtained from the SafeGraph ‘Weekly Patterns’ dataset and pre-pandemic commuting data from the US Census Bureau [17] (see Methods). Despite commuting data being collected before the pandemic and mobile phone location data being collected during our study period, we find that these two mobility data sources are highly correlated (Figure S9). We assess how the RR of observing identical sequences in two counties relates to the RR of movement (Figure 3A, Figure S10, Table S3) by implementing a generalised additive model (GAM) that includes a single predictor of smoothed RR of movement between two counties as a covariate. We use a GAM rather than linear regression as we expect the functional form of the relationship to be non-linear (Figure S11). This non-linearity can be explained by the indirect mapping between transmission events and identical sequences that encompass both direct transmission pairs and pairs of individuals a couple generations apart.

**Figure 3.**
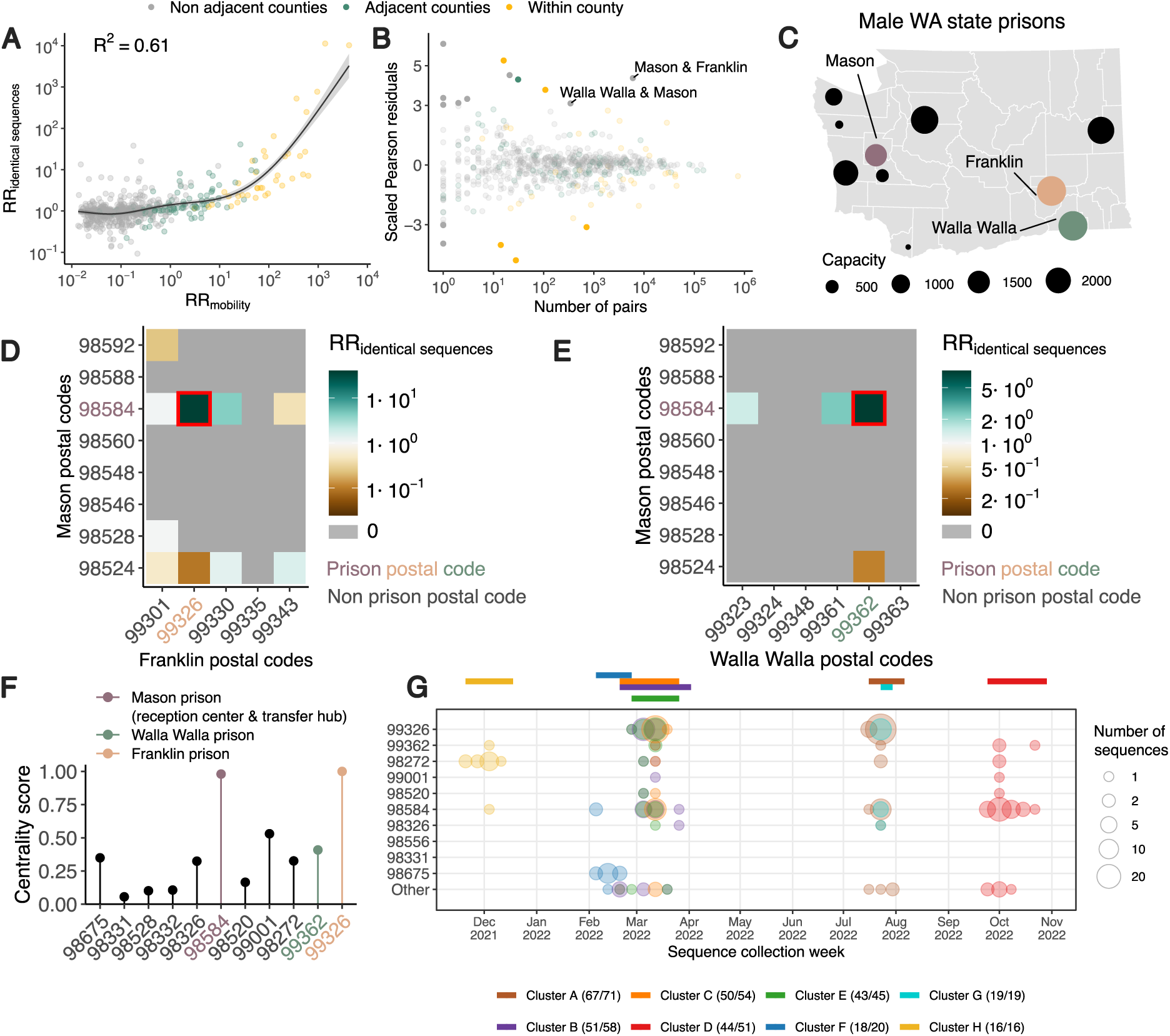
Comparison of the location of identical sequences with expectations from mobility data reveals spread between WA male prisons’ postal codes. **A.** Relationship between the relative risk of observing identical sequences in two counties and the relative risk of movement between these counties as obtained from mobile phone mobility data. The trend line corresponds to predicted relative risk of observing identical sequences in two regions from a GAM. *R*^2^ indicates the variance explained by the GAM. **B.** Scaled Pearson residuals of the GAM plotted in A as a function of the number of pairs of identical sequences observed in pairs of counties. **C.** Map of male state prisons in WA. Mason, Walla Walla and Franklin male prisons are colored. **D.** Relative risk of observing identical sequence between Mason and Franklin County’s postal codes. **E.** Relative risk of observing identical sequence between Mason and Walla Walla County’s postal codes. **F.** Centrality score (eigenvector centrality) for each postal code that is the home of a male state prison. **G.** Week of sequence collection of 8 large clusters of identical sequences identified in postal codes with WA male state prisons. In G, the top colored segments indicate the period during which each cluster was identified.

When comparing RRs at the county level, we find that 60% of the variance in identical sequence data is explained by between-county flows derived from the mobile phone data (Figure 3A, Table S3). For a subset of counties, the number of pairs of identical sequences or the number of trips reported in the mobility dataset are low. For these low counts, we expect RRs to be more noisy. To remove potential noise associated with these lower counts, we repeat this analysis at a larger spatial scale. Aggregating pairs at the regional level (9 regions in WA for 39 counties, Figure S12) increases the variance explained to 81% (Figure S13). We also find that pre-pandemic workflow data are highly informative of the spatial distribution of pairs of identical sequences with a similar strength of relationship as observed for mobile phone mobility data (Figure S10, S13, Table S3).

Non-pharmaceutical interventions along with behavioral changes have impacted human mobility patterns throughout the COVID-19 pandemic. We find that mobility data explains a high percentage of variance in the RR of observing identical sequences between WA regions across individual epidemic waves (Figure S14) but not to a greater extent than over the entire study period. This can likely be explained by the high stability of the structure of the mobility network between WA counties across epidemic waves (Figure S15). This suggests that analyzing COVID-19 waves separately tends to introduce noise rather than increase spatial resolution, in line with a former analysis concluding to the high stability of between-county mobility patterns during the beginning of the pandemic in the United States [18].

Among counties located across the Eastern / Western WA border, the risk of movement across the border is lower than the risk of movement within the same region (Figure S16). This shows that human mobility is highly predictive of the location of pairs of identical sequences and explains some of the spatial patterns reported in Figure 2.

### Outliers in the relationship between mobility and sequence data appear associated with male state prisons

We identify unexpected patterns of transmission between counties from outliers in the relationship between mobility and genetic data (Figure 3A). We define outliers as pairs of counties for which the absolute value of the scaled Pearson residuals from the GAM are greater than 3. As we expect RRs computed from a low number of pairs of identical sequences to be noisier, we focus on pairs of counties between which at least 100 pairs of identical sequences are observed. We find unexpected patterns of SARS-CoV-2 spread between two non-adjacent pairs of counties, Franklin/Mason and Walla Walla/Mason Counties, with more pairs of identical sequences observed than expected from mobility data (Figure 3B). The association between Franklin & Mason (RR of 13.4 (95% CI: 11.4-16.4)) and Walla Walla & Mason (RR of 5.9 (95% CI: 4.0-8.3)) is particularly surprising given that they are non-adjacent counties located on different sides of the Cascades. As no demographic or geographic factors provide a straightforward explanation for such an association, we hypothesise that such a pattern might arise from SARS-CoV-2 spread on a dissemination network that differs from the general community. We identify that these three counties are the home of male state correction centers (Figure 3C). We also find that identical sequences have a higher risk of being observed within Lincoln County and a lower risk of being observed within Pacific County that expected from cellphone mobility data, without identifying any demographic factor explaining these associations.

To investigate whether the unexpected pattern of association between Franklin & Mason and Walla Walla & Mason Counties can be explained by transmission within the prison network, we look at patterns of association between Franklin & Mason and Walla Walla & Mason postal codes (Figure 3D-E). For most of these pairs of postal codes, we don’t observe any pair of identical sequences throughout the study period. Interestingly, for each pair of counties, the genetic signal can be explained by a high RR of observing identical sequences between two postal codes, which correspond to the postal codes that are the home of the male correction centers we identified. The greater number of pairs of identical sequences observed between Mason & Franklin and Mason & Walla Walla counties than expected from mobile phone derived mobility data can hence be explained by a large number of pairs of identical sequences in specific postal codes with male correction centers.

We also investigate patterns of occurrence of pairs of identical sequences between the two counties (Mason and Pierce) that are the home of female prisons. At the county level, identical sequences don’t have an increased risk of occurring between Mason & Pierce counties (RR of 0.59 (95% CI: 0.49-0.67)). At the postal code level however, we find that the RR of observing identical sequences is highest between the two postal codes with female prisons (Figure S17). This shows how our framework enables exploration of patterns of spread at different spatial scales: we don’t find any signal at the county level, likely because Mason and Pierce are adjacent counties, but we can identify association at the postal code level.

It is interesting that the pairs of outliers we identified systematically involved Mason County (Figure 3B), which is the home of only the sixth (out of ten) most populated male prison in the state (Figure 3C). The prison in Mason County (Washington Corrections Center) plays a particular role in the WA prison network since it serves both as a reception center for anyone entering the WA prison system and as a transfer hub [19]. To understand whether the prison network structure can explain patterns of SARS-CoV-2 transmission, we conduct a centrality analysis. To do so, we analyse the network of postal codes with WA male prisons and we define the weight of each edge by the RR of observing identical sequences between these two postal codes. We find that the two nodes with the highest eigenvector centrality scores are the postal codes that are the home of Washington Corrections Center (Figure 3F) and of the Franklin County prison (most populated prison). This shows that patterns of occurrence of identical sequences in WA are imprinted by the structure of the prison network.

Finally, we investigate whether large clusters of identical sequences are shared between postal codes with male state prisons, which we define as clusters with more than 15 sequences in male state prisons postal codes. Figure 3G depicts the timing of the large clusters we identify. Notably, the largest cluster (Cluster A) includes 71 sequences collected between 18 July and 31 July 2022, 67 of which came from postal codes with male state prisons. The second largest cluster (Cluster B) is composed of 58 sequences collected between 21 February and 29 March 2023, among which 51 came from 7 different prison postal codes. Interestingly, the postal code of Washington Corrections Center is the only one in which all these eight clusters were observed.

Populations who are incarcerated have been particularly affected by the COVID-19 pandemic [20, 21]. To mitigate the impact of the pandemic in these congregate settings, various interventions have been implemented. In WA, for example, testing followed by quarantine protocols were carried out in Washington Correction Center upon admission and before any transfer. Active screening of staff was also implemented throughout the pandemic. Individuals incarcerated diagnosed with COVID-19 however at times had to be transferred from Washington Correction Center to other WA prisons due to the finite capacity of the reception center. With vaccine mandates, staff also had to be relocated to cope with the departure of other employees. Our results reveal multiple SARS-CoV-2 introductions between WA prisons, that could be explained by the movements of both individuals incarcerated and staff.

This analysis showcases how identical sequences can help identify under recognised viral dissemination networks that differ from transmission pathways in the general community. The counties we identified as outliers in the relationship between genetic and mobility data have a particularly high ratio between the prison population size and the county population size (between around 2% and 4%, Table S4). This likely explains why we were able to detect this signal at the county level but had to investigate transmission at the postal code level to study transmission between other prisons.

### The spatial scale of spread impacts transmission patterns between age groups

Spatial and social factors (such as age) are key determinants of the spread of respiratory infections such as SARS-CoV-2 and influenza [22–25]. We expect movement patterns to differ between age groups (such as children, adults and elderly people), which can impact patterns of disease transmission [26–28]. There has however been limited empirical evidence of this phenomenon and data sources that can be leveraged to characterise this interaction are critically needed. Here, we show that we can combine pathogen sequence information with detailed metadata to investigate how age mixing patterns vary across spatial scales.

We first examine whether we can recover the expected age mixing signature from the sequence data before delving into the interaction between age and space. We find that the age groups in which identical sequences are observed are consistent with assortative mixing patterns and mixing between generations (Figure S18). Comparing this with expectations from synthetic social contact data for WA [29], we find that the signal obtained from identical sequences are highly correlated with that expected from age mixing matrices (Figure 4A) (GAM: 90% of variance explained; Spearman *ρ* = 0.86, *p <* 10^−16^). The signal for SARS-CoV-2 transmission between generations (such as the 0-9y and the 30-39y) fades out when considering pairs of sequences separated by a greater genetic distance (Figure S19). As sequences at a greater genetic distance come from individuals who are further apart within a transmission chain (Figure 1A), fine-scale patterns of spread might indeed not be apparent from sequences at more than a couple mutations away. This emphasises the value of analysing identical pathogen sequences to characterise subtle patterns of pathogen spread and population mixing, especially when population subgroups are very mixed.

**Figure 4.**
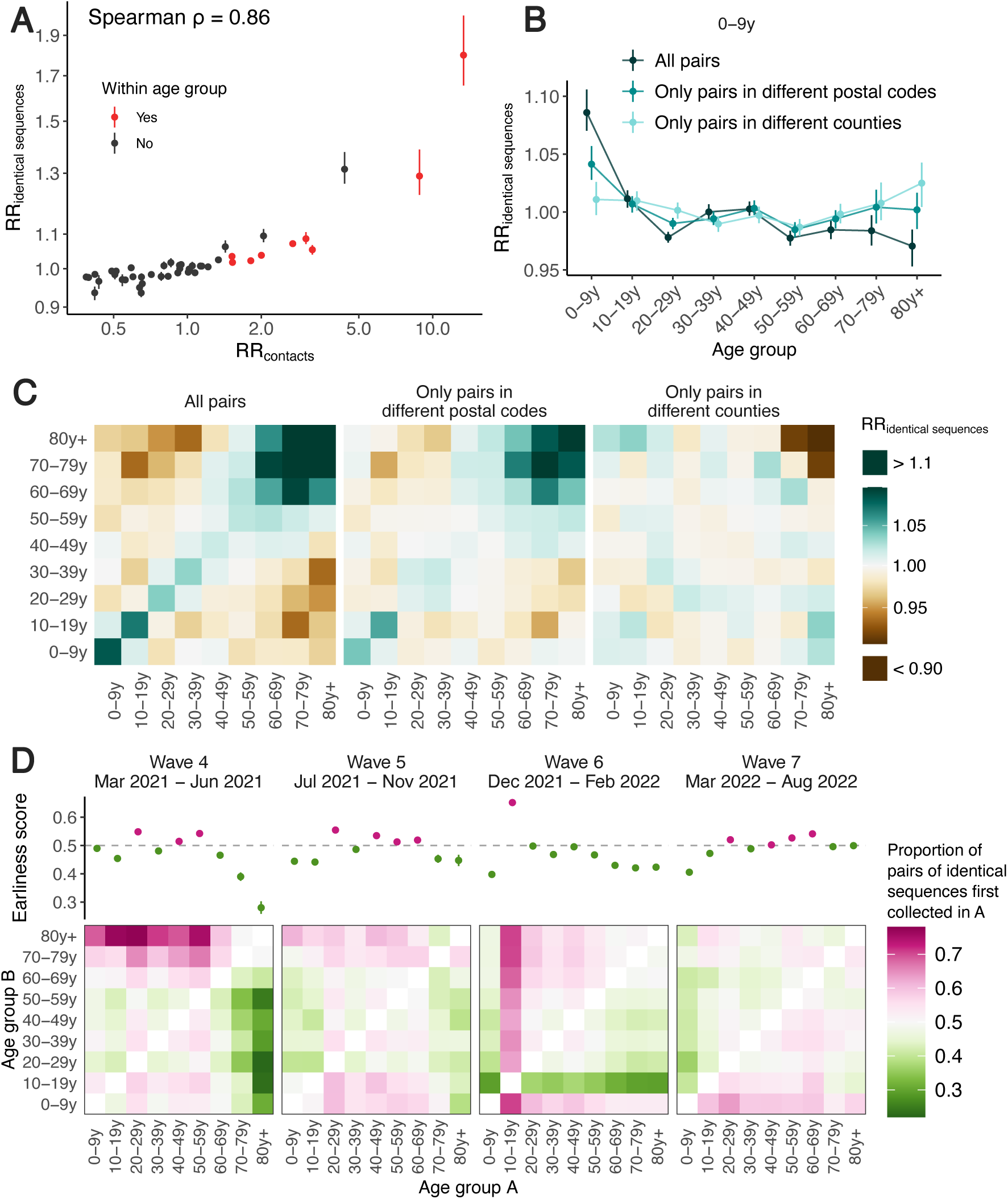
Patterns of SARS-CoV-2 transmission between age groups in WA. **A.** Relative risk of observing pairs of identical sequences in two age groups as a function of the relative risk of contact between these age groups. **B.** Impact of the spatial scale on the relative risk of observing pairs of identical sequences in the 0-9 y.o. and other age groups. We display similar plots for the other age groups in Figure S20. **C.** Relative risk of observing identical sequences between two age groups across all pairs of sequences, only pairs in different postal codes and only pairs in different counties. **D.** Proportion of pairs of identical sequences observed in age groups *A* and *B* that were first collected in age group *A* across different epidemic waves (heatmaps). The dot plots depict the earliness scores of age group *A* across epidemic waves. In A and B, vertical segments correspond to 95% subsampling confidence intervals. In D, vertical segments correspond to 95% binomial confidence intervals. In D, the heatmaps represent symmetric matrices *P* = (*p_i,j_*) characterised by *p_i,j_* + *p_j,i_* = 1.

Next, we compare the RR of observing identical sequences between two age groups by looking at either all pairs of sequences or only pairs of sequences from individuals living in different counties or postal codes. We find that the spatial scale modulates patterns of disease transmission between age groups (Figure 4B-C, S20). We find that pairs of identical sequences coming from the same county and postal code are enriched in same-age pairs. This enrichment is particularly important in elderly groups. For example, the RR of observing identical sequences in individuals aged 80 and older drops from 1.80 (95% CI: 1.65-2.01) when considering all pairs to 0.79 (95% CI: 0.71-0.85) when considering pairs coming from individuals living in different counties (Figure S20). This shows that transmission to and from older age groups tends to occur close to their home location and suggests that elderly individuals’ typical radius of movement is smaller than that of other age groups. Only considering pairs of sequences in 0-9 year-olds coming from different spatial units largely decreases the signal for SARS-CoV-2 transmission between children and adults aged 30-49y (Figure 4C). This is expected given that we anticipate most of these contacts to occur within the household [30]. Overall, we find that looking at patterns of occurrence of identical sequences at a greater geographic scale largely distorts the contact structure. For example, the location of identical sequences suggest that transmission to and from elderly individuals outside of their home counties tend to occur with younger age groups, including younger children (e.g. grandchildren).

Mixing patterns between age groups have been extensively studied [31, 32]. A social contact survey performed in southern China reported that elderly individuals’ contacts occurred closer to their homes than younger individuals’ contacts [33]. Overall, there has however been limited evidence to quantify the spatial distribution of these contacts. Spatial mixing is generally measured from aggregated mobility data sources which generally do not provide demographic information such as age. Because spatial and age mixing are reconstructed from different data sources, understanding their interplay has been difficult. Here, we show that we can directly leverage pathogen genome data with linked age and spatial information to understand where age-specific transmission is occurring. This suggests that the wider availability of sequencing data might provide an opportunity to directly infer how population groups interact in a way that is relevant for pathogen spread, without the need to implement laborious contact surveys or collect mobility data.

### The timing of identical sequence collection sheds light on the groups driving transmission

Finally, we use the timing of identical sequence collection to investigate the age groups driving SARS-CoV-2 transmission over the course of the pandemic in WA. Within pairs of identical sequences, we indeed expect age groups acting as sources to be consistently detected before groups acting as sinks (Figure S21). In Figure 4D, we display for every age group combination and across epidemic waves the proportion of pairs of identical sequences first collected in a given age group. During the fourth and fifth pandemic wave in WA (respectively mainly caused by the Alpha and Delta SARS-CoV-2 variants of concern), we find that pairs of identical sequences are consistently observed later in elderly groups even though the RR of observing identical sequences in elderly groups and younger groups is low (Figure S18). This could be consistent with younger age groups acting as source of infections for elderly individuals. During the fourth pandemic wave, sequences from individuals aged 20-29y and 40-59y are systematically observed before any other groups within pairs of identical sequences and likewise during the fifth pandemic wave, sequences from 20-29y and 40-69y individuals are observed earlier than other age groups. This could be consistent with these groups acting as sources of infection for the other age groups. During the sixth wave, sequences from 10-19y tend to be observed first within pairs of identical sequences, which suggests their role as sources for other ages groups and corresponds to the Omicron wave during a time when schools had recently returned to in-person instruction. From March 2022, the contribution to transmission is more evenly distributed across age groups.

The role played by young children during the COVID-19 pandemic has been highly debated [25,34]. Here, we find that during the Alpha and Delta epidemics (waves 4 and 5), children aged 0-9y could have acted as source of SARS-CoV-2 infections for elderly individuals but not for younger adults. This pattern disappears during the Omicron epidemic (wave 6) in which pairs of identical sequences tend to be observed first in other age groups before being collected in young children aged 0-9y. This could be explained by behavioral changes or by different immune profiles across age groups, resulting in different relative susceptibility to Omicron relative to Delta [35]. Overall, we do not find indication for young school age children to act as major sources of SARS-CoV-2 transmission in the population, even after schools reopened.

Overall, our results highlight the porosity of SARS-CoV-2 transmission across age groups and suggest a role played by lower risk groups in seeding infections in higher risk groups. We come to similar conclusions when looking at the timing of symptom onset dates (Figure S22), which suggests that our conclusions are robust to differences in testing behaviors across age groups. Our conclusions are in line with existing literature emphasising the important role played by young adults and teenagers and the limited contribution of children and elderly individuals in driving SARS-CoV-2 spread [25, 36]. Analysing the timing of identical sequence collection provides immediate insights into pathogen flow between population subgroups.

Here, we focus on understanding patterns of SARS-CoV-2 spread between age groups but this approach can be applied to investigate the spread of fast-evolving pathogens between various demographic groups, such as occupational and ethnic groups, behavioral and risk groups. For example, we find that identical sequences collected within an age group tend to be enriched in same-vaccine status pairs during the Alpha, Delta and Omicron BA.2/BA.5 waves in WA (Figure S23). Social clustering of unvaccinated individuals or generally individuals with different immune background can have important implications regarding the size and likelihood of infectious disease outbreaks [37, 38]. This suggests that our approach has potential to shed light on such a phenomenon and more generally on broad determinants of disease transmission

### Caveats

Though our RR metric explicitly accounts for sampling intensity in locations in which pairs of identical sequences are collected, it cannot describe patterns of spread from non-sampled locations. In theory, unsampled locations could impact our assessment of local transmission patterns if two individuals with an identical sequence are both infected by someone outside the study area. In practice however, we find that non-sampled locations have little impact our RR computation (Figure S24). This suggests that background sequences are not required to evaluate SARS-CoV-2 transmission at the state level. Other epidemiological settings might nonetheless require more careful considerations, for example in the hypothetical scenario where a non-sampled location is responsible for the majority of the infections within the study area.

Compared with existing phylogeographic methods, our approach however does not require including background sequences from outside locations as non-sampled locations little impact the RR computation (Figure S24). Our approach could also overestimate RRs associated with transmission events that are over-represented in the sequencing data. For example, applying this analysis to sequences predominantly collected through household studies could overestimate the contribution of contacts within the household to the overall infection burden. In our case, patterns of occurrence of identical sequences in WA are potentially affected by intensive testing performed during outbreaks within WA prisons during the pandemic. Part of the signal we have detected might hence come from a higher sequencing rate in prisons compared with the general community. The very large clusters of identical sequences shared between multiple prison postal codes however confirm that SARS-CoV-2 extensively spread within the prison network.

Our results suggest that the timing of sequence collection within clusters of identical sequences provides valuable information to understand transmission direction. Here, we report a simple quantity based on the proportion of pairs first observed in a group, that summarises the earliness of a group compared to another within pairs of identical sequences. Such an earliness metric might incompletely capture the transmission process as clusters can for example span more than 2 groups. We perform a sensitivity analysis where we only rely on clusters of identical sequences observed in two groups, that provides similar results (Figure S25). Overall, more work is warranted to robustly quantify transmission rates from the timing of identical sequences.

Finally, the magnitude of RRs is impacted by transmission intensity (Figure S2), so that values computed between two regions across different time periods cannot be directly compared. Within a time period, the ranking of RRs is however informative about patterns of transmission between groups. Overall, further work exploring how patterns of occurrence of identical sequences can be used to directly infer mixing rates between groups while incorporating temporal changes in transmissibility would be particularly interesting.

### Applicability beyond SARS-CoV-2

In this manuscript, we study patterns of SARS-CoV-2 transmission between geographies and age groups in WA using a particularly rich sequence dataset, both given the amount of sequences available and the quality of the associated metadata. This work can however readily be applied to other densely sampled pathogens.

The power of our method is determined by the number of pairs of identical sequences available which will be impacted by transmission intensity (higher reproduction numbers will tend to result in larger clusters of identical sequences [8]) and the relative timescale at which substitution and transmission events occur [8]. Overall, this approach is well tailored to study densely sampled outbreaks. Compared with phylogeny-based methods whose power comes from the number of unique haplotypes, our ability to characterise spread from identical sequences depends on the number of haplotypes with multiple observations. Whereas we expect diminishing returns of sequencing a greater proportion of cases for phylogeny-based methods with a decreasing number new haplotypes per additional sequence, the number of haplotypes with multiple occurrences will increase for each additional sequence included in the dataset (Figure S26). The number of population groups included in the analysis will also impact the amount of sequencing data required. In situations where the sample size results in a lower number of pairs of identical sequences, aggregating groups can be a valuable strategy to reduce uncertainty. Within our WA sequencing dataset, we find that assessing spread between 2 age groups requires around 10^2^ −10^3^ sequences whereas 9 age groups increases the number of sequences required to 10^4^ − 10^5^ (Figure S27).

Extending the analysis to pairs of sequences separated by a greater Hamming distance could also increase the statistical power by increasing the amount of data available, in particular for pathogens characterised by a higher mutation rate (Figure S28). Increasing the threshold however comes at the cost of diluting the signal by including pairs that are less epidemiologically linked in the analysis, hence introducing more noise (Figure S28). Other factors, such as the rate of mixing between groups or the natural history of the infection, can impact the optimal threshold.

### Perspectives

Large scale pathogen genome sequencing provides an incredible opportunity to understand where and how transmission is occurring. The computational cost of existing methods that rely on inferring the phylogenetic tree has limited their ability to elucidate fine-scale transmission patterns. Here, we show that a simple count-based metric based on pairs of identical pathogen sequences with detailed linked metadata can provide unique insights into the determinants of SARS-CoV-2 transmission. Future work investigating how to better describe asymmetry in transmission between groups and how to infer group-level contributions to epidemic growth from such data are a promising research direction. This shows that relying on pairs of identical or nearly identical pathogen sequences along with fine-grained metadata is valuable to understand how and where transmission is happening. By providing scalable new tools to understand detailed pathogen spread patterns, we believe that this work represents an important development to guide future epidemic control efforts.

## Methods

### Data sources and preprocessing

#### Sequence data and metadata

We analyse 116,791 SARS-CoV-2 sequences from Washington state genomic sentinel surveillance system [39] sampled between 1 March 2021 and 31 December 2022. Sequence metadata are collated by the Washington State Department of Health and include sample collection date, symptom onset date, de-identified patient ID, county of home location, postal code of home location, age group (0-9y, 10-19y, 20-29y, 30-39y, 40-49y, 50-59y, 60-69y, 70-79y and 80y+) and vaccination status upon positive test. For patients with multiple sequences in the database (2,309 out of 114,306 patients), we restrict our analysis to the earliest sequence collected. Among these 114,306 sequences, the age information is missing for 1 sequence, the county information for 659 sequences and the postal code information for 1011 sequences.

Consensus sequences are extracted from the GISAID EpiCoV database [40, 41] and curated using the Nextstrain nCoV ingest pipeline [42]. We discard sequences with undefined Nexstrain clade assignments (8 sequences out of 114,306). This leaves us with 114,298 sequences, with 114,297 sequences gathered from patients with known age, 113,639 sequences gathered from patients with known county of home location and 113,287 sequences gathered from patients with known postal code of home location. 96 % of sequences have coverage in greater than 90% of the genome. We match postal codes to zip code tabulation areas (ZCTAs). For postal codes that do not have a ZCTA with the same name, we manually match them by looking at ZCTA boundaries. All analyses at the postal code level use ZCTA metadata information. We extract the postal codes of WA prison facilities from [43].

#### Computing pairwise genetic distances from sequences

We compute pairwise genetic distances between Washington state sequences with the *ape* R package [44] using Hamming distances. To avoid unnecessary computational costs, we only compare sequences belonging to the same Nextstrain clade [45] and generate one distance matrix per clade. We don’t expect the clade definition to impact the pairs of identical sequences we identified since pairs of identical sequences should always belong to the same clade.

Generating pairs of identical sequences from a sequence data file is the most computationally expensive step in this analysis. To provide context, generating a distance matrix from 1,000 sequences takes 33 seconds, while 10,000 sequences takes 1 hour 37 minutes, on 1 core of an Apple M2 chip. Generating the full distance matrix for the analysis set of 113,287 sequences took around 96 hours of compute time readily parallelised across a compute cluster. More efficient software tools can significantly bring that compute time down (e.g. 1.14 hour with pairsnp [46]).

#### Workflow data

We use data describing the daily number of commuters between each WA county from the American Community Survey (2016-2020) [17]. This dataset provides the number of directed commuting flows between residence and workplace counties. We use the number of commuting flows between counties to compute the RR of commute between two regions (see below).

#### Mobile phone location data

We obtain mobile device location data from SafeGraph (https://safegraph.com/), a data company that aggregates anonymised location data from 40 million devices, or approximately 10% of the United States population, to measure foot traffic to over 6 million physical places (points of interest) in the US. Following Perofsky et al. [47], we estimate movement within and between counties in Washington from January 2020 to June 2022, using SafeGraph’s *Weekly Patterns* dataset, which provides weekly counts of the total number of unique devices visiting a point of interest (POI) from a particular home census block group. POIs are fixed locations, such as businesses or attractions. A *visit* indicates that a device entered the building or spatial perimeter designated as a POI. The *home location* of a device is defined as its common nighttime (18:00-7:00) census block group (CBG) for the past 6 consecutive weeks. We restrict our dataset to POIs that have been tracked by SafeGraph since December 2019. To measure movement within and between counties, we extract the home CBG of devices visiting POIs in each week and limit the dataset to devices with home CBGs in the county of a given POI (within-county movement) or with home CBGs in counties outside of a given POI’s county (between-county movement). To adjust for variation in SafeGraph’s device panel size over time, we divide Washington’s census population size by the number of devices in SafeGraph’s panel with home locations in Washington state each month and multiply the number of weekly visitors by that value. For each mobility indicator, we sum adjusted weekly visits across POIs from March 2021 to June 2022. We use the number of visits between counties to compute the RR of movement from mobile phone data between two regions (see below).

To explore potential geographic biases in the mobility data, we divided the weekly number of devices residing in each county by the weekly number of devices residing in WA state (“observed proportions”) and compared these values to “expected proportions” based on county and state census population sizes during 2020-2022. SafeGraph’s panel consistently captured 2-5% of each county’s population, with strong correlations between device counts and census population sizes (Spearman’s *ρ* = 0.99, Figure S29). We estimated county-level bias as the “observed proportion” of devices tracked by SafeGraph in individual counties relative to WA state minus the “expected proportion” based on census population sizes. Annual bias estimates for individual counties ranged from −2.2% to 1.7%, with no clear trend of over- or underrepresentation by population size or urban-rural classification (Figure S30). Although the most populous counties in WA state tend to have greater absolute bias, large counties are both underrepresented and overrepresented in the SafeGraph dataset (Figure S30). For example, the most populous county in WA, King County, was slightly underrepresented each year (−2.2% to −1.6% bias; green negative outlier in Figure S30), while three of the other top five largest counties (Clark, Pierce, and Spokane) were slightly overrepresented (1.1% to 1.7% bias; pink, blue, and goldenrod positive outliers in Figure S30). Our method for estimating geographic bias is based on SafeGraph’s Google Co-Lab Notebook on Quantifying Bias [48].

#### Social contact data

We use synthetic social contact data for WA generated by Mistry et al. [29] based on reconstructing synthetic populations of interacting individuals using WA population demographics. They describe the per-capita probability for an individual of age *i* of interacting with someone of age *j* during a day.

### Quantifying connectivity between different population groups

#### From genetic data: Relative risk for sequences separated by a given genetic distance of being in given subgroups of the population

Let *n* denote the number of sequences included in the study and *H_i,j_* the Hamming distance between sequence index by *i* and *j*. Let *S_i_* denote the subgroup of sequence *i*. We introduce 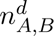 the number of Hamming distance matrix elements (excluding the diagonal) equal to *d* where sequence *i* belongs to group *A* and sequence *j* belongs to group *B*.

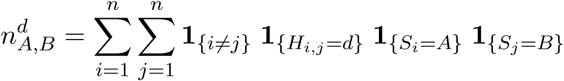

where *X* → **1***_X_* is the indicator function which is equal to 1 if *X* is true and 0 otherwise.

Let 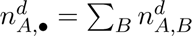 and 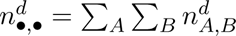.

We derive the relative risk 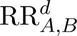 for sequences separated by a genetic distance *d* of being observed in subgroups *A* and *B* compared to what is expected from the sequencing effort in the different subgroups of the population as:

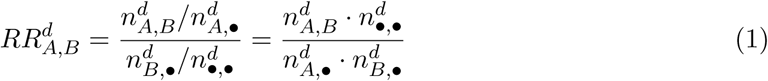

The numerator 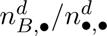 corresponds to the proportion of the pairs where the subgroup of sequence *i* is *A* that are occurring with the *B* group.

The denominator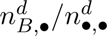 is a normalisation factor quantifying the contribution of group *B* to the total number of pairs separated by a Hamming distance of *d*. The ratio between these two quantities hence quantifies to which extent pairs of sequences observed in groups *A* and *B* are enriched compared to the number of sequences observed in these groups.

We use a subsampling strategy to compute confidence intervals around these RRs. Bootstrapping (random sampling with replacement) would result in comparing sequences with themselves and therefore lead to biased upwards RRs of observing identical sequences in the same group. To avoid this, we used a subsampling strategy (random sampling without replacement) with a 80% subsampling rate (1,000 replicate subsamples).

We provide the tools to compute this RR metric from user-provided sequence and metadata files in the GitHub repository associated with this manuscript [49].

#### From mobility data: Relative risk of movement between two geographical locations

Both the mobile phone and commuting mobility data provide directed flows between WA counties. Let *w_A_*_→*B*_ denote the number of commuters reported in the commuting data (respectively the number of visits for the mobile phone mobility data) whose home residence is in county *A* and who work in county *B* (respectively for which a visit in county *B* is reported). We compute the total movement flow between counties *A* and *B* as :

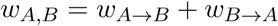

We then calculate the relative risk 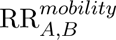 of movement between counties *A* and *B* as:

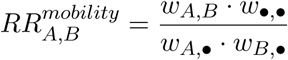

where 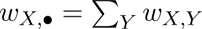 and 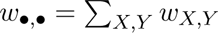.

We compute a similar statistic by aggregating counties at the regional level (Figure S12).

#### From social contact data: Relative risk of contact between two age groups

Mistry et al. estimated the average daily number of contacts *M_i,j_* that individuals of age *i* have with individuals of age *j* (considering one-year age bins) [29]. As we are interested in the age groups available in the sequence metadata, we reconstruct the average daily number of contacts *c_A,B_* that individuals within age group *A* have with individuals in age group *B* as:

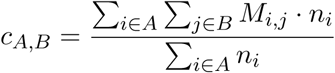

where *n_i_*is the number of individuals of age *i*. We can then derive the total daily number of contacts between age groups *A* and *B* as Γ*_A,B_* = *c_A,B_* ·*N_A_* where *N_A_* is the number of individuals in age group *A*. We then compute the relative risk 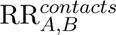 for a contact of occurring between age groups *A* and *B* compared to what we expect if contacts were occurring at random in the population as:

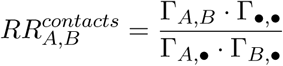

where Γ*_A,_*_•_ is the total daily number of contacts involving individuals within age group *A* and Γ_•_,_•_ is the total daily number of contacts in the population.

### Using the timing of sequences to understand directionality in transmission

#### From sequence collection dates

We introduce *t_x_* as the time at which the sequence *x* was collected. Let *I_A,B_* denote the ensemble of pairs of identical sequences observed in groups *A* and *B*.

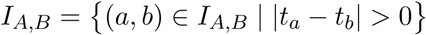

thus denotes the subset of these pairs with different sequence collection dates. We compute the proportion *p_A_*_→*B*_ of pairs consistent with the transmission direction *A* → *B* as:

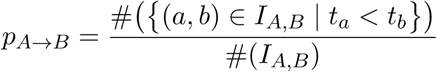

where #(*X*) is the cardinal of *X*.

We also report 95% binomial confidence intervals around these proportions.

#### From symptom onset dates

The delay between infection and sequence collection can be impacted by healthcare seeking behaviours and access to testing which might vary across age groups, geographical locations and time periods. If the distribution of the delay until testing differs between two subgroups *A* and *B*, the proportion of pairs of identical sequences *p_A_*_→*B*_ that are first collected in group *A* will both reflect the timing of infections and healthcare seeking behaviours. When available, symptom onset dates should be less impacted by healthcare seeking behaviours.

Among the 113,638 SARS-CoV-2 sequences with associated age group and county of home location information, symptom onset dates are available for 34,167 of them (30%). The availability of symptom onset information is susceptible to be impacted by individual demographic profiles (such as age), which could result in sequences with symptom onset information not being representative of all the sequences available. To avoid this, we impute missing symptom onset dates based on the empirical delay distribution between symptom onset and sequence collection (computed from individuals with known symptom onset dates) stratified by age group, time period and Eastern/Western WA region (Figure S31). Out of the sequences with known symptom onset dates, the absolute value of the delay between sequence collection and reported symptom onset is strictly greater than 30 days for 192 of them (*<*0.6%). We discarded these sequences in the computation of the symptom onset to sequence collection delay and considered that they were equivalent to sequences with missing symptom onset information (and hence imputed their symptom onset dates). We generate 1,000 imputed datasets. For each of these imputed datasets, we compute the proportion 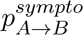 of pairs with symptom onset dates occurring first in group *A* among pairs of identical sequences in groups *A* and *B* with distinct symptom onset dates. We then report the median across these 1,000 imputed datasets. We also generate a measure of uncertainty by computing on each of the imputed datasets 95% binomial confidence intervals around the proportion 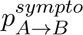. We then report an uncertainty range around these proportion by using the minimum lower bound of the 95% CI and the maximum upper bound of the 95% CI across the imputed datasets.

### Spatial analyses

#### Assessing the spatial extent of clusters of identical sequences

We reconstruct clusters of identical sequences from the pairwise genetic distance matrices [8]. Figure S32 depicts the typical size and duration of these clusters of identical sequences throughout the study period. To assess the spatial and temporal signal in clusters of identical sequences, we evaluate how the spatial extent of a cluster (summarised by its radius) evolves over time. For each cluster, we define primary sequences as the cluster’s earliest collected sequence. We then define the cluster’s primary ZIP Code Tabulation Areas (ZCTAs) as the ZCTAs of its primary sequences. We exclude clusters with ambiguous primary ZCTA (several primary ZCTAs) from this analysis. We define the radius of a cluster at a given time as the maximum distance between the primary ZCTA and the ZCTAs of the sequences collected by that time. We also compute the time required for sequences to be collected outside the primary ZCTA and primary county (using a similar definition as for the primary ZCTA). We report the mean cluster radius and the proportion of clusters remaining within the same geographical unit (ZCTA and county) as a function of the time since collection of the first sequence within a cluster. We generate 95% confidence intervals using a bootstrap approach with 1,000 replicates.

We compare the observed cluster radius and the observed proportion of clusters remaining within the same geographical unit to those expected from a null distribution assuming no spatial dependency between sequences within a cluster of identical sequences. We simulate a null distribution by randomly permuting the geographical locations of the WA sequences and recomputing our statistics of interest (cluster radius, proportion of clusters within the same county and proportion of clusters within the same ZCTA).

#### Comparing the location of pairs identical sequences by counties’ adjacency status

We compare the RR of observing identical sequences between two counties depending on counties’ adjacency status (within the same county, between adjacent counties and between non-adjacent counties) by using Wilcoxon signed rank tests.

#### Assessing the relationship between the RR of observing identical sequences between two counties and the geographic distance separating them

We explore how the RR of observing identical sequences in two distinct counties compare with the geographic distance between counties’ centroids. We summarise this trend by reporting the LOESS curves with 95% confidence intervals between the log RRs and the distance in kilometers.

#### Mapping the association between counties using multidimensional scaling

We evaluate the extent to which patterns of association obtained when looking at the location of pairs of identical sequences are consistent with global spatial structure. To do so, we perform Non-metric Multidimensional Scaling (NMDS) based on the matrix of RR of observing pairs of identical sequences between two counties. We restrict our analysis to the subset of counties for which there was always at least 5 pairs of identical sequences observed with the other counties in the subset. This is done to remove potential noise associated with low number of pairs observed. As the NMDS algorithm requires a measure of similarity between counties, we define the similarity *s_A,B_*between counties *A* and *B* as:

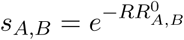

We perform 2-dimensional NMDS using the *vegan* R package [50].

#### Exploring directionality in transmission between Eastern and Western WA

We evaluate whether the timing of identical sequence collection is consistent with transmission rather occurring from Western to Eastern or Eastern to Western WA. We define 4 time periods corresponding the 4 epidemic waves experiences by WA during our study period (Figure S33):

- Wave 4: March 2021 - June 2021
- Wave 5: July 2021 - November 2021
- Wave 6: December 2021 - February 2022
- Wave 7: March 2022 - August 2022

For each of these time periods, we compute the proportion of pairs of identical sequences first collected in Eastern WA among pairs of identical sequences observed in both Eastern and Western WA that were not collected on the same day. We report 95% binomial proportion CI around these proportions.

To explore whether our conclusions could be explained by differences in testing behaviours between Eastern and Western WA, we conduct a sensitivity analysis by imputing the date of symptom onset.

### Mobility analyses

#### Evaluating the relationship between genetic and mobility data

We compute the Spearman correlation coefficient between the RR of observing identical sequences between two counties and the RR of movement between two counties (both from mobile phone derived and commuting mobility data) as well as the geographic distance between counties’ centroids. We determine the percentage of variance in the genetic data explained by the mobility data by fitting generalised additive models (GAMs) predicting the RR of observing identical sequences based on the RR of movement between two counties, both on a logarithmic scale, using a thin plate smoothing spline with 5 knots. For the GAM analyses, we remove pairs of counties for which the number of pairs of identical sequences or the total mobility flow is equal to 0, which ensures that both the RR of observing identical sequences and the RR of movement are strictly positive. We also fit a GAM between the RR of observing identical sequences between two counties (on a logarithmic scale) and the distance between counties centroids. We repeat these analyses at the regional level, instead of at the county one.

#### Identifying outliers in the relationship between genetic and mobility data

We define outliers in the relationship between genetic and mobility data as pairs of counties for which the absolute value of the scaled Pearson residuals of the GAM is greater than 3. As we expect RRs computed from a low number of pairs of identical sequences to be more noisy, we focus on pairs of counties for which at least 100 pairs of identical sequences are observed throughout the study period.

### Characterising spread between postal codes with male prisons

#### Centrality analysis

We characterise transmission between the 10 postal codes with male state prisons by performing a network centrality analysis. We consider a network with 10 nodes corresponding to these different postal codes. We define the weight of each edge as the RR of observing identical sequences between the two postal codes that define the nodes connected by this edge. This results in a fully connected network. For each node (postal code with a male state prison), we compute eigenvector centrality scores using the R *igraph* package. This centrality score measures a node’s influence in the network: nodes have higher scores when they are connected to other influential nodes.

#### Identifying large clusters of identical sequences shared between different male prison postal codes

We define large clusters of identical sequences in the prison networks as clusters of identical SARS-CoV-2 sequences (i) which are observed in at least two postal codes with male prisons and (ii) with at least 15 sequences in prison postal codes.

### Age analyses

#### Evaluating the relationship between genetic and social contact data

We quantify the association between the relative risk 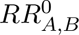 of observing identical sequences between two age groups *A* and *B* and the relative risk of contacts 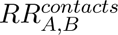 between these two groups using a generalised additive model (GAM) on a logarithmic scale. We report the percentage of variance in the RR of identical sequences explained by the RR of contact from the GAM. We also report the Spearman correlation coefficient between 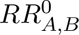 and 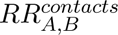.

#### Investigating age-specific transmission patterns across spatial scales

To understand how age-specific transmission patterns vary across spatial scales, we compare the RR of observing identical sequences between age groups using all pairs of identical sequences, using only pairs of identical sequences in different postal codes and using only pairs of identical sequences in different counties.

#### Exploring typical transmission direction between age groups

We use sequence collection dates to explore transmission direction between age groups across 4 periods (March 2021 - June 2021, July 2021 - November 2021, December 2021 - February 2022 and March 2022 - August 2022). To facilitate the interpretation of these results, we introduce an earliness score that measures the contribution of a given age group to transmission to other age groups. For an age group *A*, this score is equal to the proportion of pairs of identical sequences first observed in age group *A* among all pairs of identical sequences observed in age groups *A*. We also report 95% binomial confidence interval around this score.

To explore whether our conclusions could be explained by differences in testing behaviours between age group, we conduct a sensitivity analysis by imputing the dates of symptom onset and using symptom onset dates instead of sequence collection ones (Figure S22). We also compute earliness scores on each of the 1,000 datasets with imputed symptom onset dates using the same definition as that based on sequence collection dates. We then report the median earliness score across all 1,000 datasets as well as an uncertainty range defined as the minimum lower bound and the maximum upper bound of the 95% binomial confidence interval around this score for each of the imputed dataset.

#### Relative risk of observing identical sequences between vaccination groups

Available matched patient information include details regarding the individuals’ vaccination statuses upon positive test:

- No valid vaccination record (denoted Unvaccinated)
- Completed primary series (denoted Vaccinated)
- Completed primary series with an additional dose (denoted Boosted)

Here, we use this information to quantify mixing between groups characterised by their vaccination status. We focus on the mixing between vaccination groups within age groups to avoid biases coming from age group and vaccination status being correlated. Among sequences collected within each period (4 waves) and age groups in decade, we compute the RR of observing identical sequences between vaccination groups. We only include the Boosted vaccination group for wave 6 (Omicron BA.1 wave) for age groups older than 10, and from wave 7 for the 0-9y. We only include the 0-9y in our analysis from wave 6 (Omicron BA.1 wave) and the 10-19y from wave 5 (Delta wave).

To quantify the tendency of individuals of transmitting to individuals with the same vaccination status, we compute for each vaccination groups (*V*_1_, *V*_2_) the ratio *RR_V_*_1_*_,V_*_2_ */RR_V_*_1_*_,V_*_1_. Values lower than 1 indicate that the enrichment of pairs of identical sequences is greater within the same vaccination group than between different vaccination groups. Such values suggest assortativity in mixing patterns between vaccination groups.

### Sensitivity analysis for the typical transmission direction analysis

In the former paragraphs, we describe an approach based on the timing of pairs of identical sequences to better understand the typical transmission direction between groups. The interpretation of this pair-based analysis is complicated by several factors. First, clusters of identical sequences can span more than two groups. Second, even in instances where clusters only span two groups, counting pairs could improperly capture transmission direction, for example if local transmission of the cluster is occurring within the two groups. We implemented this pair-based approach as an intuitive exploration of whether the timing of sampling of identical sequences might provide some signal about transmission direction. This pair-based approach is crude but yet interesting as we do expect groups that tend to be the source to more often be observed first within clusters or pairs of identical sequences. As a sanity check, we perform a sensitivity analysis relying on clusters of identical sequences that are only observed within two groups. We define the *source* group as the group of the earliest collected sequence within the cluster of identical sequences. We remove ambiguous clusters, meaning clusters with two potential source groups, from the analysis. For clusters observed in groups *A* and *B*, we compute the proportion of clusters with source group *A*. We refer to this proportion as “proportion from clusters” to distinguish it from the “proportion from pairs” that we use in our main analysis. We compute 95% confidence interval around the proportion from clusters. This proportion from clusters should be more robust that the proportion from pairs but tends to be noisier as we are computing the proportion from less observations. We then compare the proportion obtained from pairs and from clusters. We compute the Spearman correlation coefficient between these two proportions using all pairs of groups or only pairs of groups for which the confidence intervals don’t contain 50% for the two proportions.

### Deriving the distribution of the number of mutations conditional on the number of generations separating two infected individuals

In this section, we derive the probability distribution of the number of mutations *M_AB_* separating the consensus genomes of two infected individuals *A* and *B* conditional on the number of transmission generations *G_AB_* separating them.

#### Generation time distribution

We assume that the generation time (i.e. the average duration between the infection time of an infector and an infectee) follows a Gamma distribution of shape *α* and scale *β*. The time between *g* generations then follows a Gamma distribution of shape *g*·*α* and scale *β* assuming independence of successive transmission events. Let *f_α,β_*(·) denote the probability density function of a Gamma distribution of shape *α* and scale *β*.

#### Mutations events

Let *M_AB_* denote the number of mutations separating their infecting viruses. Let *µ* denote the mutation rate of the virus (in mutations per day). Let 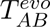 denote the evolutionary time separating *A* and *B* (in days).

Assuming a Poisson process for the occurrence of mutations, we have:

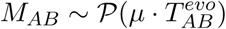

#### Distribution of the number of mutations conditional on the number of generations

Let *G_AB_* denote the number of generations separating two infected individuals *A* and *B* belonging to the same transmission chain.

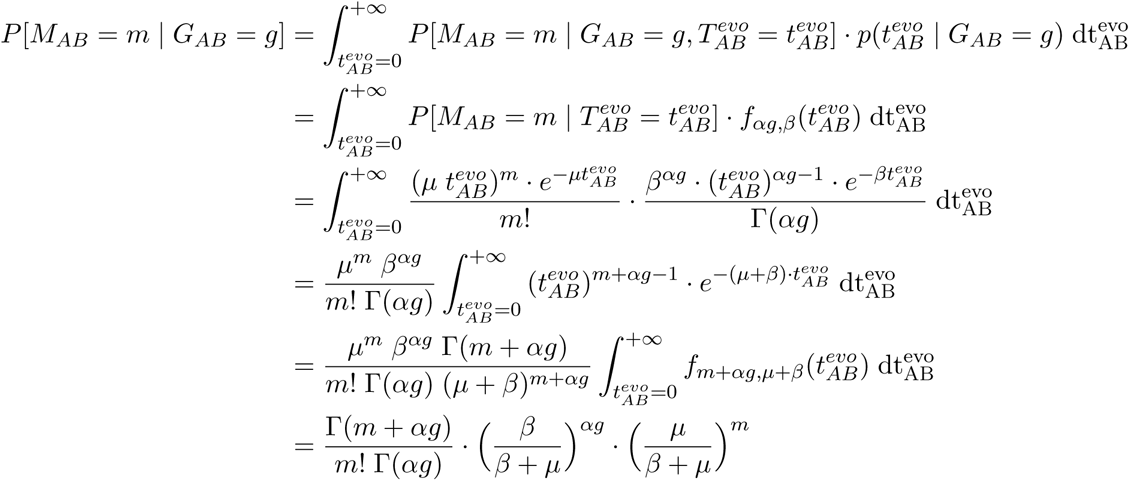

which is the probability mass function of a negative binomial distribution of parameters:

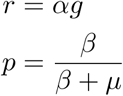

#### Application to SARS-CoV-2

We compute these probabilities for SARS-CoV-2 considering a mutation rate *µ* = 8.98 · 10^−2^ substitutions per day (32.76 substitutions per year) [51]. We assume that the generation time is Gamma distributed with a mean 5.9 days and standard deviation 4.8 days [52].

### Simulation study to evaluate the ability of the RR framework to describe migration patterns

We conduct a simulation study to evaluate how our RR framework performs under different sequencing scenarios. We also compare the results obtained from a phylogeographic analysis.

#### Simulating synthetic sequence data

We use ReMASTER [53] to simulate an SEIR epidemic in a structured population with 5 demes, each populated with 100,000 inhabitants. We simulate an epidemic characterised by a basic reproduction number *R*_0_ of 2 with a daily time-step. We initiate the simulations by introducing a single infected individual (compartment *I*) in the population (group of index 0). We consider a pathogen with a 3,000 kb genome evolving following Jukes-Cantor evolution model with a substitution rate of 3 · 10^−5^ substitutions per site per day. Upon infection, infected individuals enter an Exposed (E) compartment during which they are not infectious yet and that they exit at a rate of 0.33 per day. They then enter an Infectious (I) compartment where they are infectious that they exit at a rate of 0.33 per day. Sequencing occurs upon exit of the I compartment. Given that our RR does not account for directionality in transmission, we considered a scenario with symmetric migration rates. We draw migration rates between demes from a log-Uniform distribution of parameters (10^−3^, 10^−1^).

We then explore two sequencing scenarios. In an unbiased scenario, we assume that each individual has the same probability of being sequenced in each deme. In a biased scenario, we assume that the sequencing probability varies between demes. We draw deme-specific relative sequencing probabilities from a log-Uniform distribution of parameters (10^−3^, 10^−1^). In the unbiased scenario, we fix sequencing probability to the mean of the sequencing probabilities across demes in the biased scenario. We explore different sequencing intensities by scaling these probabilities by different multiplicative factor (Table S1):

- a scaling factor of 0.1 resulting in a mean sequencing probability of 0.43% and a dataset of around 1700 sequences (used for the DTA analyses)
- a scaling factor of 0.5 resulting in a mean sequencing probability of 2.16% and a dataset of around 8600 sequences (used for the RR and the DTA analyses)
- a scaling factor of 2 resulting in a mean sequencing probability of 8.66% and a dataset of around 34,500 sequences (used for the RR analyses)

#### Discrete trait analysis

We conduct phylogeographic inference using symmetric discrete trait analysis (DTA) [9] using the Bayesian stochastic search variable selection (BSSVS) model implemented in BEAST 1.10.4 [54] applied to the synthetic data simulated in our two sequencing scenarios. In order to isolate the accuracy and precision of the phylogeographic reconstruction, we run our discrete trait analysis using an empirical tree that is generated directly from ReMASTER simulations. Directly inputting such a tree is not possible in real-world scenarios where the genealogical tree must be (noisily) estimated from empirical sequence data. In this case, it serves a demonstration of the power of DTA when provided perfect genealogical signal. The empirical tree approach also requires substantially less computation and so allowed us to analyse datasets of thousands of sequences using DTA in acceptable time.

Two independent Markov chain Monte Carlo (MCMC) procedures are run for 2.5 · 10^8^ iterations and sampled every 1,000 iterations. Resulting posterior distributions are combined after discarding initial 10% of sampled trees as burn-in from each of them. We use Tracer 1.7 [55] to assess convergence and to estimate effective sampling size (ESS), ensuring ESS values greater than 200 for each migration rate estimate. We adjust the estimated migration rates by the estimated rate scalar in order to calculate the per-day rates of transition between demes.

To evaluate how estimating the genealogical tree from empirical sequence data impacts both results and computing times, we perform an additional phylogeographic analysis based on simulated but this time jointly inferring the genealogical tree and migration rates. We run this model for 24 days (corresponding to 475,733,000 MCMC steps), until each migration rate parameter has an ESS greater than 200.

#### Relative risk analysis

We compute the RR of observing identical sequences between two demes *i* and *j* and compare these RR to the daily probability *p_i,j_* of migration between these two demes which is computed as:

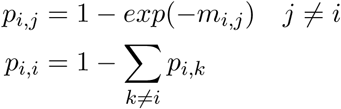

where *m_i,j_* is the migration rate between demes *i* and *j*. We generate 95% subsampling confidence intervals around the RRs using an 80% subsampling rate.

#### Simulation study to explore the expected relationship between the RR of observing identical sequences between groups and the RR of contacts between groups

We perform a simulation study to characterise the expected relationship between the RR of observing identical sequences between groups and the RR of contacts between these groups. We illustrate this by looking at transmission between age groups but we expect a similar relationship between the RR of observing identical sequences between regions and the RR of movement between these regions.

To do so, we generate clusters of identical sequences including the age group of the corresponding infected individuals assuming a probability that an infector and an infectee have the same consensus sequences *p* of 0.7 (close to the value we previously estimated for SARS-CoV-2 [8]), a reproduction number *R* of 1.2 and a sequencing fraction *p_seq_* of 0.1.

We use the contact matrix estimated by Mistry et al. [29] to characterise disease transmission between age groups. We assume that the probability *p_A,B_* for a contact from an infected individual in age group *A* to occur with a susceptible individual in age group *B* is equal to:

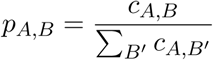

where *c_A,B_* is the mean daily number of contacts that an individual in age group *A* has with individuals in age group *B*. We introduce an age-specific reproduction number (*R_A_*) describing the average number of secondary cases infected by a single primary case within age group *A*. As different age groups have different average daily total number of contacts, the age-specific reproduction number varies between age groups. It can be derived as:

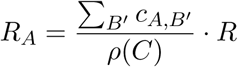

where *ρ*(*C*) is the maximum eigenvalue of the matrix *C* = (*c_A,B_*) [56]. We then simulate individual clusters of identical sequences using the following steps:

1. **Initialization** Draw the age of the primary case *A_primary_* from a uniform distribution.
2. **Simulate clusters (successive infections with identical genomes)** While the number of generations is lower than 10 For each individual infected at the previous generation

- Let *A* denote the age of this infectious individual.
- Draw the number of infections with the same genotype: *I* ∼ P((*p* · *R_A_*)^−1^) (Poisson distribution).
- Draw the age of these individuals: (*A*_1_, *…, A_I_*) ∼ M(*I, n_age_,* (*p_A,_*_1_, *…, p_A,nage_*)) where M(*n, k,* (*p*_1_, *…, p_k_*)) is a multinomial distribution with *n* trials, *k* possible events with probabilities (*p*_1_, *…, p_k_*)
3. **Simulate sequencing** Draw the sequencing status of each cluster member from a Bernoulli distribution of parameter *p_seq_*.

We end simulations after 10 generations to minimise computational costs.

### Downsampling approach to explore the genome dataset size required to compute relative risks

We implement a downsampling strategy to understand the amount of sequencing data required to compute RR estimates. We consider genome datasets of the following sizes:

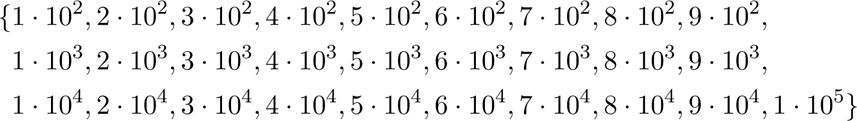

For each of these dataset sizes, we generated 100 downsampled datasets from our WA sequencing data. For each of these downsampled datasets, we compute the RR of observing identical sequences between age groups (Figure S34). To understand how the number of groups studied impacts the amount of data required, we also compute RR of observing identical sequences between aggregated age groups:

- 0-39y and over 40y for 2 age groups,
- 0-29y, 30-59y and over 60y for 3 age groups,
- 0-19y, 20-39y, 40-59y and over 60y for 4 age groups,
- 0-9y, 10-19y, 20-29y, 30-39y, 40-49y, 50-59y, 60-69y, 70-79y and over 80 (standard definition used throughout the paper) for 9 age groups.

We compute the error between the RR obtained from a subsampled dataset *RR^d^* and the RR from the full dataset *RR^f^* as:

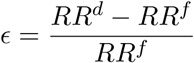

For each pair of age groups, we then compute the number of pairs of identical sequences required for the error to be below 10%.

### Simulation study to evaluate the possibility to extend the Hamming distance threshold

In this work, we assess how pairs of infected individuals whose infected genome is separated by 0 mutation can help characterise population level transmission patterns. We apply this method to SARS-CoV-2 sequences from WA but our approach should be broadly applicable for epidemics caused by pathogens where the timescale of mutation events is similar to that of transmission events. In this section, we describe a simulation approach to understand how a pathogen’s mutation rate could impact the optimal Hamming distance threshold to apply our RR framework.

We implement the same simulation framework as the one described in the section “Simulation study to evaluate the ability of the RR framework to describe migration patterns”. We consider that each infection has the same probability of being sequenced (equal to 4.33 %, corresponding to a sequencing probability scaling factor of 1). We explore a range of scenarios for the pathogen’s mutation rate. To do so, we introduce a multiplicative scaling factor for the baseline pathogen’s mutation rate (3 · 10^−5^ substitutions per site per day) with values ranging between 0.1 and 10. For each multiplicative scaling factor, we perform 100 replicate simulations.

For each simulated epidemic, we count the number of pairs separated by less than *d* mutations between two regions (for *d* varying between 0 and 10). In certain scenarios (for example those characterised by a high mutation rate and a low Hamming distance threshold *d*), we sometimes don’t observe any pairs less than *d* mutations away in a specific group. To be able to compute the RR even in those scenarios, we report a modified version of the RR of observing sequences separated by less than *d* mutations between two regions:

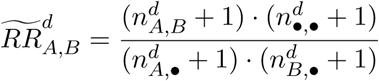

with the same notations as the ones used in the definition of the RR of observing sequences less than *d* mutations away in groups *A* and *B*.

We then compute the Spearman correlation coefficient between the relative risk for pairs of sequences less than *d* mutations apart of being in two regions and the daily migration probability between these regions. In simulations where the standard deviation of the RR is equal to 0 (all modified RRs have the same value), we assume that the Spearman correlation coefficient is equal to 0 (RRs are not informative about migration rates).

## Data Availability

GISAID and are publicly available with standard metadata (generally consisting of date of sample collection and sometimes county of sample collection). More detailed metadata curated by Washington State Department of Health (WA DOH) of county, postal code, age group and vaccination status were shared with the Fred Hutchinson Cancer Center under a Data Sharing Agreement for Confidential Data with an associated IRB. These more detailed metadata are not currently shared publicly while we seek clearance with WA DOH. GISAID accessions and a sequence-level acknowledgements table are provided in the GitHub repository associated with this manuscript (https://github.com/blab/phylo-kernel-public).

## Data and code accessibility

Code to reproduce our analyses can be found at https://github.com/blab/phylo-kernel-public [49]. A step by step tutorial describing how to implement our RR approach is available at https://github.com/CecileTK/tutorial-rr-identical-sequences [57]. We also share counts of pairs of identical sequences between population groups (counties and age groups) obtained from the WA Sentinel surveillance data. All sequences referenced in this manuscript are publicly shared to GISAID and are publicly available with standard metadata (generally consisting of date of sample collection and sometimes county of sample collection). More detailed metadata curated by Washington State Department of Health (WA DOH) of county, postal code, age group and vaccination status were shared with the Fred Hutchinson Cancer Center under a Data Sharing Agreement for Confidential Data with an associated IRB. These more detailed metadata are not currently shared publicly while we seek clearance with WA DOH. GISAID accessions and a sequence-level acknowledgements table are provided in the GitHub repository associated with this manuscript [49].

## Author contributions

CTK and TB conceived the study. CTK developed the methods. CTK, ACP and MIP performed the analyses with input from CV and TB. LF, HX, KK, AW, ALG, PR, JMP, AD, HH, DM, PD, LG, CDF, ER, JS, DR, AT, CY, FA and AB collected and curated the data. CTK, AP, MP and TB wrote the manuscript. All authors reviewed and edited the manuscript.

## Acknowledgments

We would like to thank Sixtine Gurrey, Lara B. Strick and Michael A. Santoro for insightful discussions on the control of SARS-CoV-2 in WA Correction Centers. We gratefully acknowledge all data contributors, i.e., the Authors and their Originating laboratories responsible for obtaining the specimens, and their Submitting laboratories for generating the genetic sequence and metadata and sharing via the GISAID Initiative, on which this research is based.

## Ethics approval

The Washington State and University of Washington Institutional Review Boards determined this project to be surveillance activity and exempt from review; the need for informed consent was waived through this determination. Under Washington State IRB Exempt Determination 2020-102, symptom onset date, age group, residence county, residence postal code and vaccination history was provided by the Washington Department of Health from the Washington Disease Reporting System for individuals with linked sequenced SARS-CoV-2 samples from March 1, 2021 through December 31, 2022. Sequencing and analysis of samples from the Seattle Flu Study was approved by the Institutional Review Board (IRB) at the University of Washington (protocol STUDY00006181). Sequencing of remnant clinical specimens at UW Virology Lab was approved by the University of Washington Institutional Review Board (protocol STUDY00000408).

## Disclaimer

The findings and conclusions in this report are those of the authors and do not necessarily represent the official position of the US National Institutes of Health or Department of Health and Human Services.

## Supplementary materials

**Figure S1.**
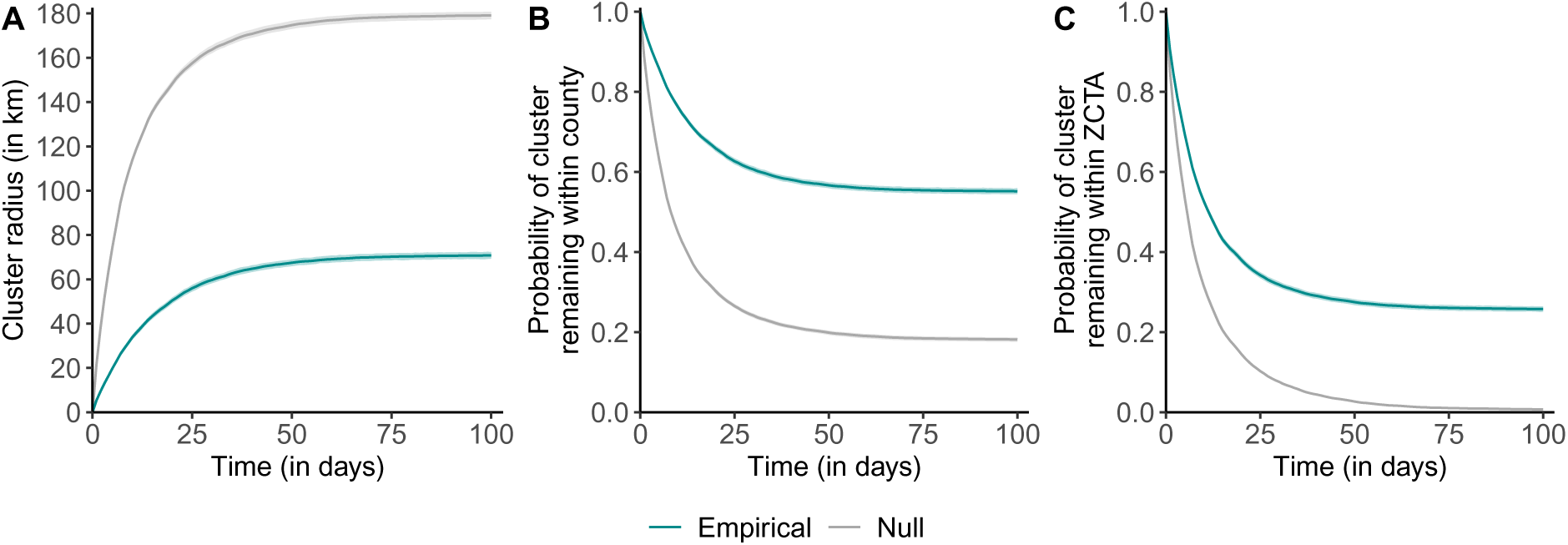
Permutation test to explore the spatio-temporal signal in clusters of identical SARS-CoV-2 sequences. **A.** Radius of clusters of identical sequences as a function of time since first sequence collection. **B.** Probability for all sequences within a cluster of identical sequences of remaining in the same county as a function of time since first sequence collection. **C.** Probability for all sequences within a cluster of identical sequences of remaining in the same ZCTA as a function of time since first sequence collection. The grey shaded areas correspond to 95% confidence intervals of a null distribution generated from 100 simulation where the geographical location of sequences from WA Sentinel surveillance are permuted. The grey lines correspond to the medians the simulated null distributions.

**Figure S2.**
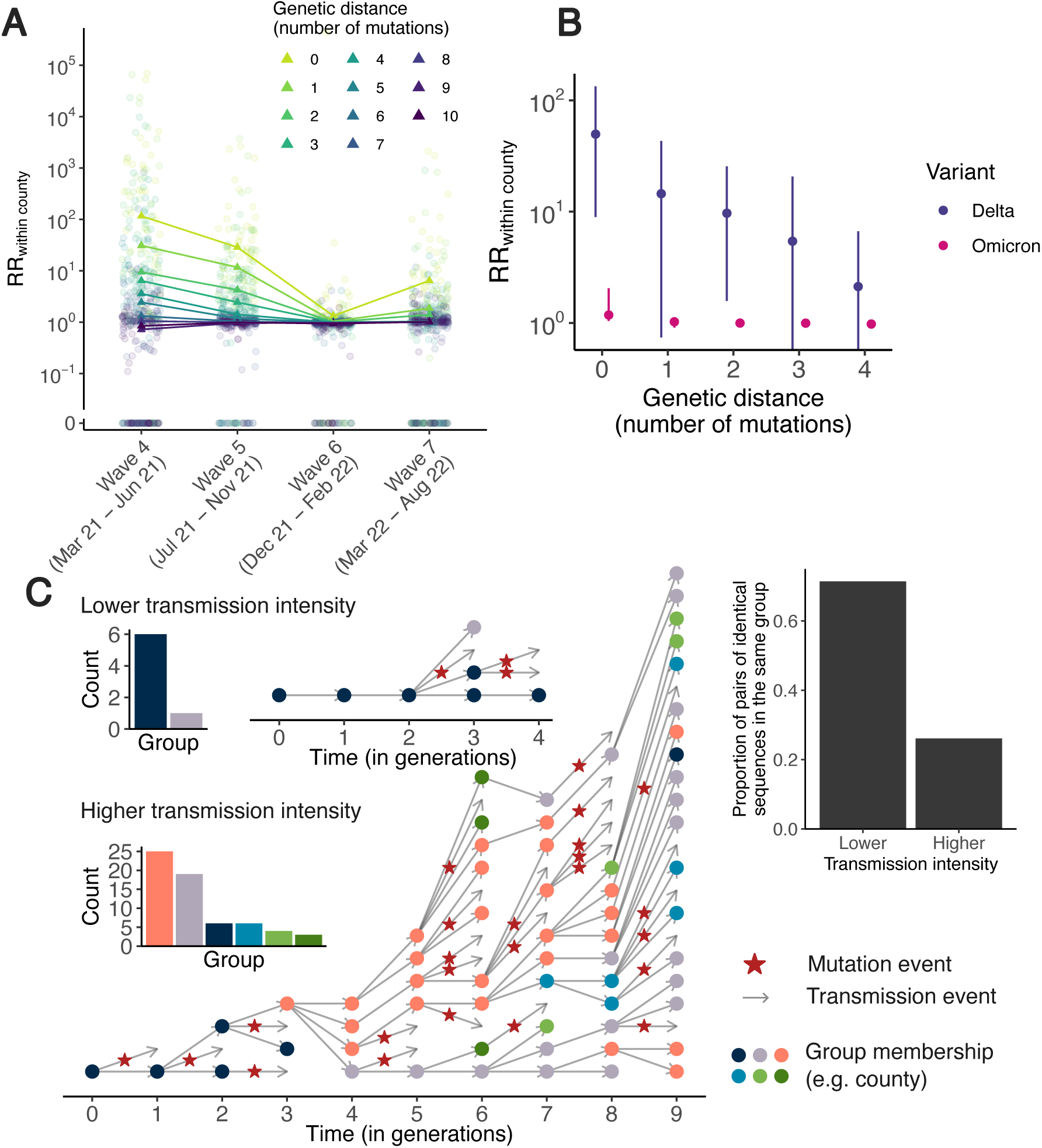
The magnitude of the relative risk of observing sequences at a given genetic distance within the same county is impacted by transmission intensity. **A.** Relative risk of observing sequences at a given genetic distance within the same county across multiple epidemic waves. We defined waves as: March 2021-June 2021 (Wave 4), July 2021-November 2021 (Wave 5), December 2021-February 2022 (Wave 6) and March 2022-August 2022 (Wave 7). In A, circular points correspond to individuals counties and triangles correspond to the median across counties. **B.** Median relative risk of observing pairs sequences within the same county (with IQR) as a function of genetic distance stratified by variant during Wave 6. **C.** A higher transmission intensity results in larger clusters of identical sequences that tend to be more mixed across groups. In C, the two clusters are simulated using a branching process with mutation [8] by assuming the probability for an infector and an infectee to have the same consensus sequence equal to 0.69 and a probability for an infectee of being in the same groups as its infector of 0.7. We consider a reproduction number of 1.2 for the lower transmission intensity scenario and of 2.0 in the higher transmission intensity scenario.

**Figure S3.**
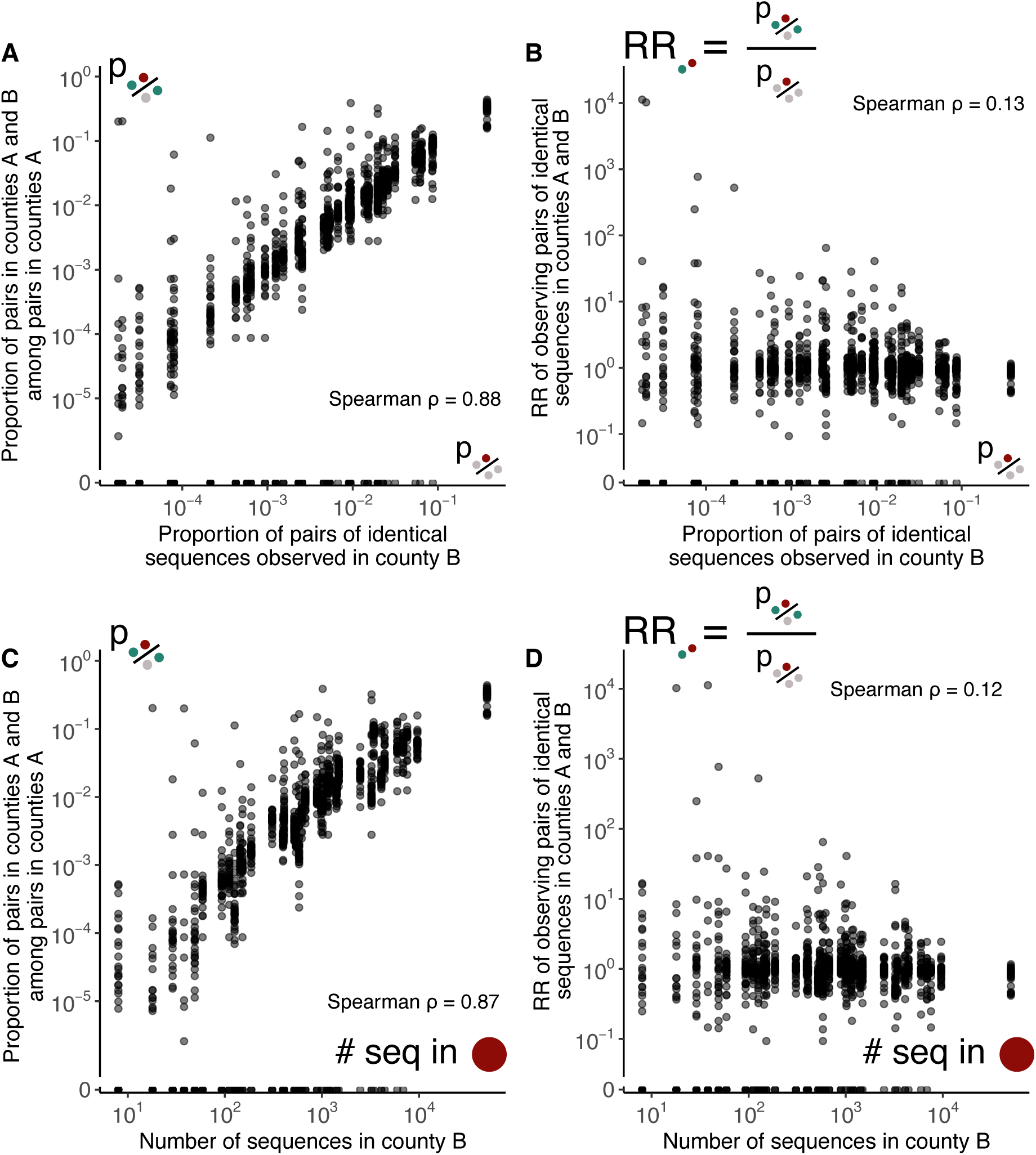
Our measure of relative risk corrects for uneven sequencing between regions. **A.** Proportion of pairs of identical sequences shared between counties A and B among pairs observed in county A as a function of the proportion of pairs of identical sequences observed in county B. **B.** Relative risk for pairs of identical sequences of being observed in counties A and B as a function of the proportion of pairs of identical sequences observed in county B. **C.** Proportion of pairs of identical sequences shared between counties A and B as a function of the number of sequences available in county B. **D.** Relative risk for pairs of identical sequences of being observed in counties A and B as a function of the number of sequences available in county B.

**Figure S4.**
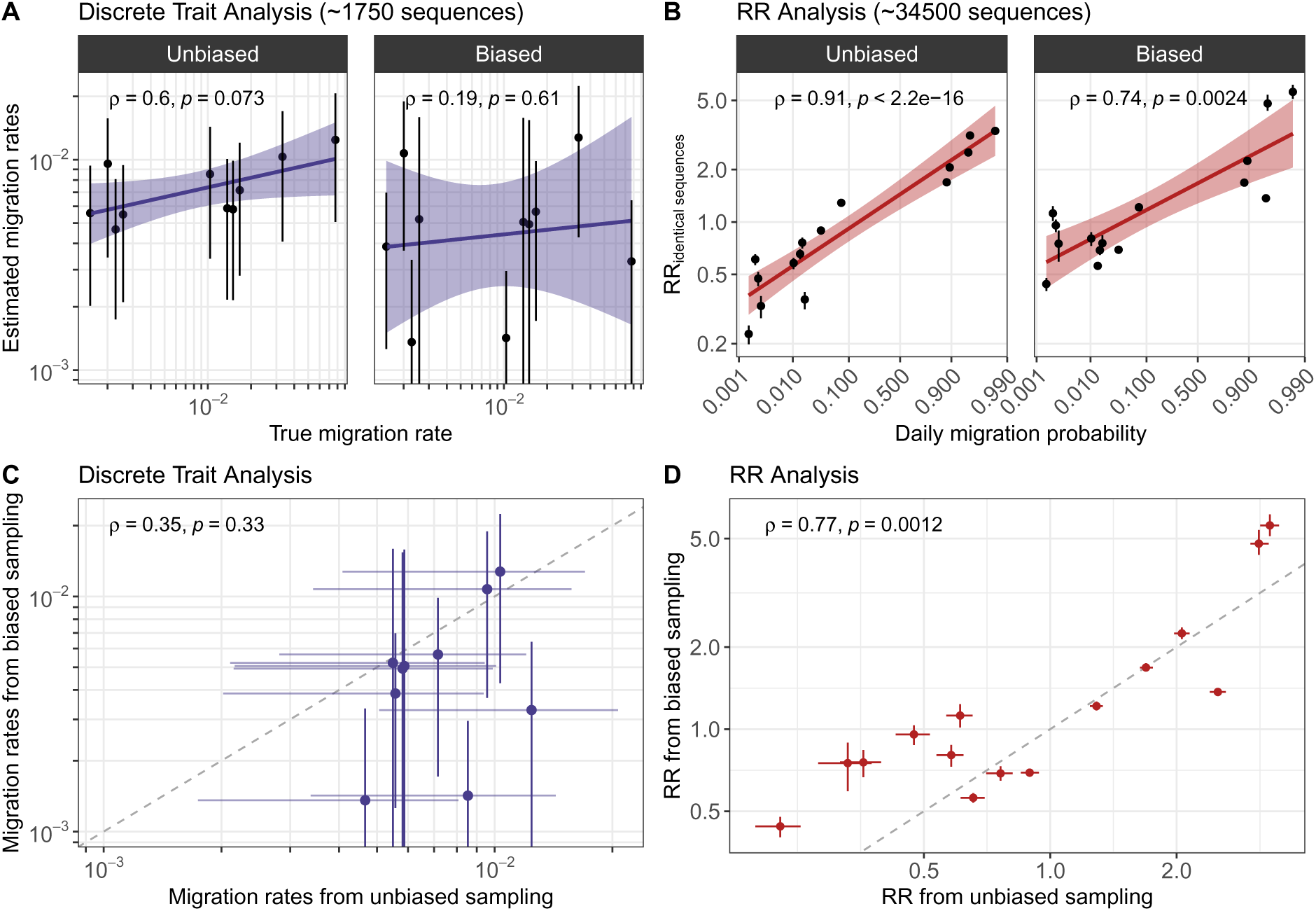
Simulation study exploring the impact of sequencing bias on results from a discrete trait analysis and from our RR framework. **A.** Comparison between migration rates estimated from a discrete trait analysis and the true migration rates used to simulate the sequence data. **A.** Comparison between the relative risk of observing identical sequences between two demes and the weekly migration probability between demes. **C.** Comparison between migration rates inferred from a sequence dataset generated in a biased sampling and an unbiased sampling scenario. **D.** Comparison between the relative risk of observing identical sequences in two groups from a sequence dataset generated in a biased sampling and an unbiased sampling scenario. For the RR, segments indicate 95% subsampling confidence intervals. For the migration rates, segments indicate 95% highest posterior density intervals. For each plot, we indicate the Spearman correlation coefficient (and the associated p-value).

**Figure S5.**
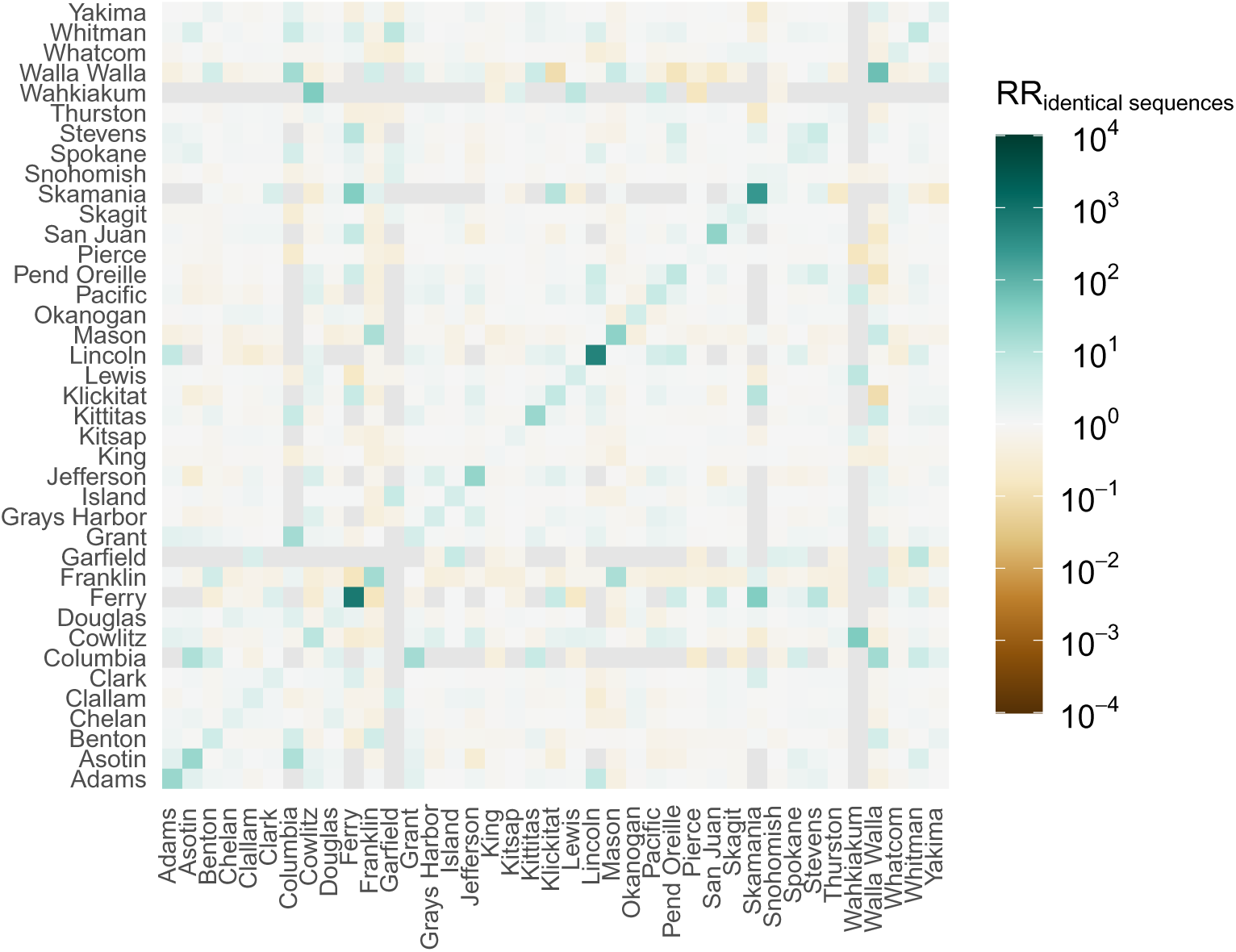
Relative risk of observing identical sequences in two counties. Grey squares correspond to pairs of counties between which no pairs of identical sequences were observed during the study period.

**Figure S6.**
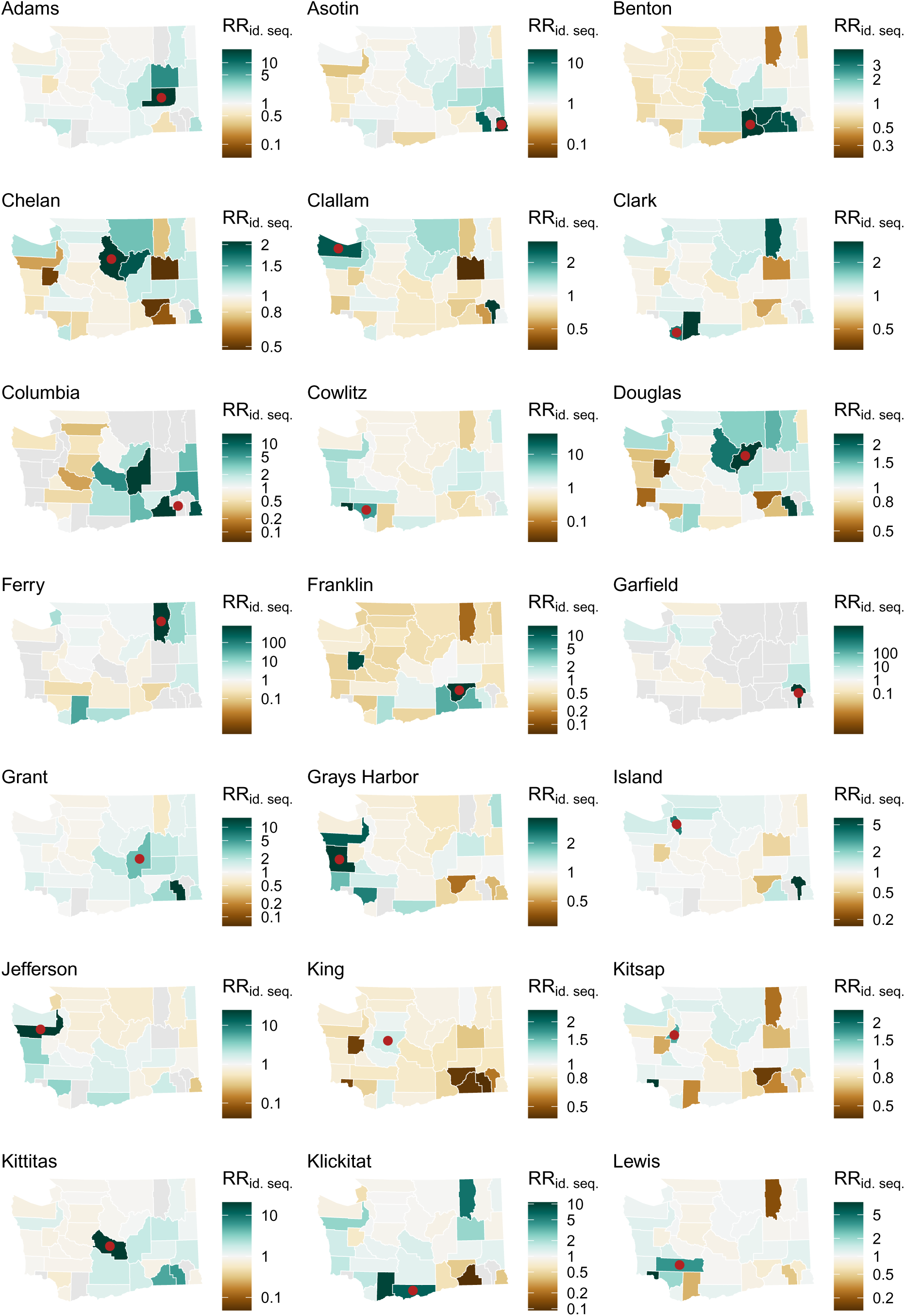
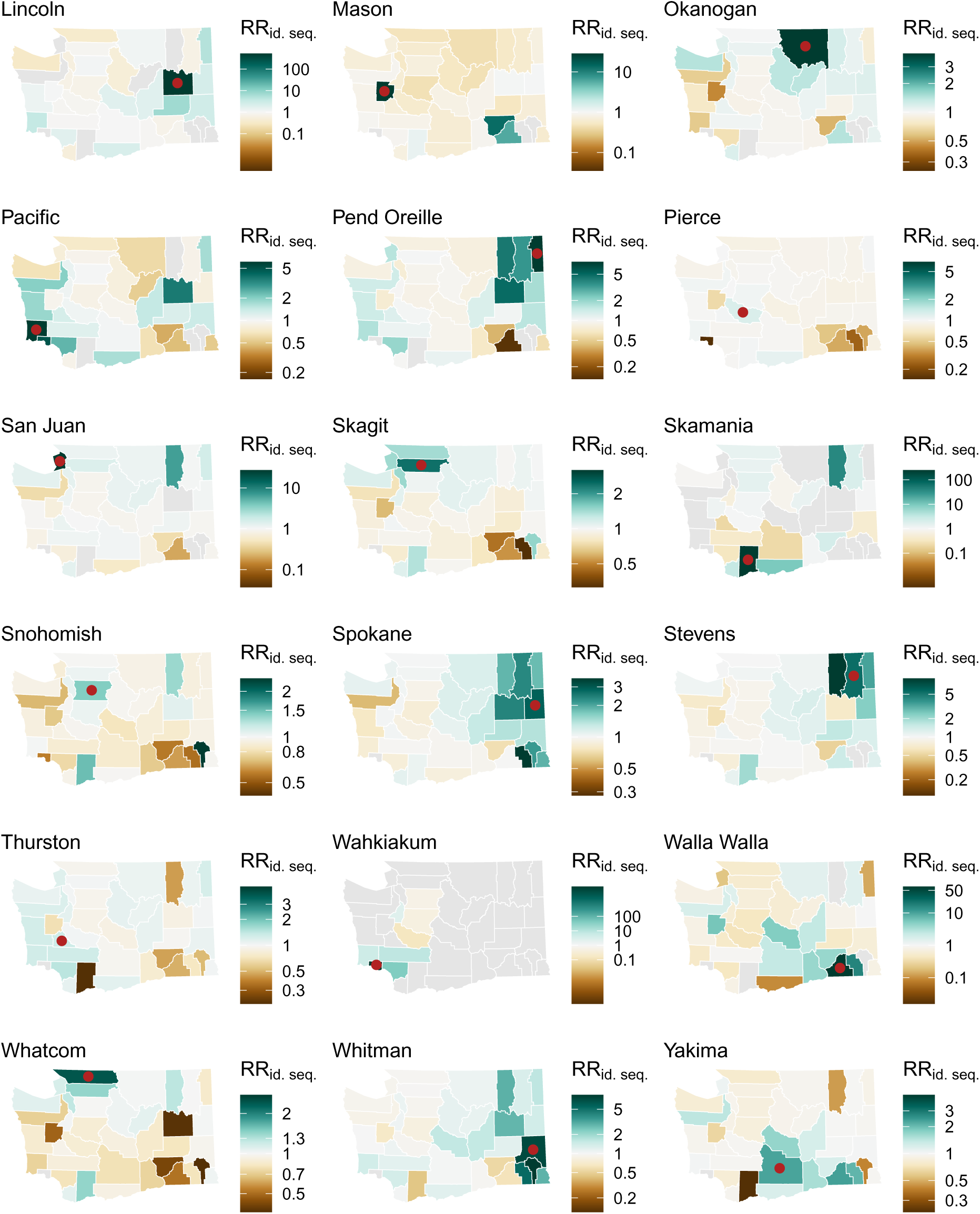
Relative risk of observing pairs of identical sequences between counties. On each map, we represent the relative risk of observing pairs of identical sequences in the county indicated by a red point (map title) and all the other counties in Washington state. Areas are coloured in grey when no pairs of identical sequences are observed. To increase readability, each map has its own colour scale.

**Figure S7.**
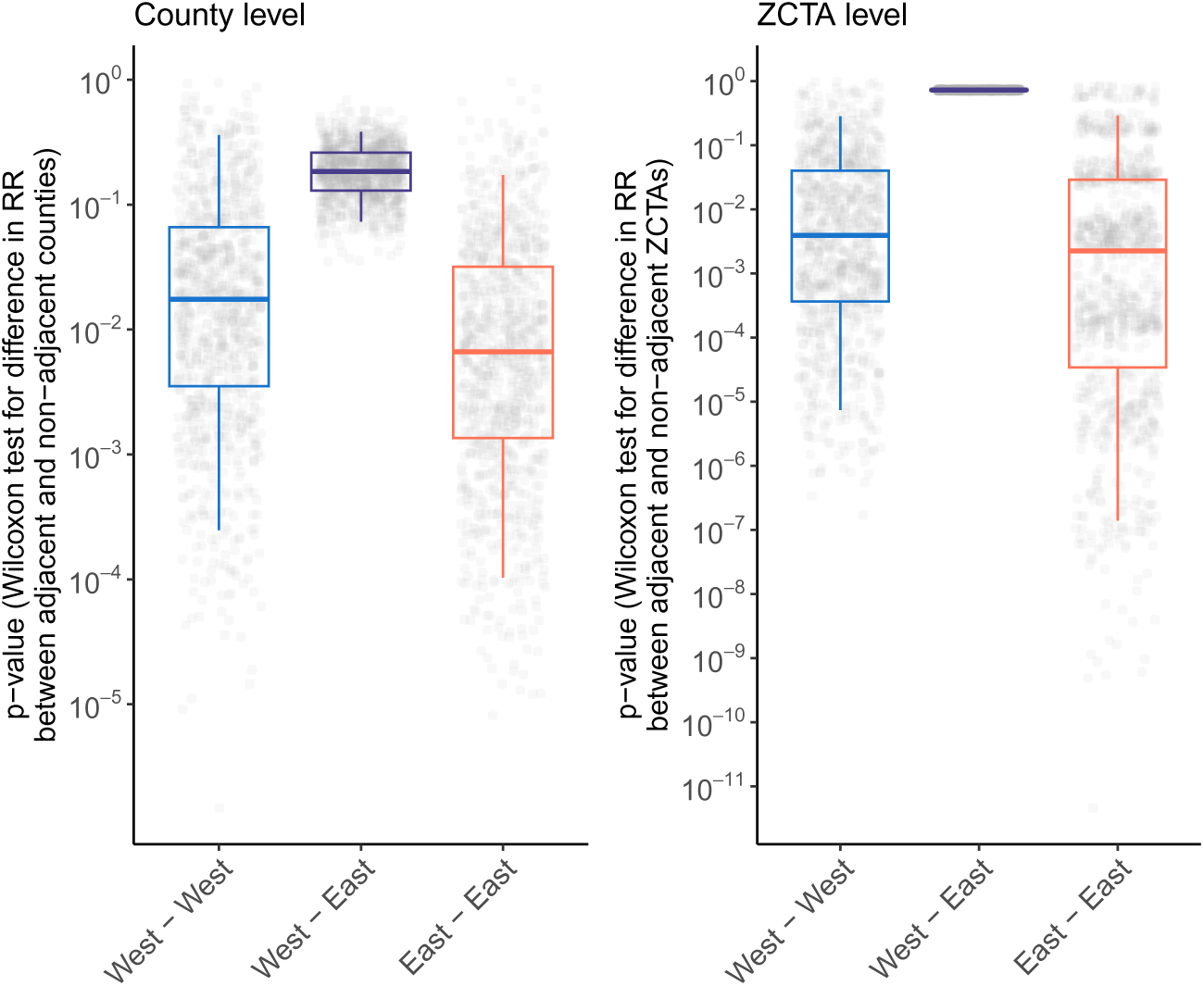
Impact of subsampling on the significance of the association between the relative risk of identical sequences and whether counties and ZCTAs are adjacent or not. We investigate whether our conclusions regarding the significance of the association between the relative risk of identical sequences falling in two distinct counties / ZCTAs and their adjacency status (adjacent / non-adjacent) can be impacted by the number of pairs of counties involved in the comparison (within Eastern WA, within Western WA and between Eastern and Western WA). At the county level, we subsample the pairs of counties involved in these 3 comparisons to 12 adjacent pairs of counties (number of pairs of adjacent counties between Eastern and Western WA) and 132 non-adjacent pairs of counties (number of pairs of non-adjacent counties within Western WA). This ensures that all comparisons are performed on the same number of pairs of counties. On each subsampled dataset, we compute the p-value from a Wilcoxon test evaluating differences between the relative risk of observing identical sequences in adjacent and non-adjacent counties. This is done for 1,000 subsampled datasets. Boxplots indicate the p-values obtained across these different subsampling iterations (5%, 25%, 50%, 75% and 95% quantiles). We do a similar analysis at the ZCTA level.

**Figure S8.**
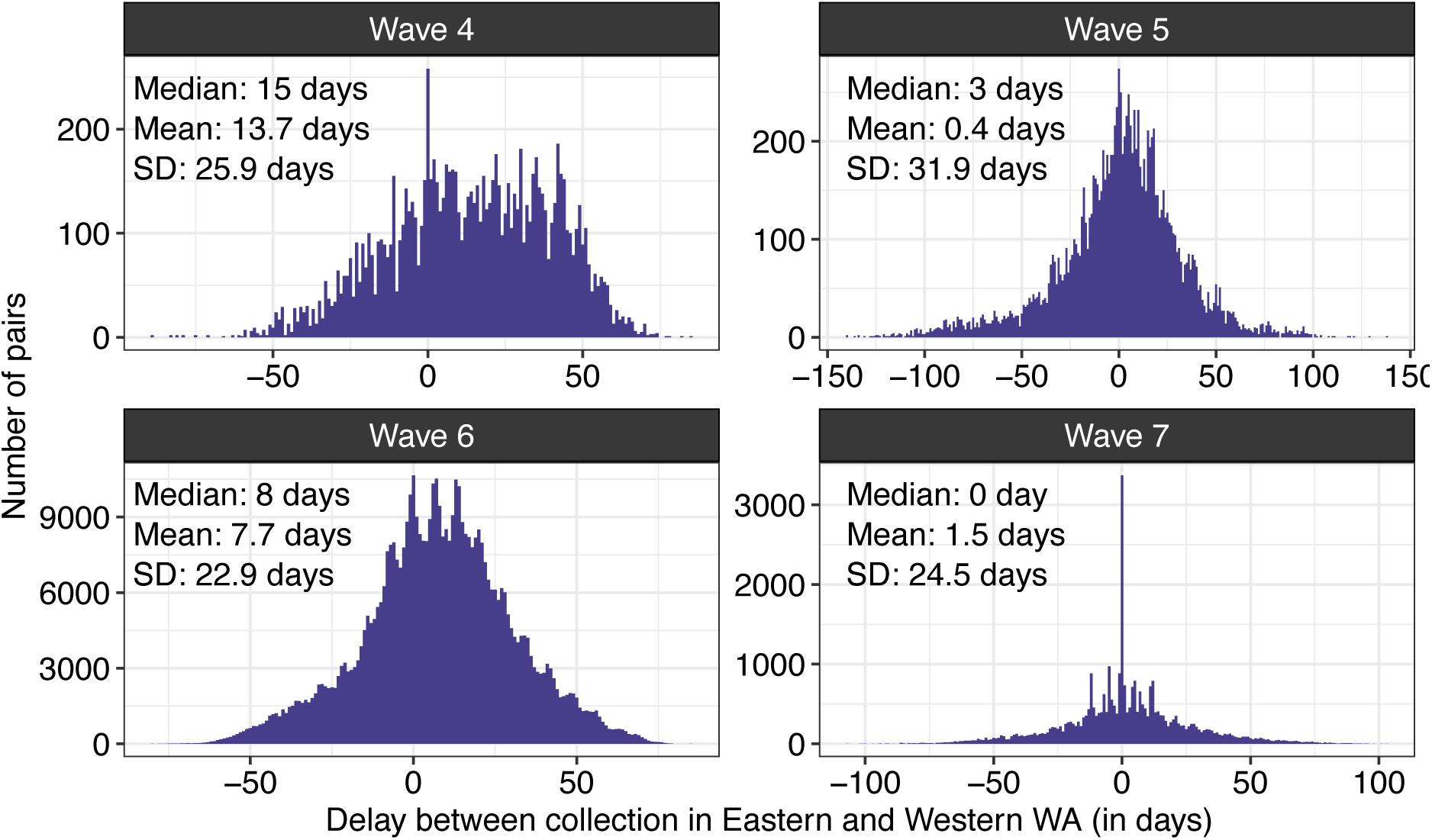
Distribution of the delay between sequence collection within pairs of identical sequences collected between Eastern and Western WA and across epidemic waves.

**Figure S9.**
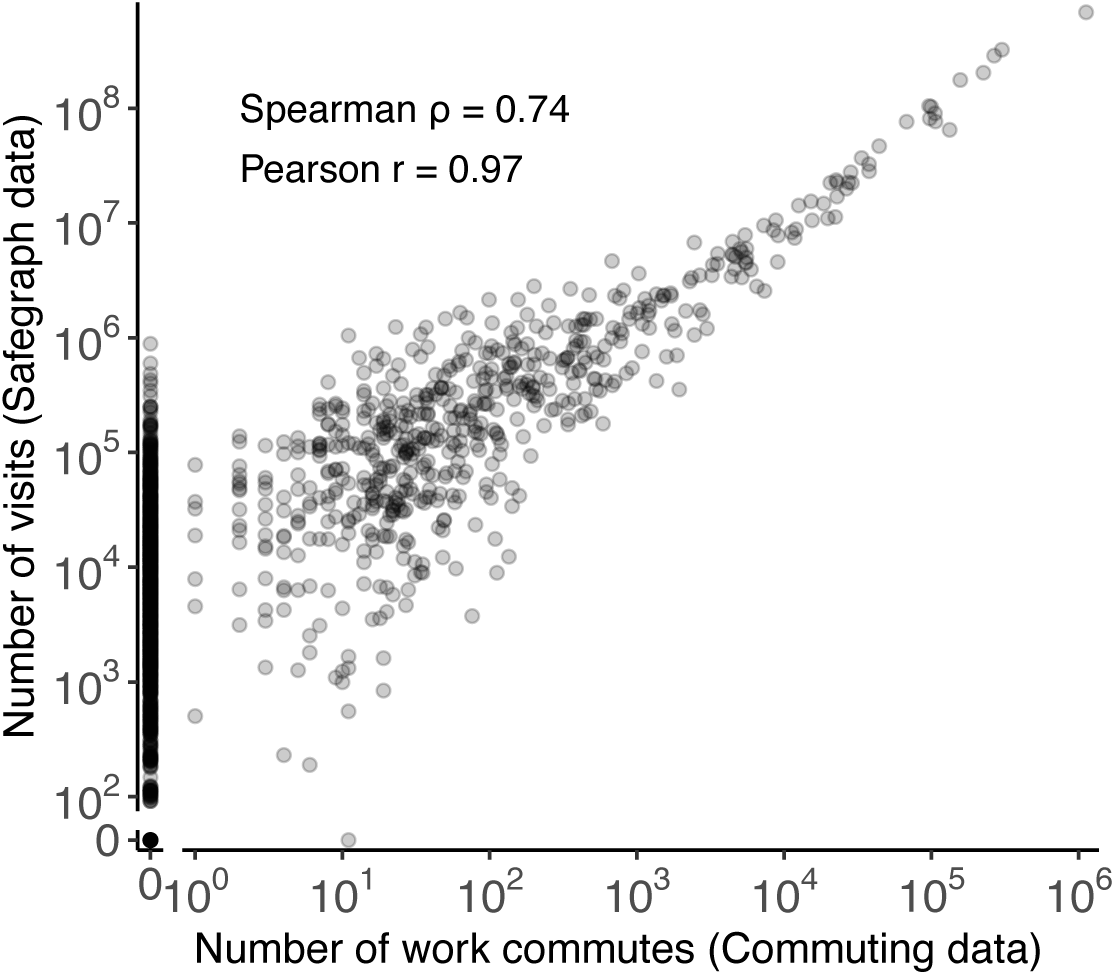
Comparison between the number of directed commuting flows and the number of directed visits between two counties. The number of work commutes is extracted from [17]. The number of visits is estimated using Safegraph *Weekly patterns* mobility data. The comparison is done by matching the origin county in the mobile phone data to the residence county in the workflow data and the destination county in the mobile phone data to the workplace county in the workflow data.

**Figure S10.**
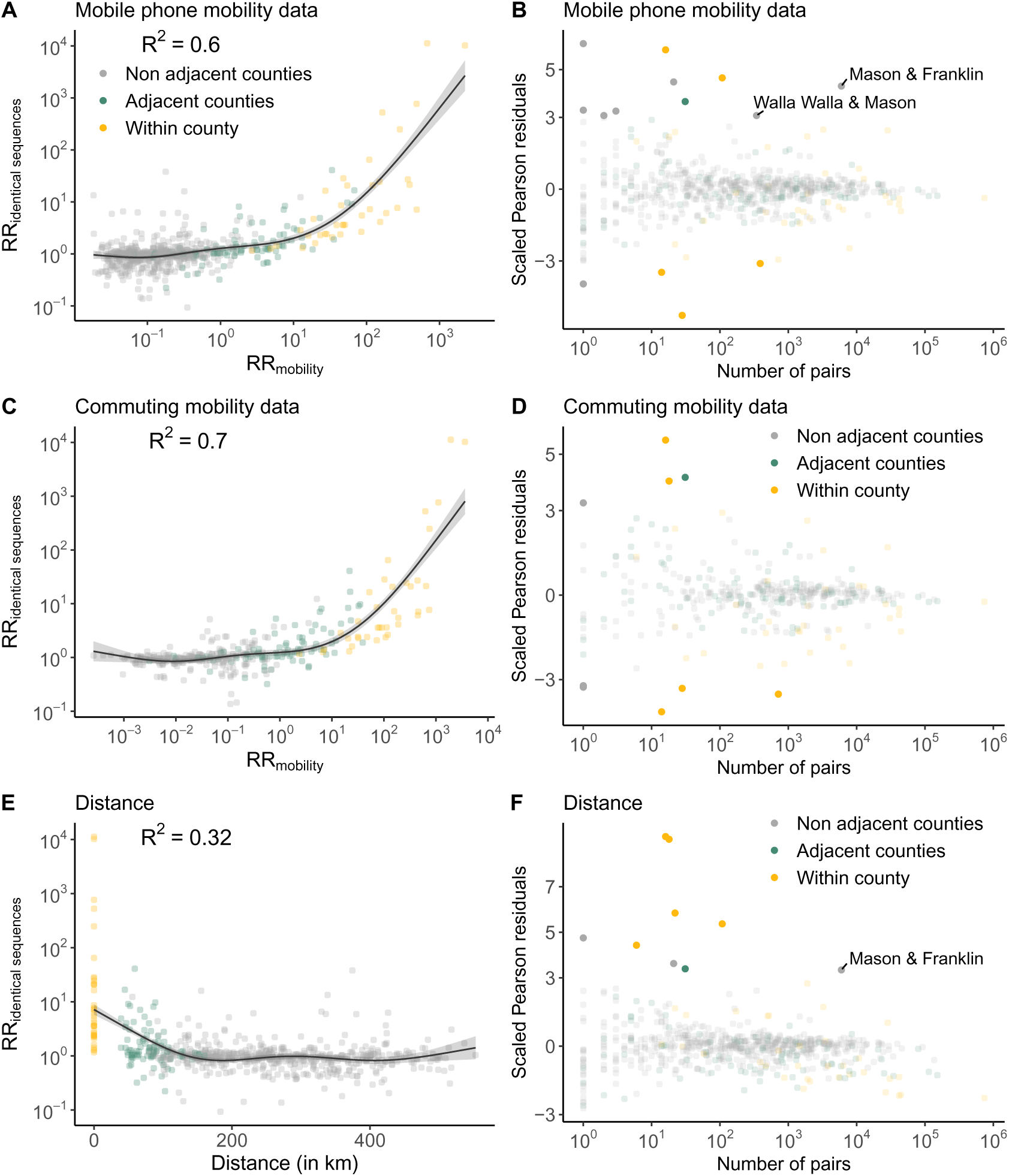
Comparison between the relative risk of observing identical sequences and the relative risk of movement at the county level. **A.** Relationship between the relative risk of observing identical sequences in two counties and the relative risk of movement between these counties as obtained from mobile phone mobility data. **B.** Scaled Pearson residuals of the GAM plotted in A as a function of the number of pairs of identical sequences observed in pairs of counties. **C.** Relationship between the relative risk of observing identical sequences in two counties and the relative risk of movement between these counties as obtained from workflow mobility data. **D.** Scaled Pearson residuals of the GAM plotted in C as a function of the number of pairs of identical sequences observed in pairs of counties. **E.** Relationship between the relative risk of observing identical sequences in two counties and the Euclidean distance between counties centroids. **F.** Scaled Pearson residuals of the GAM plotted in E as a function of the number of pairs of identical sequences observed in pairs of counties. In B, D and F, we label pairs of non-adjacent counties sharing at least 100 pairs of identical sequences and for which the absolute value of the Scaled Pearson residual is greater than 3. The trend lines correspond to predicted relative risk of observing identical sequences in two regions from each GAM. *R*^2^ indicate the variance explained by each GAM.

**Figure S11.**
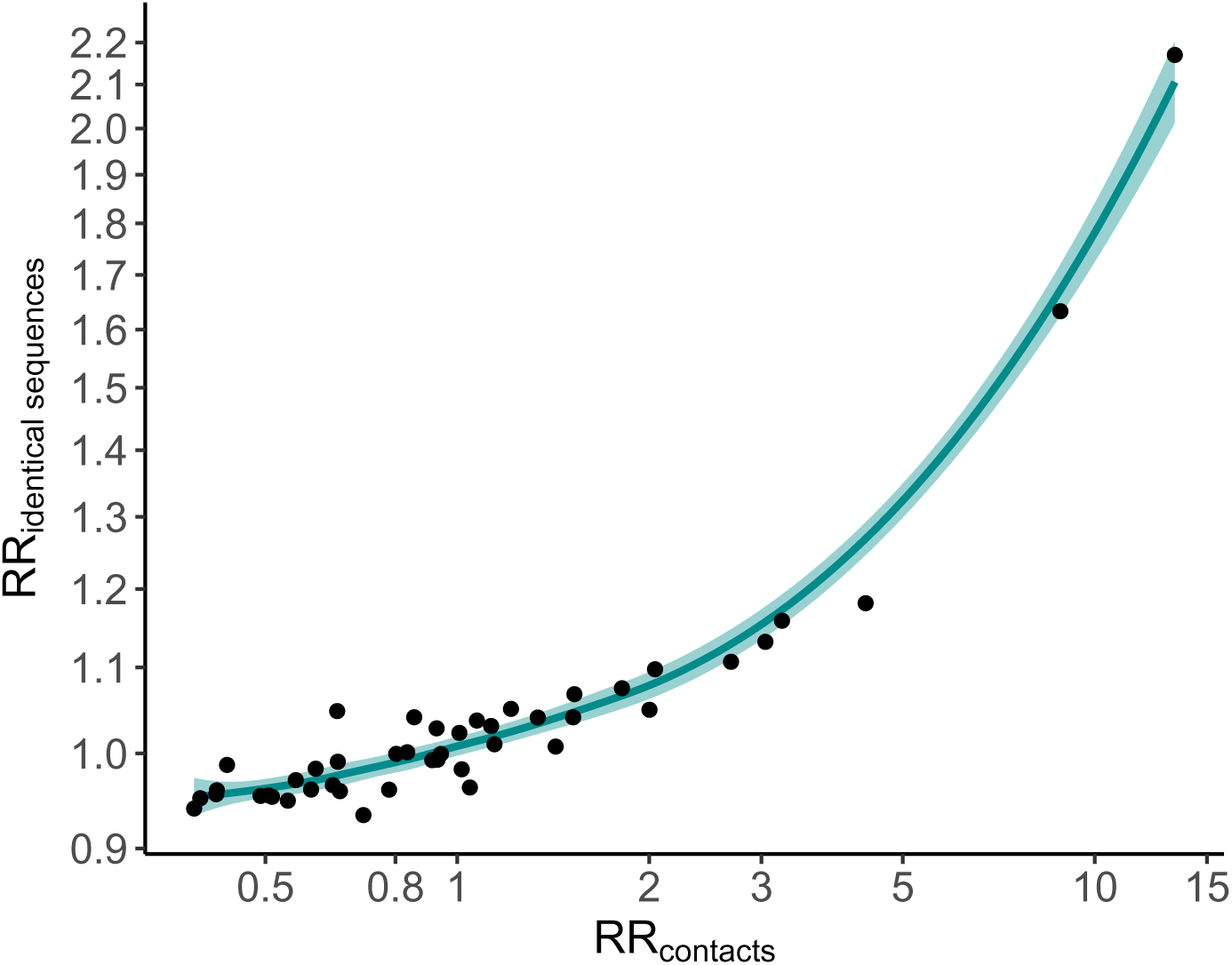
Expected relationship between the RR of observing identical sequences in two age groups and the RR of contacts between these age groups. These results were obtained by simulating 10^5^ clusters of identical sequences from a branching process with mutations [8] using a Poisson offspring distribution. The simulation was parametrised by a reproduction number of 1.2, a probability that an infector and an infectee have the same consensus sequence of 0.7 and a sequencing fraction of 10%.

**Figure S12.**
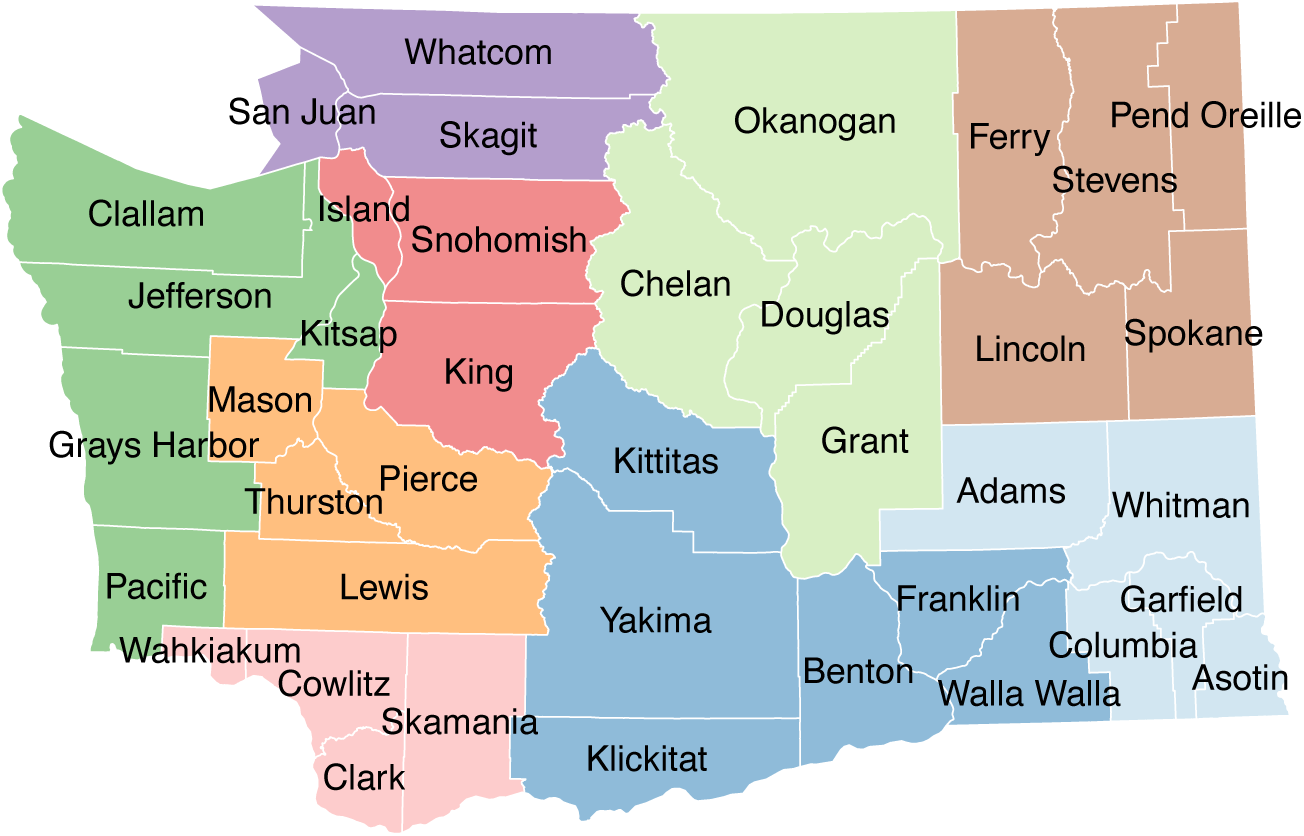
Geographical regions used to aggregate counties. Dark green: Peninsula/Coastal. Orange: South Puget Sound. Purple: Northwest. Red: North Puget Sound. Pink: Southwest. Dark blue: South Central. Light blue: Southeast.Brown: Northeast. Light green: North Central.

**Figure S13.**
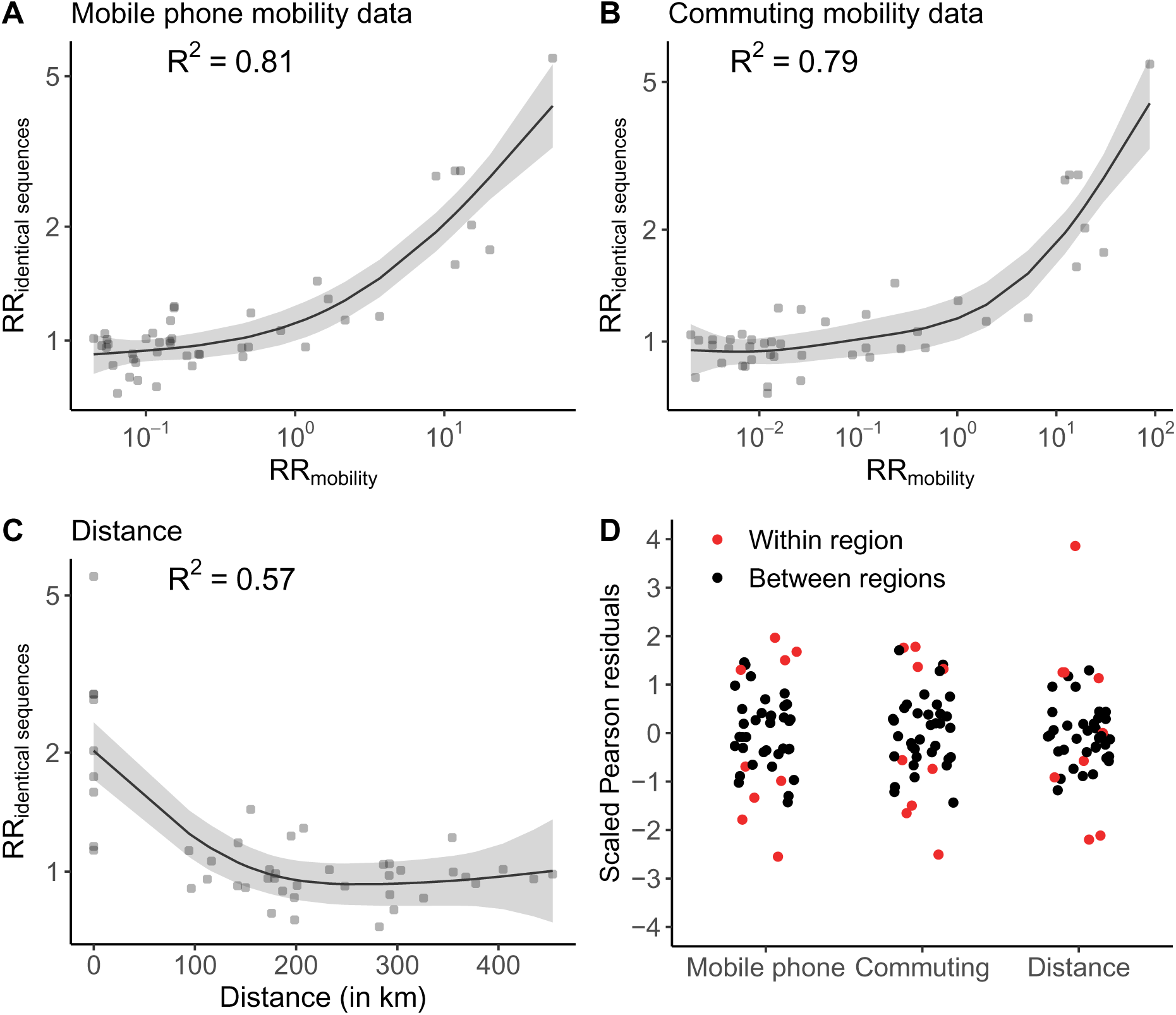
Comparison between the relative risk of observing identical sequences and the relative risk of movement at the region level. **A.** Relationship between the relative risk of observing identical sequences in two regions and the relative risk of movement between these regions as obtained from mobile phone mobility data. **B.** Relationship between the relative risk of observing identical sequences in two regions and the relative risk of movement between these regions as obtained from workflow mobility data. **C.** Relationship between the relative risk of observing identical sequences in two regions and the euclidean distance between region centroids. **D.** Scaled Pearson residuals of the GAM between the relative risk of observing identical sequences in two regions and (i) the relative risk of movement from commuting data, (ii) the relative risk of movement from mobile phone data and (iii) the geographic distance between regions’ centroids. The trend lines correspond to predicted relative risk of observing identical sequences in two regions from each GAM. *R*^2^ indicate the variance explained by each GAM.

**Figure S14.**
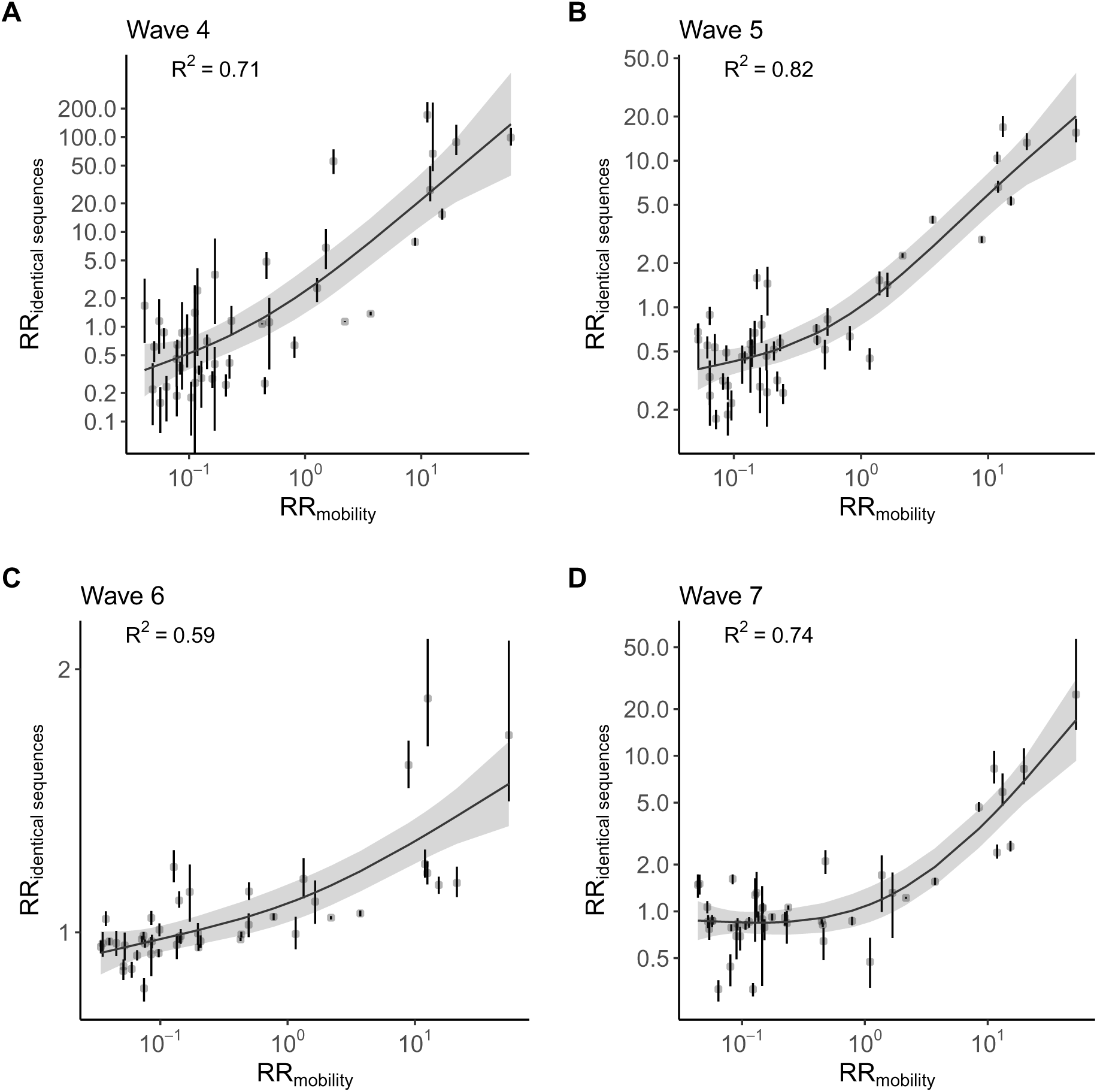
Relationship between the relative risk of observing identical sequences in two regions and the relative risk of movement between these regions obtained from mobile phone mobility data across epidemic waves. **A.** Wave 4. **B.** Wave 5. **C.** Wave 6. **D.** Wave 7. Vertical segments indicate 95% subsampling confidence intervals. The trend line correspond to predicted relative risk of observing identical sequences in two regions from a GAM. *R*^2^ indicate the variance explained by each GAM.

**Figure S15.**
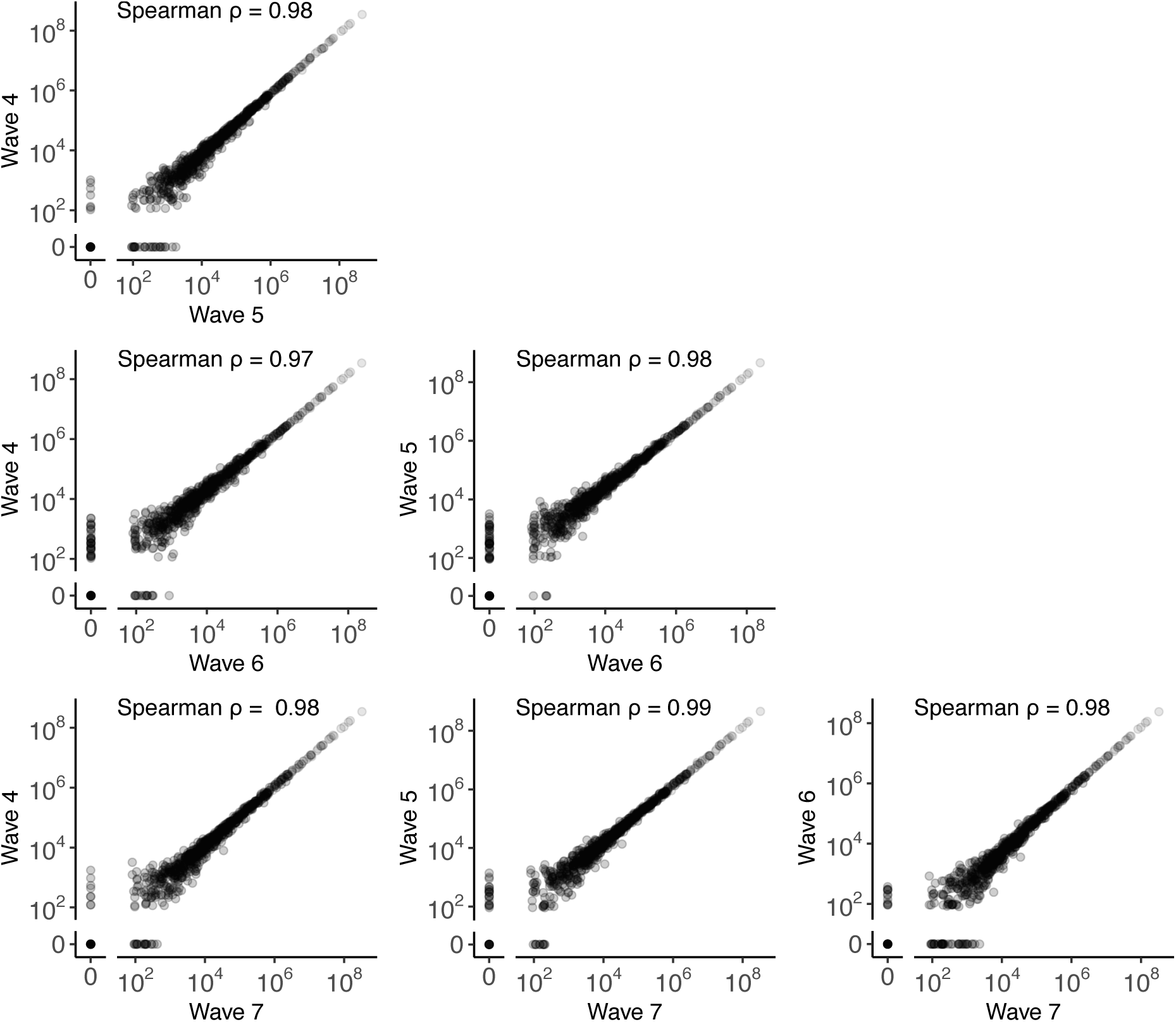
Comparison of the total number of visits between pairs of WA counties across the 4 epidemic waves during our study period. Points indicate the total number of visits between pairs of counties over the study periods labeled on the plot axes.

**Figure S16.**
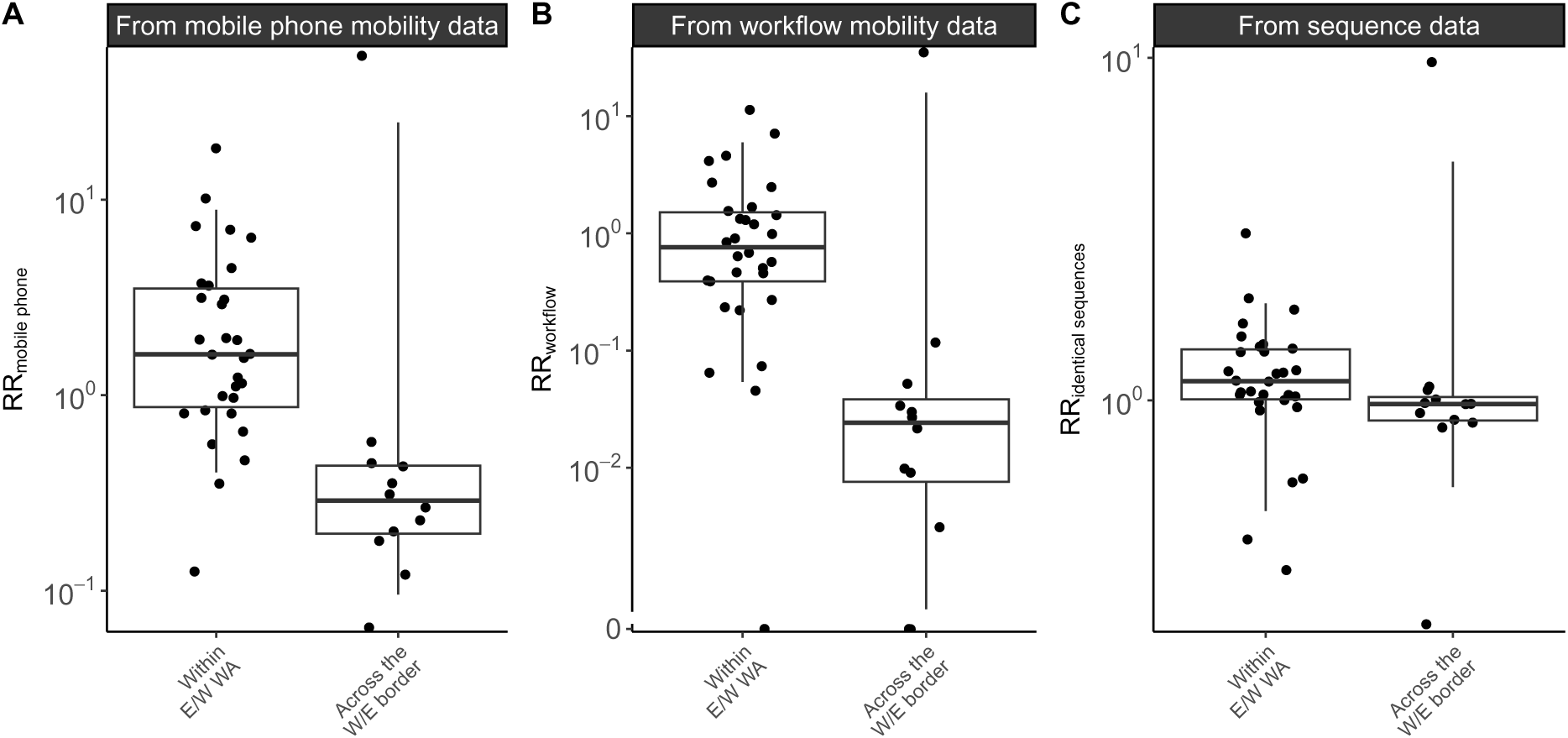
Comparison of connectivity metrics across the Eastern / Western WA border among counties located on the border. **A.** Relative risk of movement from mobile phone data across the border or within Eastern / Western WA (p-value for Wilcoxon rank sum test of 6.1 · 10^−5^). **B.** Relative risk of movement from commuting data across the border or within Eastern / Western WA (p-value for Wilcoxon rank sum test of 1.6 · 10^−4^). **C.** Relative risk of observing identical sequences across the border or within Eastern / Western WA (p-value for Wilcoxon rank sum test of 2.5 · 10^−2^). In this analysis, we only consider WA counties along the W/E border.

**Figure S17.**
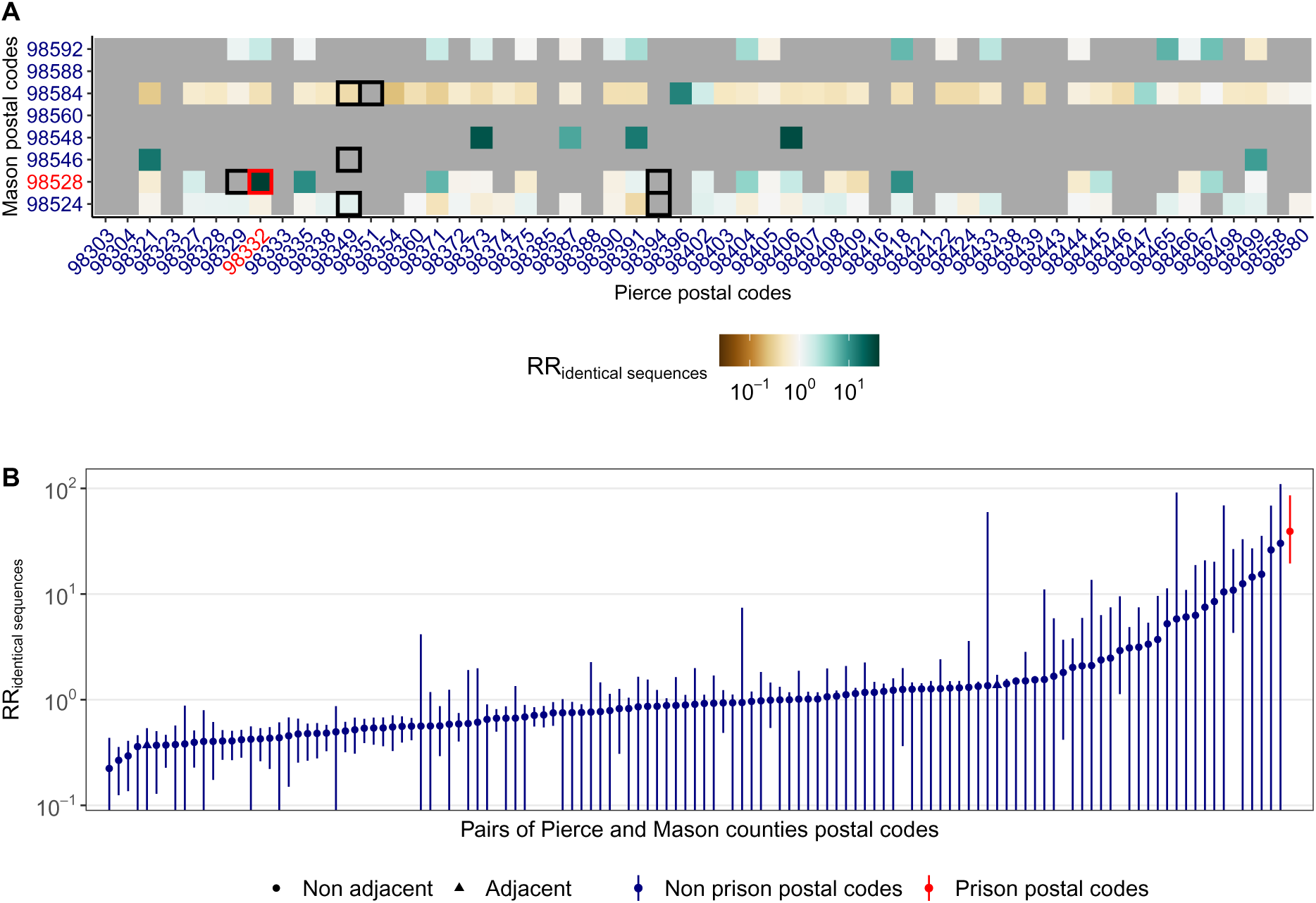
Patterns of occurrence of pairs of identical sequences between ZCTAs in Pierce and Mason counties, the two counties that are home of WA female prisons. **A.** Relative risk of observing identical sequences between ZCTAs in Mason and Pierce counties. Black squares indicate adjacent ZCTAs. ZCTAs in red correspond to postal codes that are the home of female prisons. **B.** Relative risk of observing identical sequences between Mason and Pierce counties ZCTAs ordered by increasing values. Vertical segments correspond to 95% subsampling confidence intervals.

**Figure S18.**
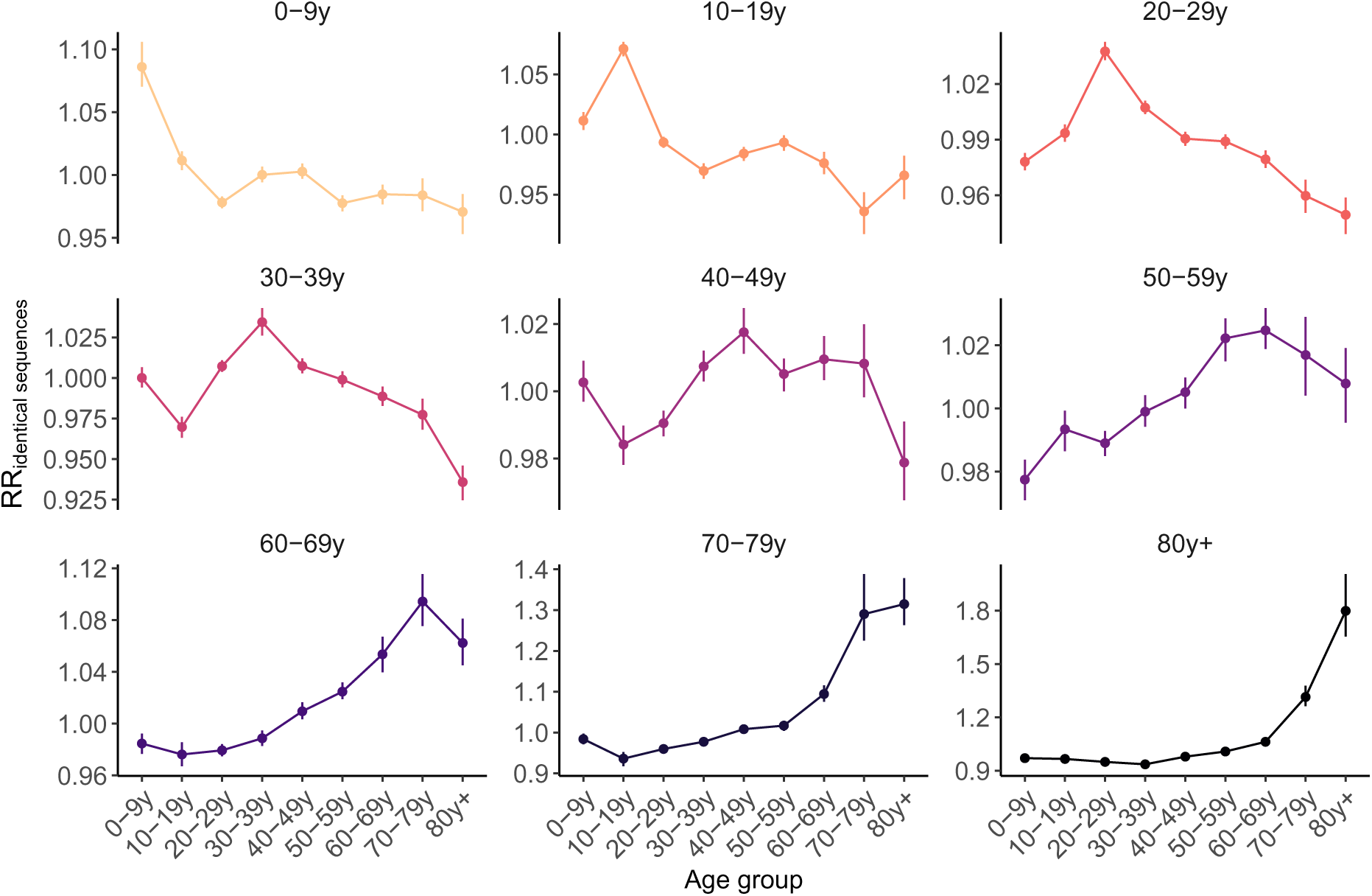
Relative risk for pairs of identical sequences of being observed between two age groups. Vertical segments correspond to 95% confidence intervals obtained through subsampling.

**Figure S19.**
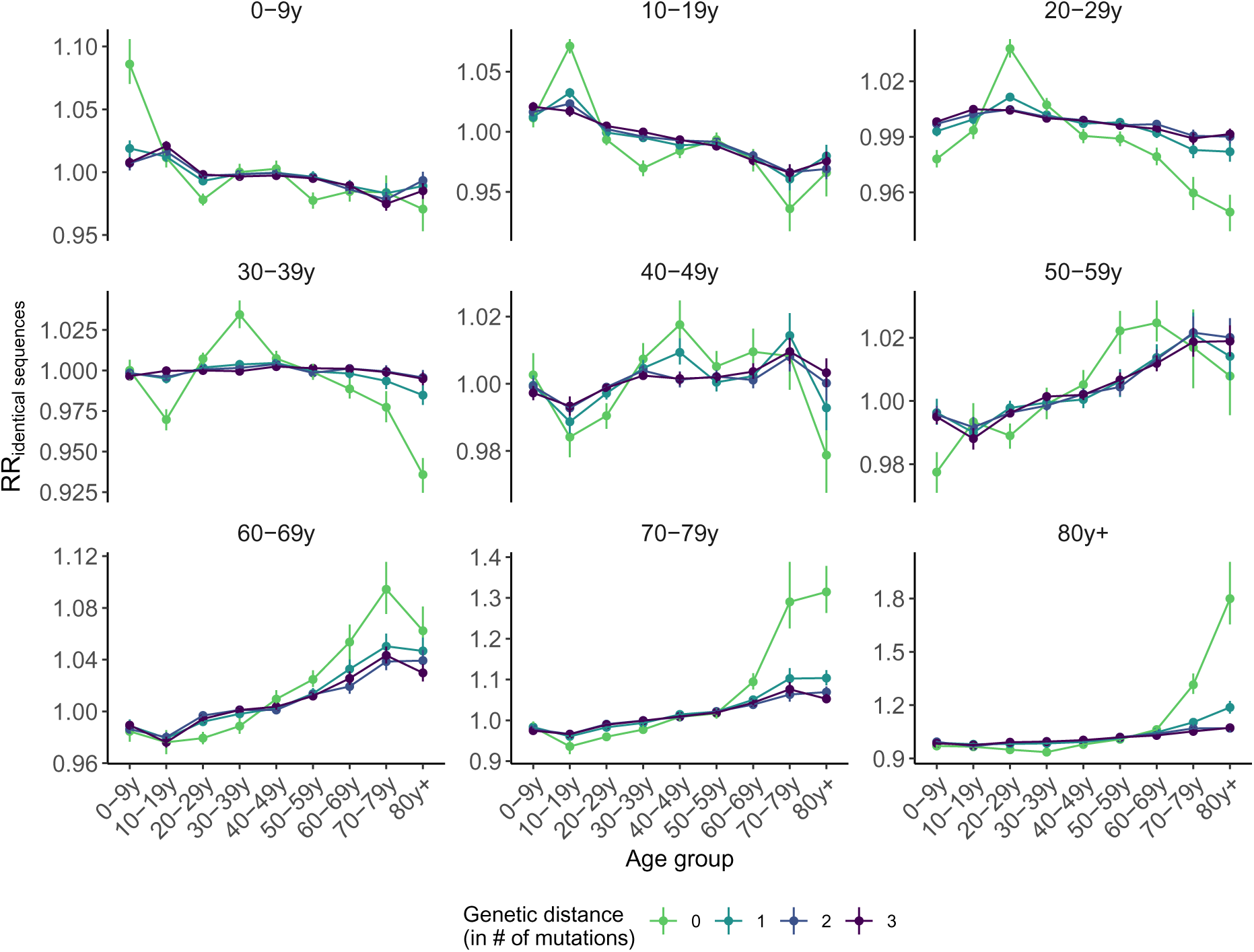
Relative risk for pairs sequences of being observed between two age groups depending on their genetic distance. Vertical segments correspond to 95% confidence intervals obtained through subsampling.

**Figure S20.**
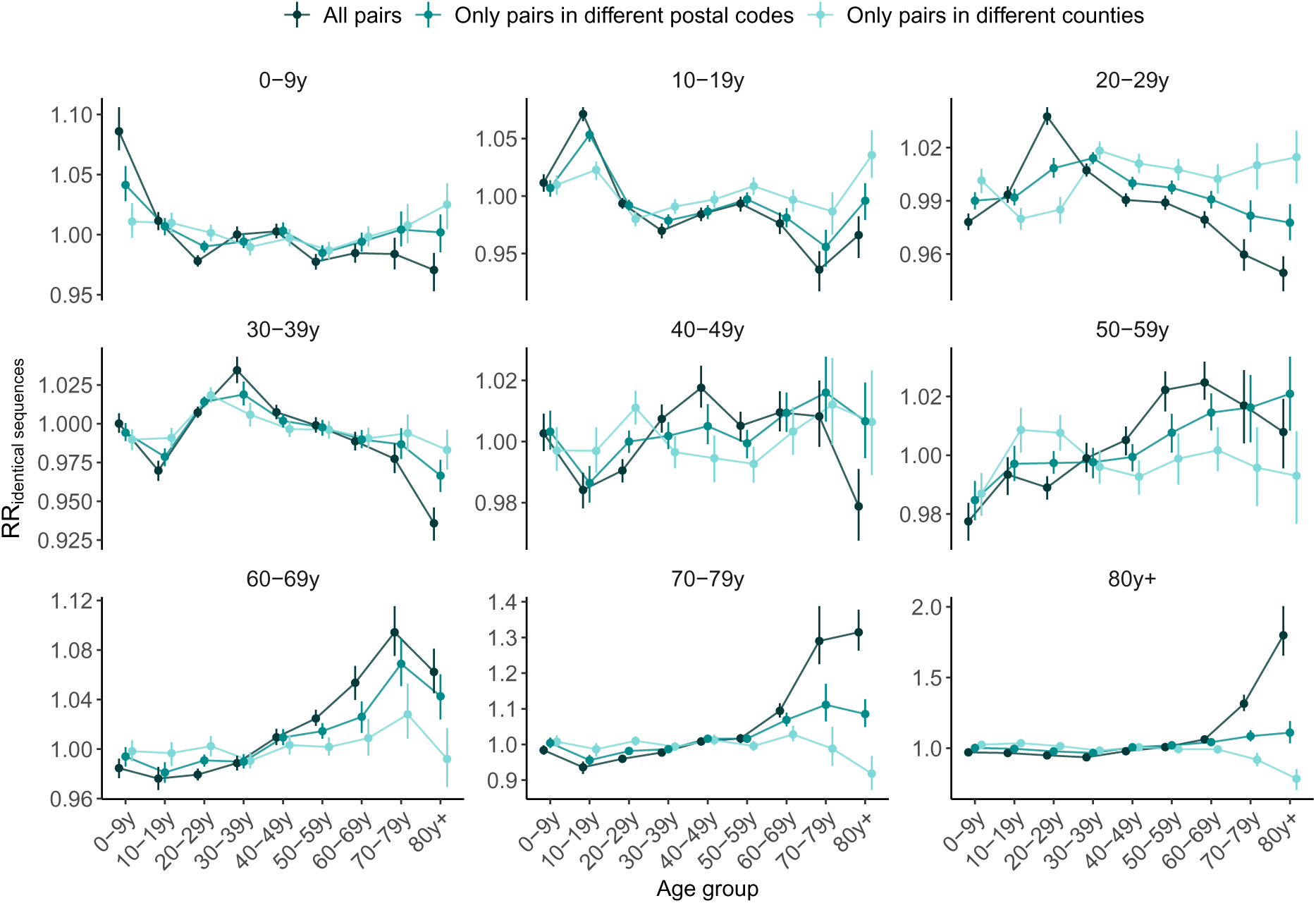
Impact of the spatial scale on the relative risk for pairs sequences of being observed between two age groups. Vertical segments correspond to 95% confidence intervals obtained through subsampling.

**Figure S21.**
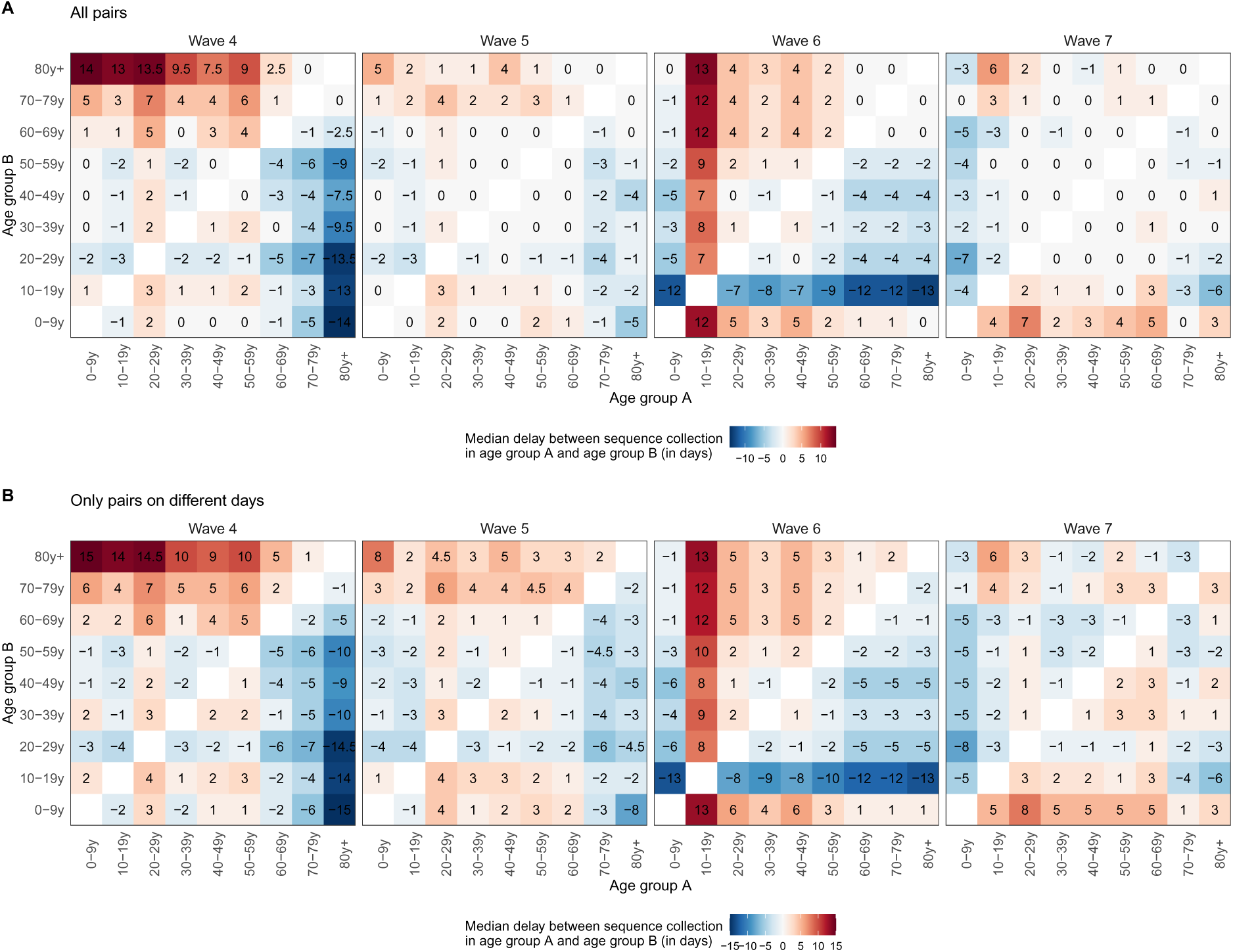
Median delay between the dates of sequence collection within pairs of identical sequences. **A.** considering all pairs of identical sequences collected in two age groups and **B.** considering only pairs of identical sequences collected on different days in two age groups.

**Figure S22.**
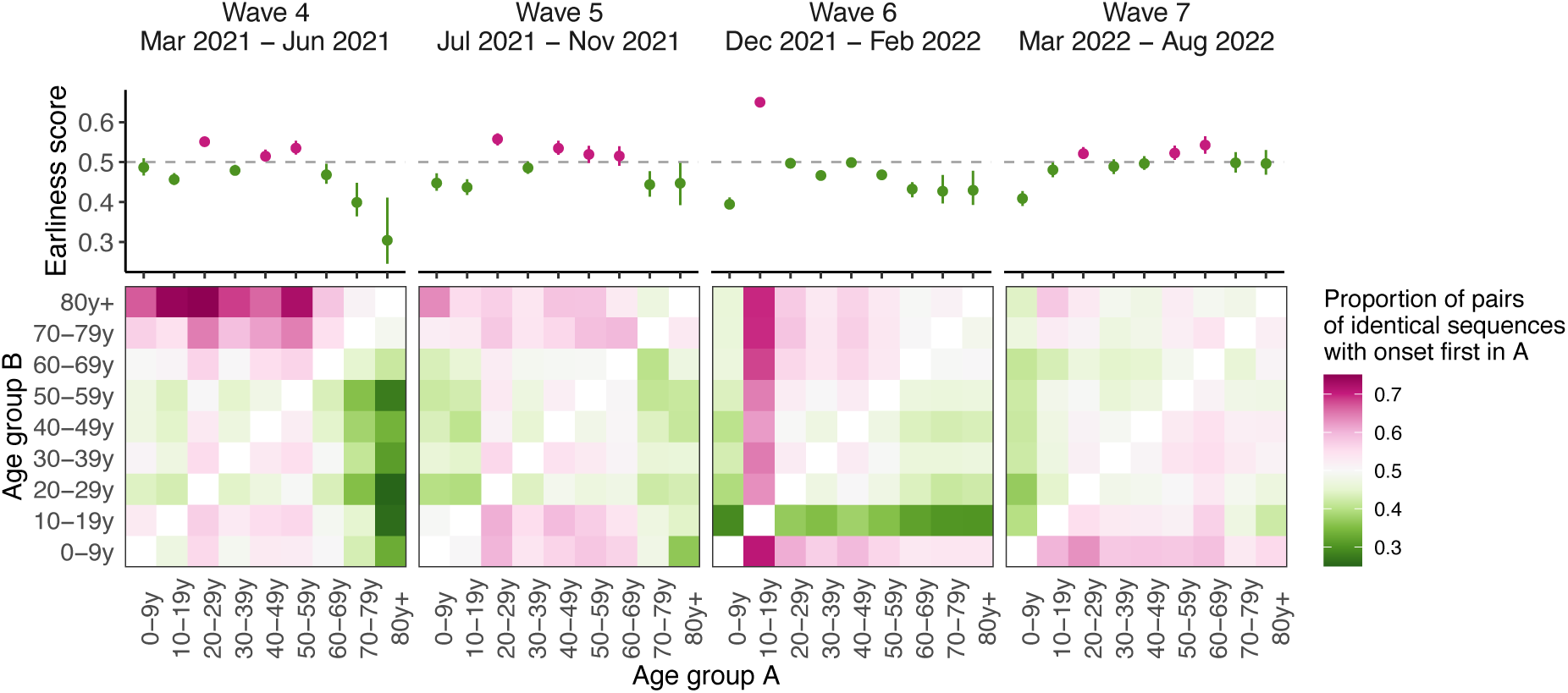
Sensitivity analysis on the timing of pairs identical sequences between age groups using symptom onset dates. Median proportion of pairs of identical sequences with onset dates in age groups A before age group B across different epidemic waves from 1,000 imputed datasets (heatmaps). The dots plots depict the median earliness scores of age group *A* across 1,000 imputed datasets for the different epidemic waves. Vertical segments indicate uncertainty range around earliness score (see Methods).

**Figure S23.**
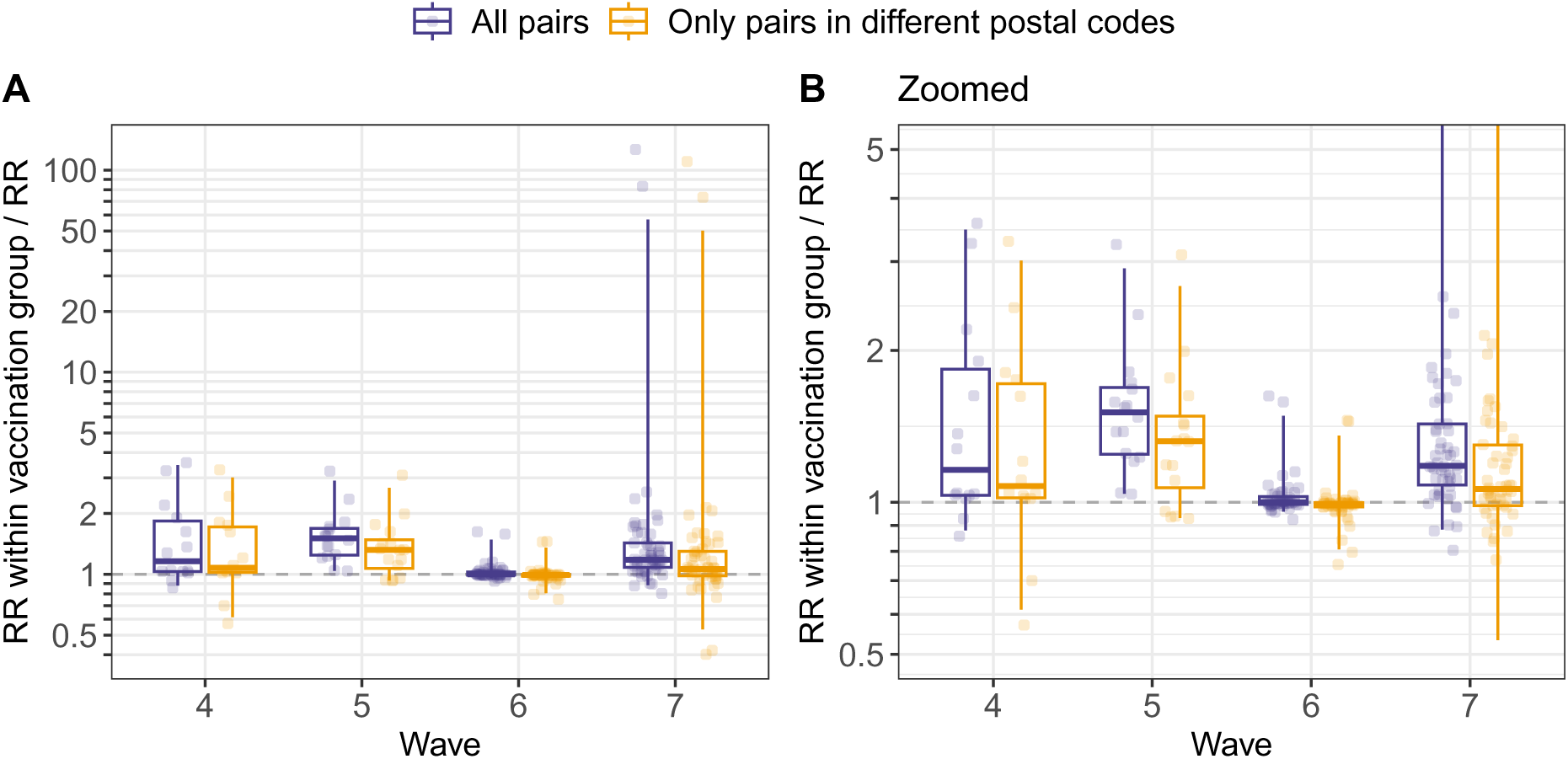
Ratio between the relative risk of observing identical sequences within a given vaccination group (denoted *V*_1_) and between two vaccination groups (denoted *V*_1_ and *V*_2_). Values above 1 indicate that pairs of identical sequences tend to be enriched in pairs observed within the same vaccination group. The analysis is restricted to pairs observed within the same age group. Each point correspond to the ratio computed for a given pair of vaccination status (*V*_1_, *V*_2_) and age group. Boxplots indicate the 2.5%, 25%, 50%, 75% and 97.5% percentiles.

**Figure S24.**
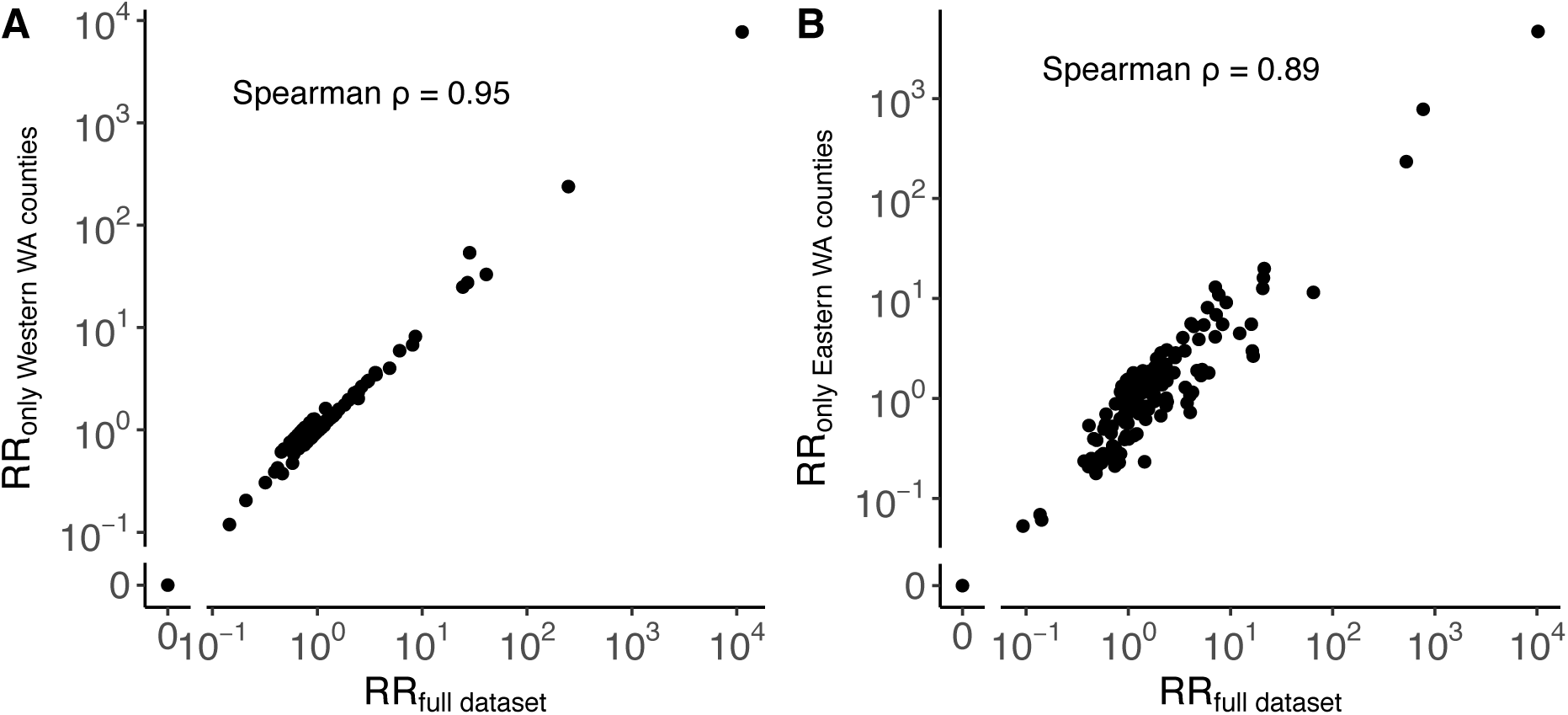
Impact of non-sampled locations on the computation of the RR. **A.** Comparison between the relative risk of observing identical sequences between Western WA counties using only sequence in Western WA counties or the entire sequence dataset. **B.** Comparison between the relative risk of observing identical sequences between Eastern WA counties using only sequence in Eastern WA counties or the entire sequence dataset.

**Figure S25.**
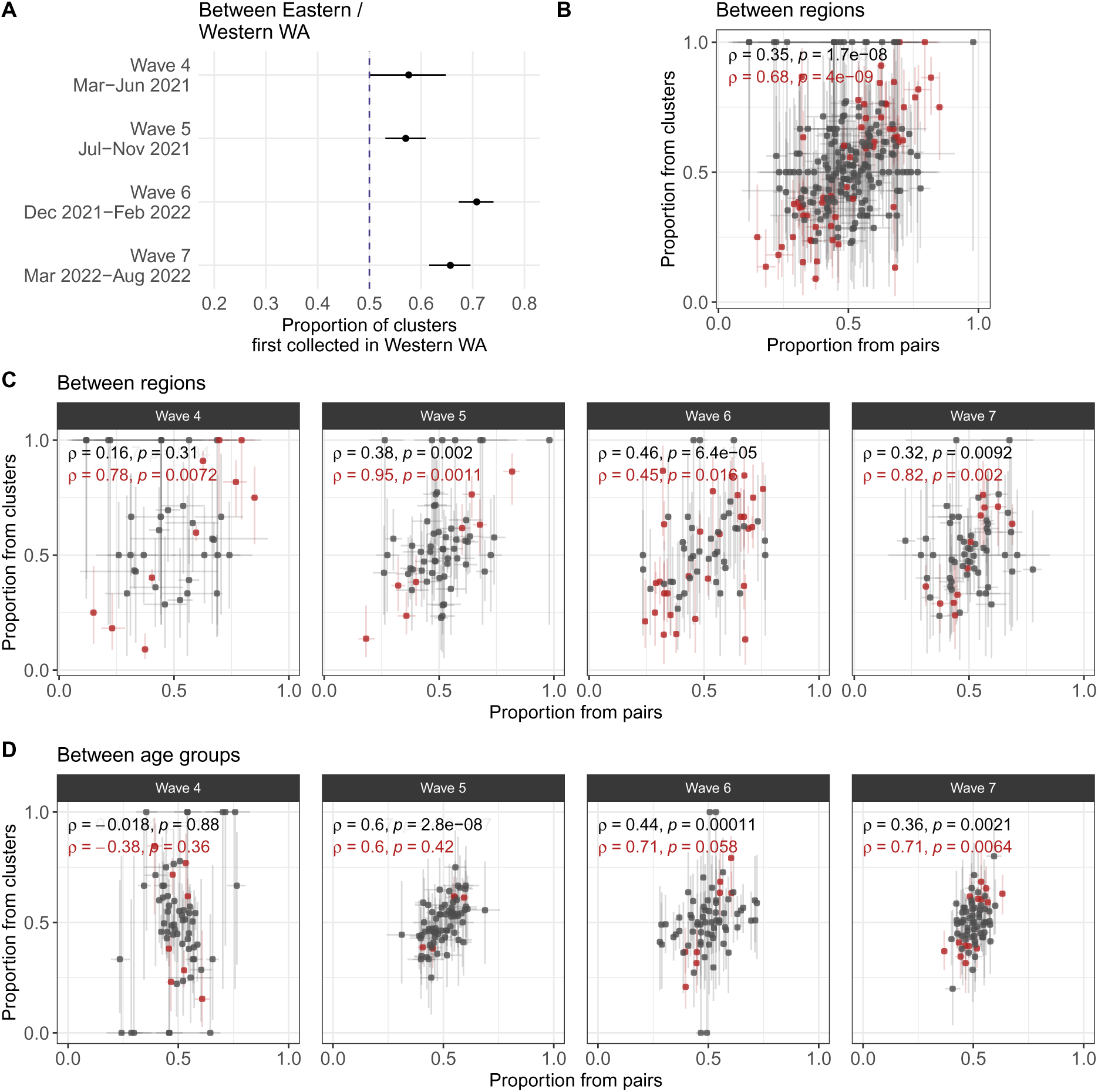
Sensitivity analysis for our transmission direction analysis relying on clusters of identical sequences observed only in two groups. **A.** Proportion of clusters first collected in Western WA among clusters observed in Eastern and Western WA and across pandemic waves. Like in Figure 2G, these proportions are all greater than 0.5. **B.** Sensitivity analysis at the regional level (Figure S12) comparing the proportion from pairs and the proportion from clusters. **C.** Sensitivity analysis at the regional level comparing the proportion from pairs and the proportion from clusters across waves. **D.** Sensitivity analysis at the age level comparing the proportion from pairs and the proportion from clusters across waves. For wave 4, the cluster based analysis relies on less than 10 clusters in 13 out of 36 pairs of age groups, which could explain the poor correlation. Segments indicate 95% CIs around proportions. In B, C and D, the colour red depicts points for which the CIs don’t cross 0.5 for both the proportion from clusters and the proportion from clusters. We report in red the Spearman correlation coefficient with p-values for these red points and in black for all the points.

**Figure S26.**
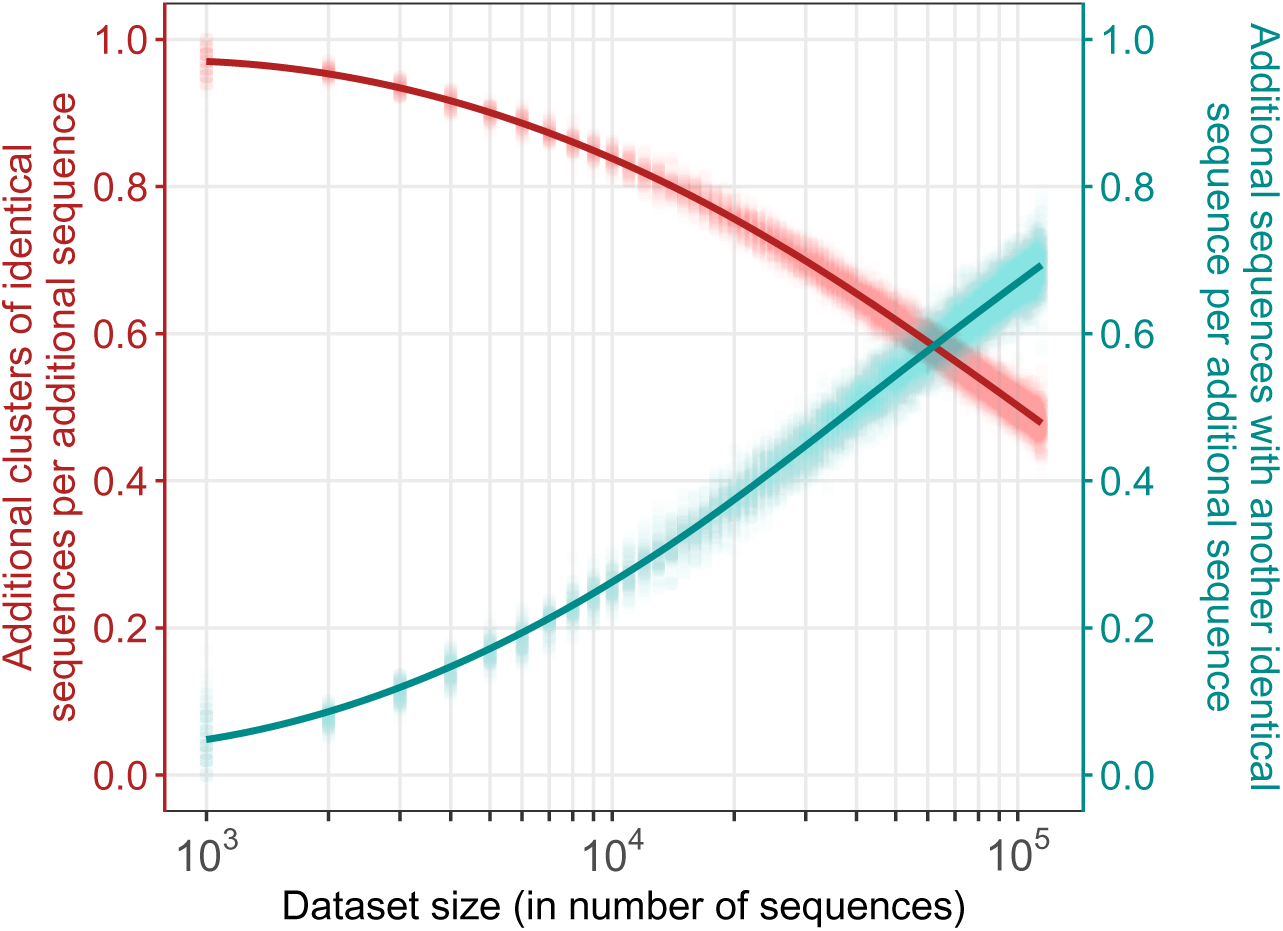
Impact of dataset size on the number of clusters of identical sequences and the number of sequences with another identical sequence in the dataset. We generate this figure by considering sequence datasets of increasing sizes, ranging between 10^2^ and the 114,298 (the size of our WA dataset) with an increment of 10^2^ between 10^2^ and 10^3^ and an increment of 10^3^ above 10^3^. We run 100 simulations where we first downsample 10^2^ sequences from our full dataset and then incrementally include more sequences (drawn from the total remaining sequences not yet included). At every step, we compute the additional number of clusters of identical sequences per additional sequences (red) as well as the additional number of sequences with another identical sequence in the dataset per additional sequence (cyan). Points indicate the results from individual simulations and lines the LOESS curves.

**Figure S27.**
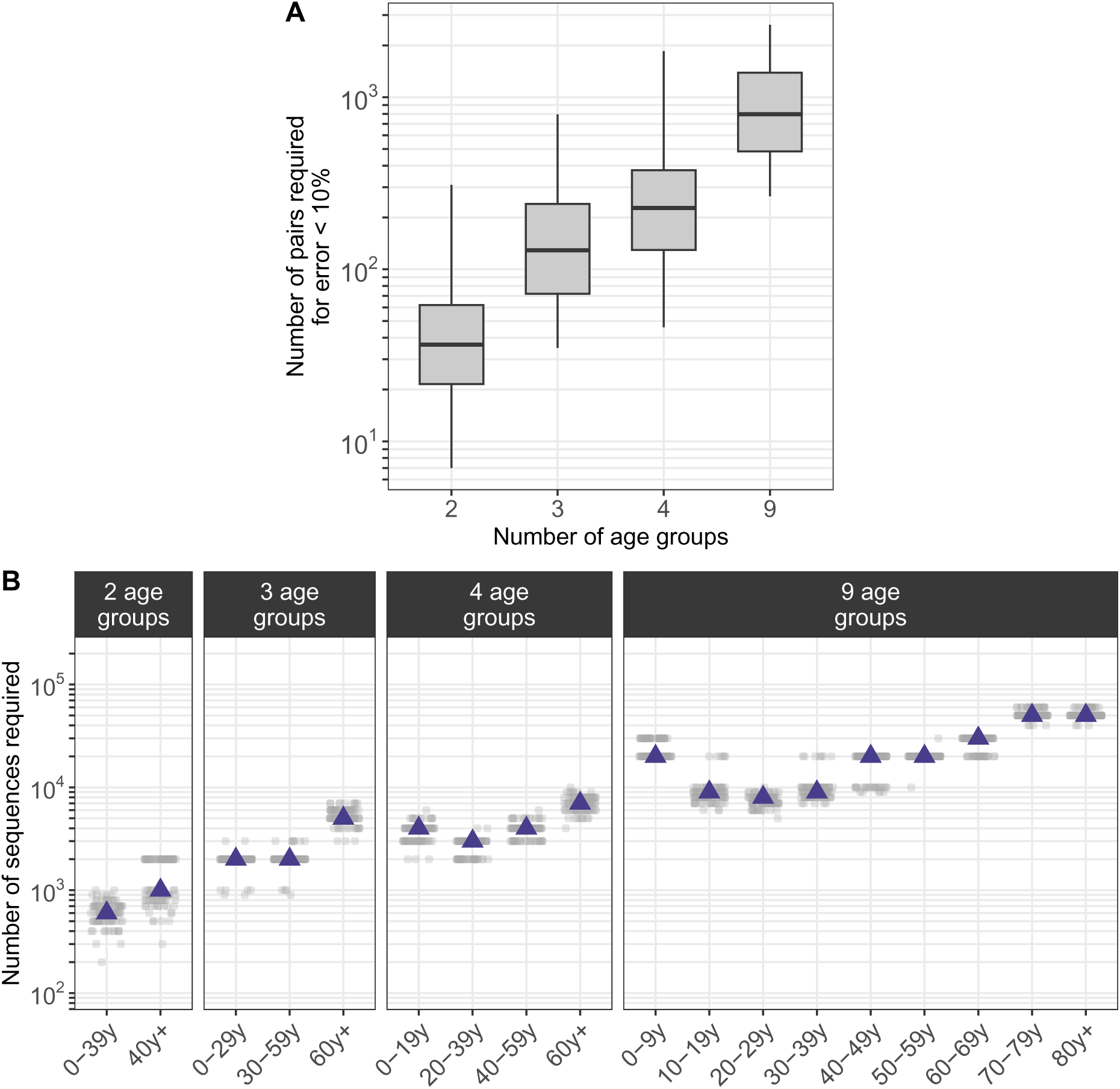
Impact of the number of groups included in the analysis on the dataset size required for the error in the relative risk of observing identical sequences to be lower than 10%. **A.** Number of pairs of identical sequences required for the error relative risk of observing identical sequences to be lower than 10%. Boxplots indicate the 2.5%, 25%, 50%, 75% and 97.5% percentiles. See Methods for a description of the downsampling strategy. **B.** Number of sequences required for the number of pairs of identical sequences observed within the age group on the x-axis to reach the median depicted in A. Each point corresponds to a subsampled dataset. Purple triangles indicate the median.

**Figure S28.**
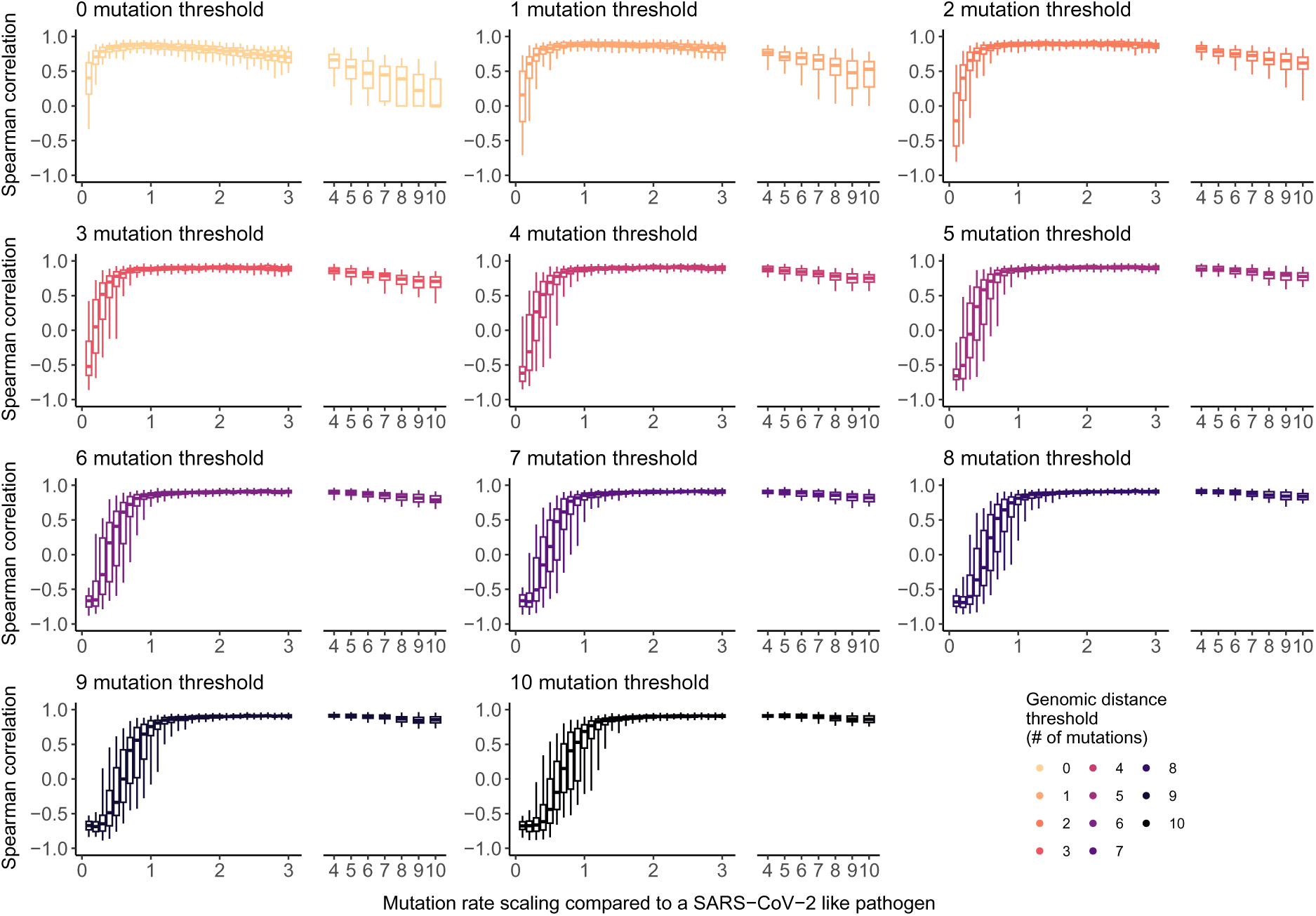
Impact of the pathogen’s mutation rate on the optimal Hamming distance threshold to apply our RR framework. Boxplots indicate Spearman correlation coefficient between the relative risk of pairs of sequences below the genomic distance threshold of being observed in two regions and the daily migration probability between these two regions. Boxplots indicate the 2.5%, 25%, 50%, 75% and 97.5% percentiles. See Methods for a description of the simulation approach.

**Figure S29.**
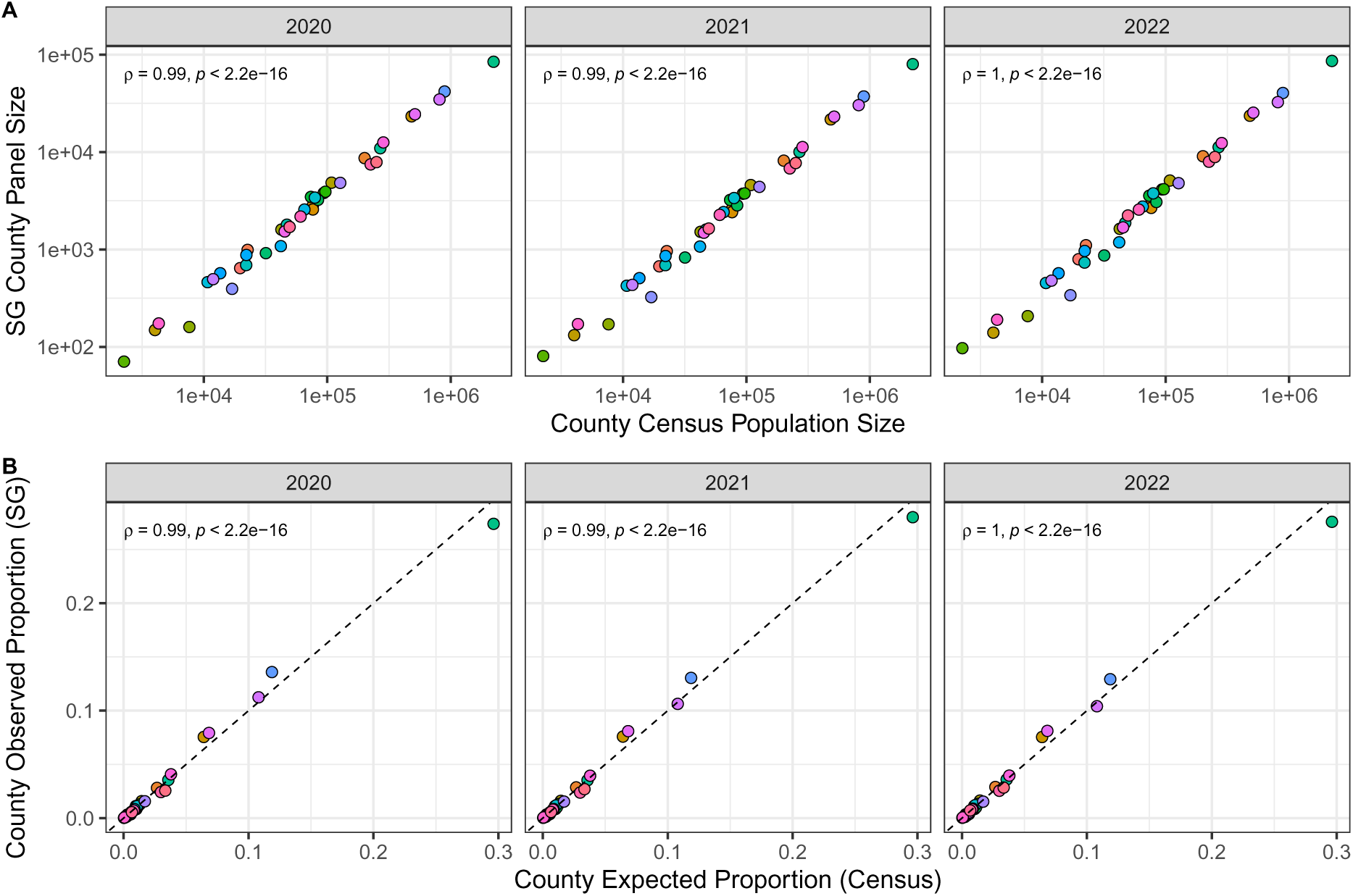
Comparisons between county census population sizes and SafeGraph panel sizes in Washington state, 2020 – 2022. **A.** County census population sizes strongly correlate with the mean number of devices tracked by SafeGraph (“SG”) in each year. **B.** Expected proportions of devices based on county and state census population sizes strongly correlate with the observed proportion of devices tracked by SafeGraph (“SG”) in each year. Points represent individual counties in WA state. In B, the black dashed line indicates the expected relationship for a true random sample of devices.

**Figure S30.**
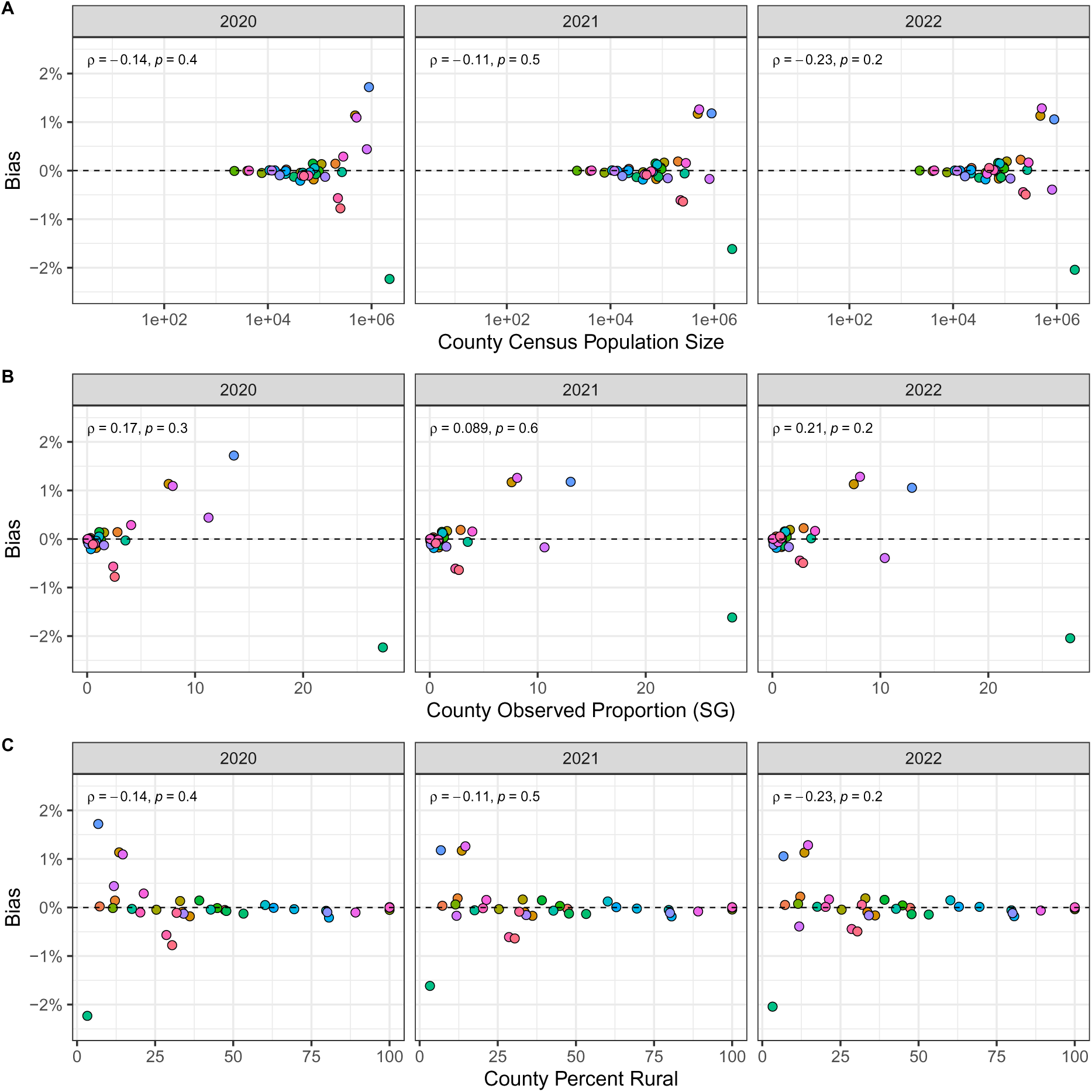
County-level bias of SafeGraph data in Washington state does not correlate with A. census population size, B. SafeGraph panel sizes in individual counties relative to WA state (“county observed proportion”), or C. census urban-rural classification, 2020 – 2022. Points represent individual counties in WA state. Bias is estimated as the “observed proportion” of devices tracked by SafeGraph in individual counties relative to WA state minus the “expected proportion” of devices based on census population sizes. Negative values indicate under-represented counties, and positive values indicate over-represented counties. The black dashed line (y = 0) indicates no bias in the SafeGraph panel of devices.

**Figure S31.**
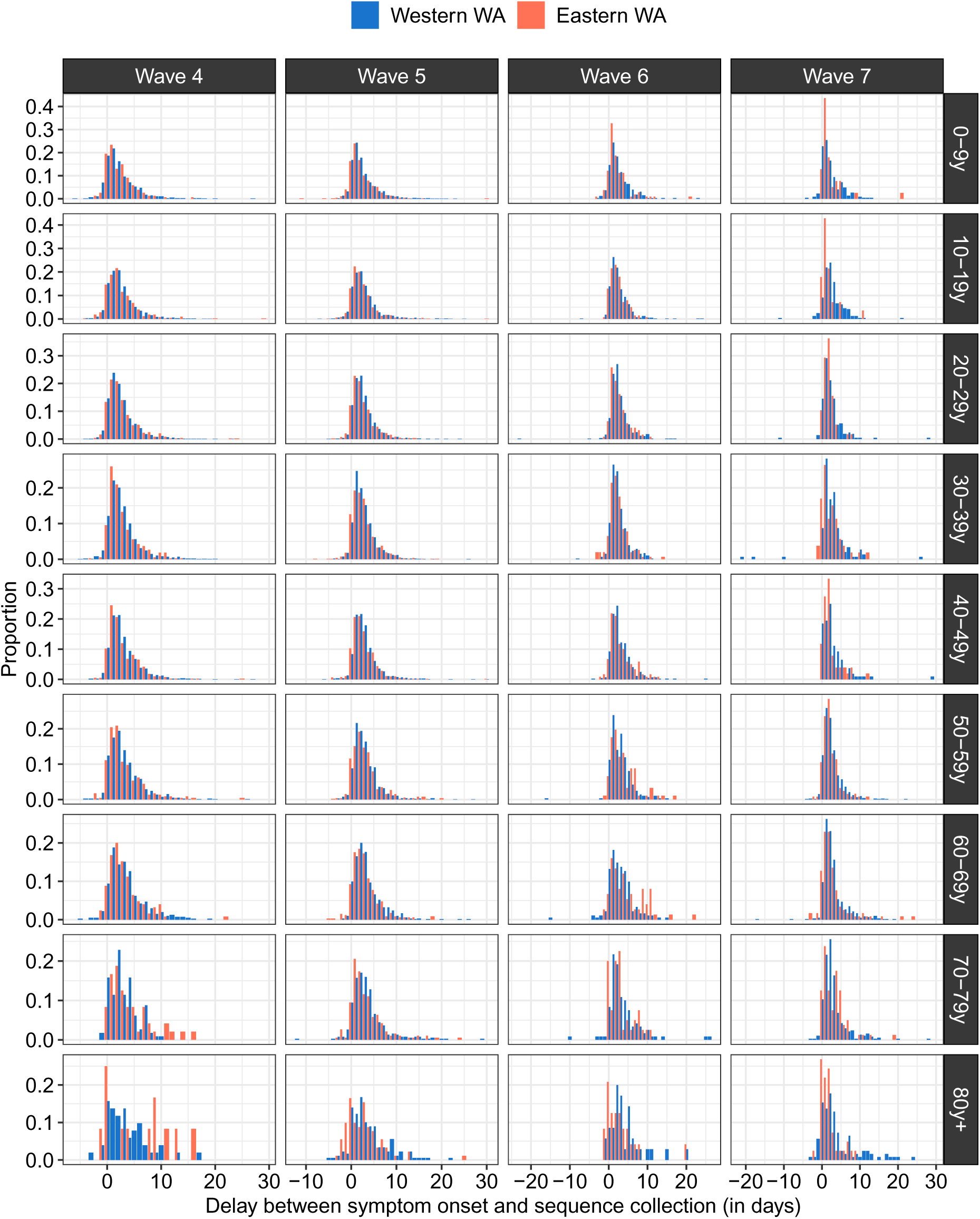
Empirical distribution of the delay between symptom onset and sequence collection by age (rows), period (columns) and geographic region (colours).

**Figure S32.**
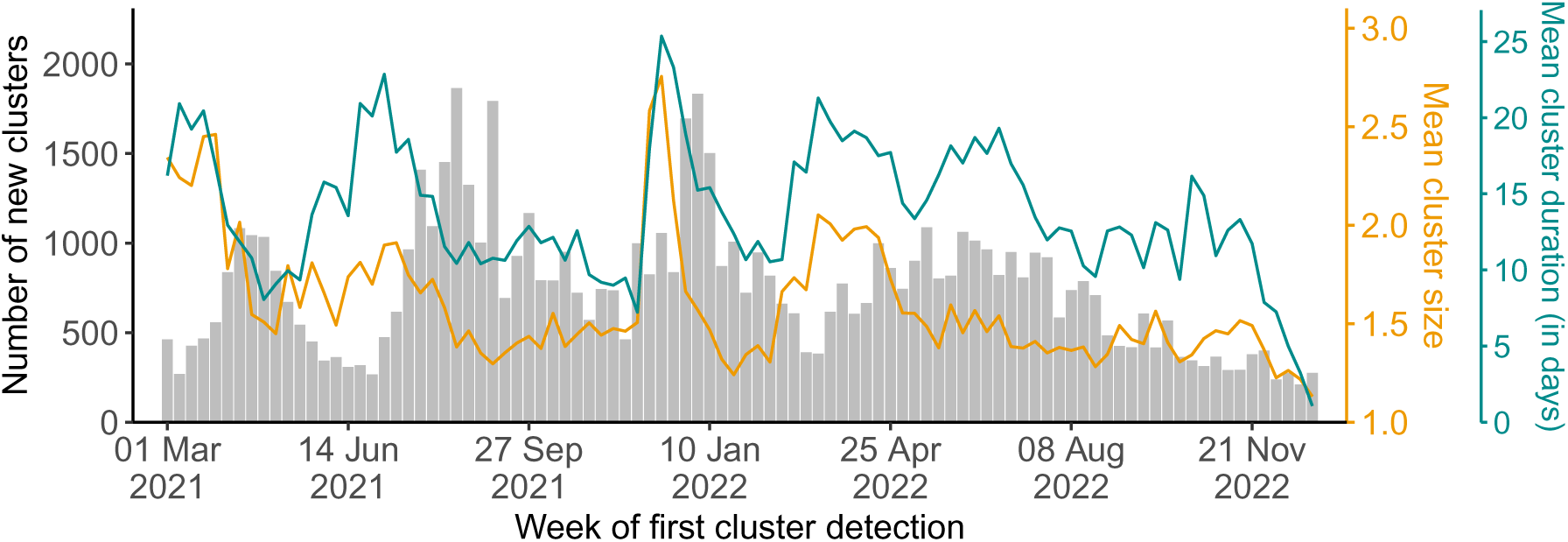
Characteristics of clusters of identical sequences across the study period. Grey bars (left y-axis) indicate the number of clusters by week of first cluster detection (defined as the week where the sequence with the earliest collection date was collected). The orange line (right orange y-axis) depicts the mean size of clusters of identical sequences by week of first cluster detection. The cyan line (right cyan y-axis) depicts the mean cluster duration by week of first cluster detection.

**Figure S33.**
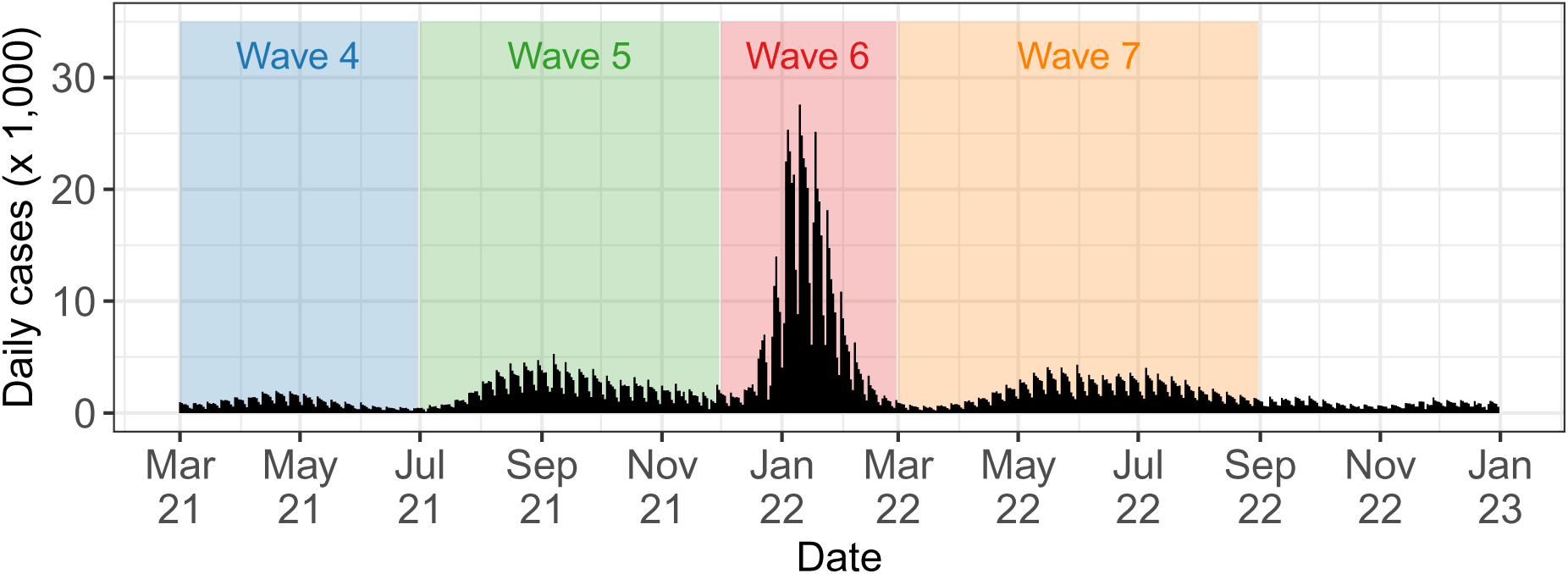
Time-series of COVID-19 cases in WA over the study periods. Shaded rectangles indicate the periods used to define the successive epidemic waves.

**Figure S34.**
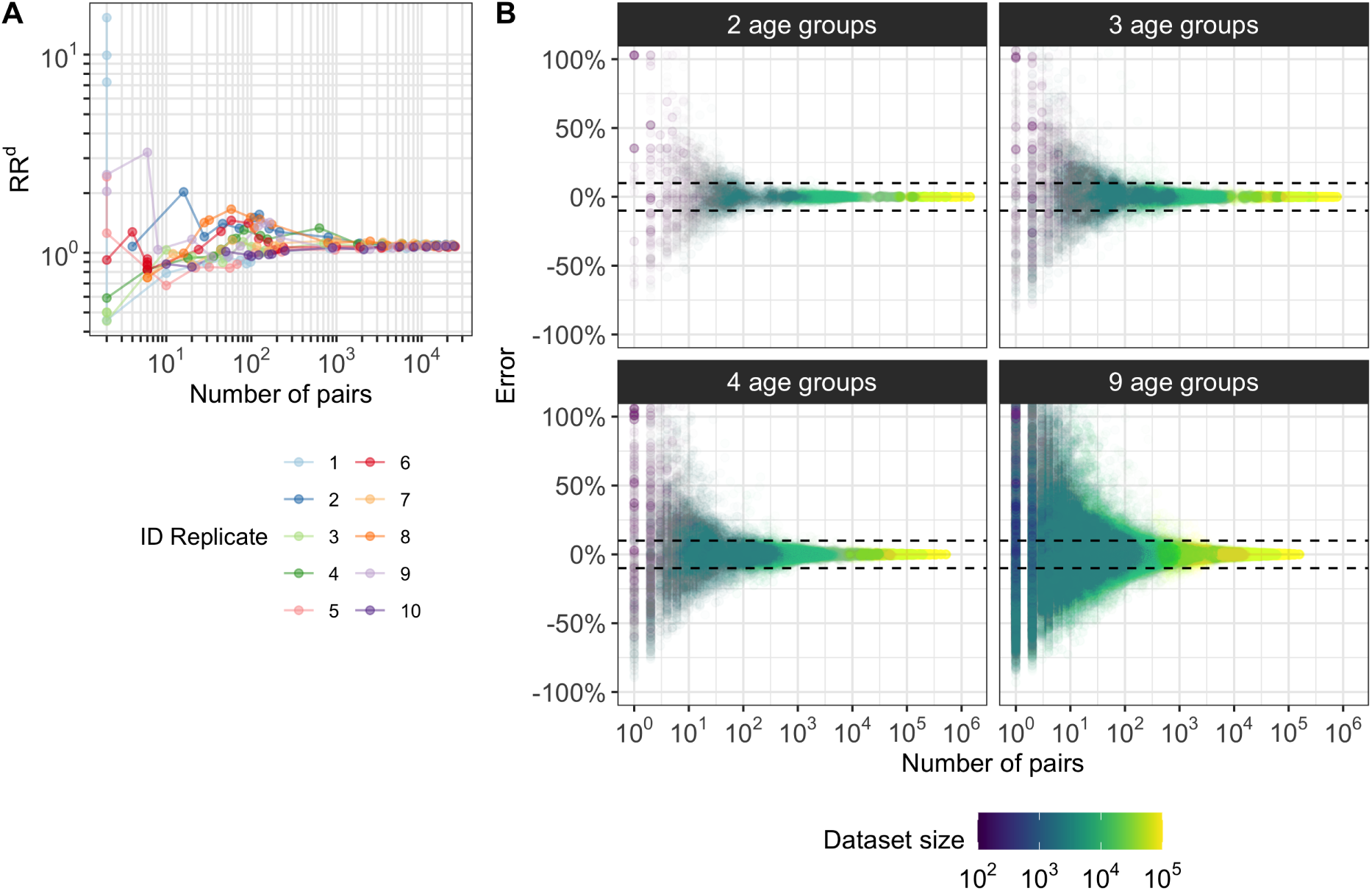
Illustration of the downsampling strategy used to quantify the amount of data required to compute relative risks. **A.** Relative risk *RR^d^* of identical sequences being shared between the 0-9y computed on 10 downsampled datasets as a function of the number of pairs of identical sequences shared between the 0-9y. **B.** Error *ɛ* on the relative risk of observing identical sequences in downsampled datasets as a function of the number of pairs of identical sequences present for a pair of age groups in the downsampled datasets.

**Table S1.**
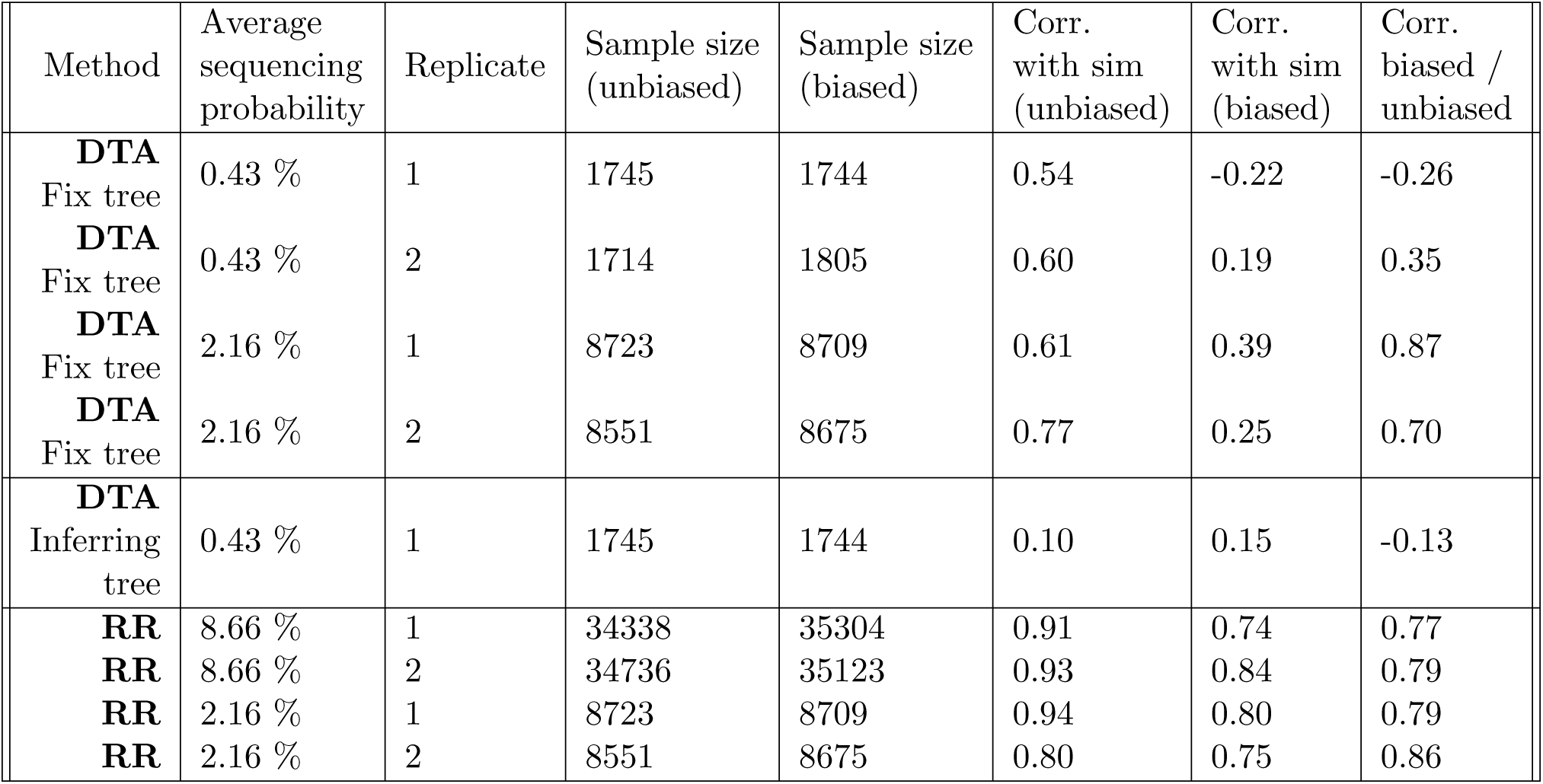
Performance of Discrete Trait Analysis (DTA) and our relative risk metric (RR) in quantifying migration patterns. The sample sizes correspond to the number of sequences on which the inference is performed. All correlation coefficients reported are Spearman rank correlation coefficients. In the DTA analysis, we report the correlation between estimated and true migration rates (both for the biased and unbiased sequencing scenarios) and the correlation between the migration rates estimated on the biased and unbiased datasets. In the RR analysis, we report the correlation between the RR and the migration probability between demes (both for the biased and unbiased sequencing scenarios) as well as the correlation between the RR estimated on the biased and unbiased datasets.

**Table S2.** Comparison of the relative of risk of observing identical sequences at the ZCTA level by adjacency level. We report the p-values of Wilcoxon rank sum test using either all pairs of ZCTAs or only pairs of ZCTAs for which pairs of identical sequences are collected (column “without 0”).

| Region | Adjacency status compared | p-value (Wilcoxon test) | p-value (Wilcoxon test) without 0 |
| --- | --- | --- | --- |
| East-East | Within ZCTA & Adjacent ZCTAs | $5 \cdot 10^{-10}$ | $3 \cdot 10^{-12}$ |
| | Adjacent ZCTAs & Non-adjacent ZCTAs | $< 10^{-16}$ | $< 10^{-16}$ |
| East-West | Adjacent ZCTAs & Non-adjacent ZCTAs | 0.73 | 0.39 |
| West-West | Within ZCTA & Adjacent ZCTAs | $< 10^{-16}$ | $< 10^{-16}$ |
| | Adjacent ZCTAs & Non-adjacent ZCTAs | $< 10^{-16}$ | $< 10^{-16}$ |

**Table S3.** Comparison between the relative risk of observing identical sequences between two geographic regions and the risk of movement between different geographies. We consider three data sources to inform the relative risk of movement between geographies: the relative risk for a visit to occur between two geographies (from mobile phone data), the relative risk for a work commute to occur between two geographies (from workflow data) and the geographic distance between geographies’ centroids.

|  | At the county level | At the region level |
| --- | --- | --- |
| *Spearman correlation $\rho$* |  |  |
| Mobile phone mobility | 35% | 61% |
| Workflow mobility data | 40% | 59% |
| Geographic distance | -35% | -48% |
| *Spearman correlation $\rho$ (without 0)* |  |  |
| Mobile phone mobility | 43% | 61% |
| Workflow mobility data | 56% | 59% |
| Geographic distance | -36% | -48% |
| *Variance explained (GAM)* |  |  |
| Mobile phone mobility | 60% | 81% |
| Workflow mobility data | 70% | 79% |
| Geographic distance | 32% | 57% |

**Table S4.** Characteristics of WA male prisons.

| Facility name | County | Prison capacity | County population size | Ratio prison capacity / county population |
| --- | --- | --- | --- | --- |
| Washington State Penitentiary | Walla Walla | 2439 | 62584 | $3.90 \cdot 10^{-2}$ |
| Stafford Creek Corrections Center | Grays Harbor | 1936 | 75636 | $2.56 \cdot 10^{-2}$ |
| Coyote Ridge Corrections Center | Franklin | 2468 | 96749 | $2.55 \cdot 10^{-2}$ |
| Washington Corrections Center | Mason | 1268 | 65726 | $1.93 \cdot 10^{-2}$ |
| Clallam Bay Corrections Center | Clallam | 858 | 77155 | $1.11 \cdot 10^{-2}$ |
| Airway Heights Corrections Center | Spokane | 2258 | 539339 | $4.19 \cdot 10^{-3}$ |
| Olympic Corrections Center | Clallam | 272 | 77155 | $3.53 \cdot 10^{-3}$ |
| Monroe Correctional Complex | Snohomish | 2400 | 827957 | $2.90 \cdot 10^{-3}$ |
| Cedar Creek Corrections Center | Thurston | 480 | 294793 | $1.63 \cdot 10^{-3}$ |
| Larch Corrections Center | Clark | 240 | 503311 | $4.77 \cdot 10^{-4}$ |

## Supplementary text 1: Relationship between the number of transmission pairs and the number of pairs with timing consistent with a transmission direction

### Notations

Let *N_X_*_→*Y*_ denote the number of transmission pairs where the infector is in subgroup *X* and the infectee in subgroup *Y*. Let *N_X<Y_* denote the number of transmission pairs between subgroups *X* and *Y* (regardless of the transmission direction) where the timing of symptom onset is earlier in *X* than in *Y*. Let *N_XY_* denote the total number of transmission pairs between subgroups *X* and *Y* (regardless of the transmission direction). Let *p_X_*_→*Y*_ = *N_X_*_→*Y*_ */N_XY_* denote the proportion of transmission pairs between *X* and *Y* that were in the direction *X* → *Y*. Let *p_X<Y_* = *N_X<Y_ /N_XY_* denote the proportion of transmission pairs between *X* and *Y* where the timing of symptom onset is earlier in *X* than in *Y*.

We introduce *p*_0_ as the proportion of transmission events with positive serial intervals (defined by the delay between the onset of symptom of the infectee and the infector).

### Relationship between *p_X<Y_* and *p_X_*_→_*_Y_*

Here, we demonstrate that comparing *N_X_*_→_*_Y_* and *N_Y_* _→_*_X_* is equivalent to comparing *N_X<Y_* and *N_Y <X_* as long as *p*_0_ is greater than 50%.

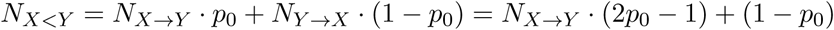

By diving the two sides of this equation by *N_XY_*, we have:

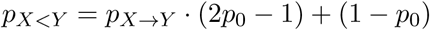

Therefore, if *p*_0_ *>* 0.5,

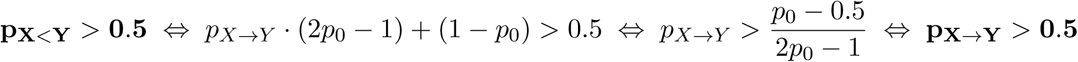

This means that as long as *p*_0_ is greater than 0.5, comparing the number of transmission pairs between *X* and *Y* with symptom onset dates first occurring in *X* to the transmission pairs between *X* and *Y* with symptom onset dates first occurring in *Y* provides direct insights into the proportion of transmission pairs between *X* and *Y* happening in the *X* → *Y* direction.

### Estimation of *p*_0_ for SARS-CoV-2

For a given pathogen, *p*_0_ can directly be estimated from a known serial interval distribution or from transmission pair data. For SARS-CoV-2, Geismar et al. [15] estimated this from reconstructed SARS-CoV-2 transmission events across a range of variants of concerns. In all analyses, their results show values greater of *p*_0_ greater than 50 %.

Here, we focus on the timing of symptom onset within transmission pairs. However, this argument is directly transposable to other timing definitions, such as the timing of sequence collection dates by replacing *p*_0_ by the proportion of transmission pairs where the sequence collection date of the infectee occurs after the sequence collection date of the infector.

